# Bayesian Anchored Latent Learning for Estimating State Trajectories

**DOI:** 10.64898/2026.09.30.26363325

**Authors:** Jason B. Gibbons, Nicolas Leenaerts, Amar D. Mandavia, Jacob C. Jameson, Annie Liang, David Hathaway, Jie Yang, Bowen Gu, Brian P. Marx, Shan Siddiqi, Rachel Wood, Matt Ruble, Stanley Lyndon, Philip S. Wang

**Affiliations:** Division of Pharmacoepidemiology & Pharmacoeconomics, Mass General Brigham, Harvard Medical School; National Center for PTSD at VA Boston Healthcare System; Department of Psychiatry, Mass General Brigham, Harvard Medical School; Department of Statistics, Harvard University; Department of Psychiatry, Boston University Chobanian & Avedisian School of Medicine; Department of Health Policy, Harvard Medical School; Discovery Behavioral Health

**Author notes:** These authors contributed equally as co-senior authors (Stanley Lyndon, Philip S. Wang). Corresponding author Jason B. Gibbons, PhD, Division of Pharmacoepidemiology and Pharmacoeconomics Department of Medicine Brigham and Women’s Hospital, Harvard Medical School 399 Revolution Drive, Boston, MA, USA 02145.

**Keywords:** Latent state estimation, state-space models, knowledge distillation, longitudinal data, sparse measurement, measurement-based care, psychiatric symptom trajectories

## Abstract

Dynamic systems often evolve faster than reliable measurements can be collected. More frequent indirect observations may fill these gaps, but their noise and differing scales complicate estimation of the underlying state. We developed Bayesian Anchored Latent Learning (BALL) to combine sparse reference measurements with denser records on an interpretable scale. A bidirectional transformer teacher reconstructs trajectories from complete training histories, and a forward-only transformer student learns from those reconstructions and observed measurements. Across 30 simulations, BALL reduced composite trajectory error by 7% relative to the same transformer trained directly from measurements, achieving lower error across all datasets. In an applied example using psychiatric records from 1,609 patients, BALL improved the prediction of withheld questionnaire scores compared with direct transformer training at 21- and 28-day measurement schedules. Requests guided by estimated uncertainty further improved prediction. BALL connects retrospective learning with prospective state estimation and provides a framework for directing additional measurement.

---

Many scientific processes have dynamic states that are sparsely measured over time, yet decisions about them require an accurate understanding of their trajectories. Additional records can help identify these states between reference measurements, but their observations are often noisy, incomplete, irregularly timed, and recorded on different scales. Combining these sources requires a model that estimates the changing state while preserving the interpretation supplied by the reference measurements.

Psychiatry is a particularly relevant example of this problem in measurement-based care, which uses repeated symptom assessments to guide treatment.^1–2^ Validated questionnaires such as the Patient Health Questionnaire-9 (PHQ-9), Beck Depression Inventory-II (BDI-II), and Generalized Anxiety Disorder-7 (GAD-7) provide clinically interpretable measures of depression and anxiety,^3–6^ but in routine care, these assessments are spaced across a treatment course, leaving gaps in the record of symptom change. Clinical notes, diagnoses, comorbidities, and treatment records provide additional information about patients’ health states between questionnaires but lack the same interpretability as the assessments.^7–8^ Questionnaires could define the state’s scale, with regularly recorded clinical information helping estimate change between assessments and provide a more detailed symptom trajectory.

State-space models offer a path forward by estimating a changing latent state from noisy observations,^9–11^ and modern sequence models extend this approach to high-dimensional, irregularly observed data.^12–18^ Prospective estimates must use only the history available at each session. The challenge is to transfer what complete training records reveal about trajectories into a model that estimates interpretable states as new observations arrive.

We developed Bayesian Anchored Latent Learning (BALL) to address this problem. In BALL, questionnaire measurement equations define the meaning and scale of latent depression and anxiety, while clinical observations, treatment history, and timing inform their trajectories. A bidirectional transformer teacher uses complete training records to reconstruct the state distribution at each session. A forward-only transformer student then learns from these distributions and observed questionnaires, with questionnaire links and teacher-fitted transitions held fixed during student training.^12, 19^ BALL combines these state estimates with uncertainty quantification and a transition model that uses fixed persistence and learned relationships between prespecified variables and subsequent symptom change (Fig. 1).

**Figure 1.**
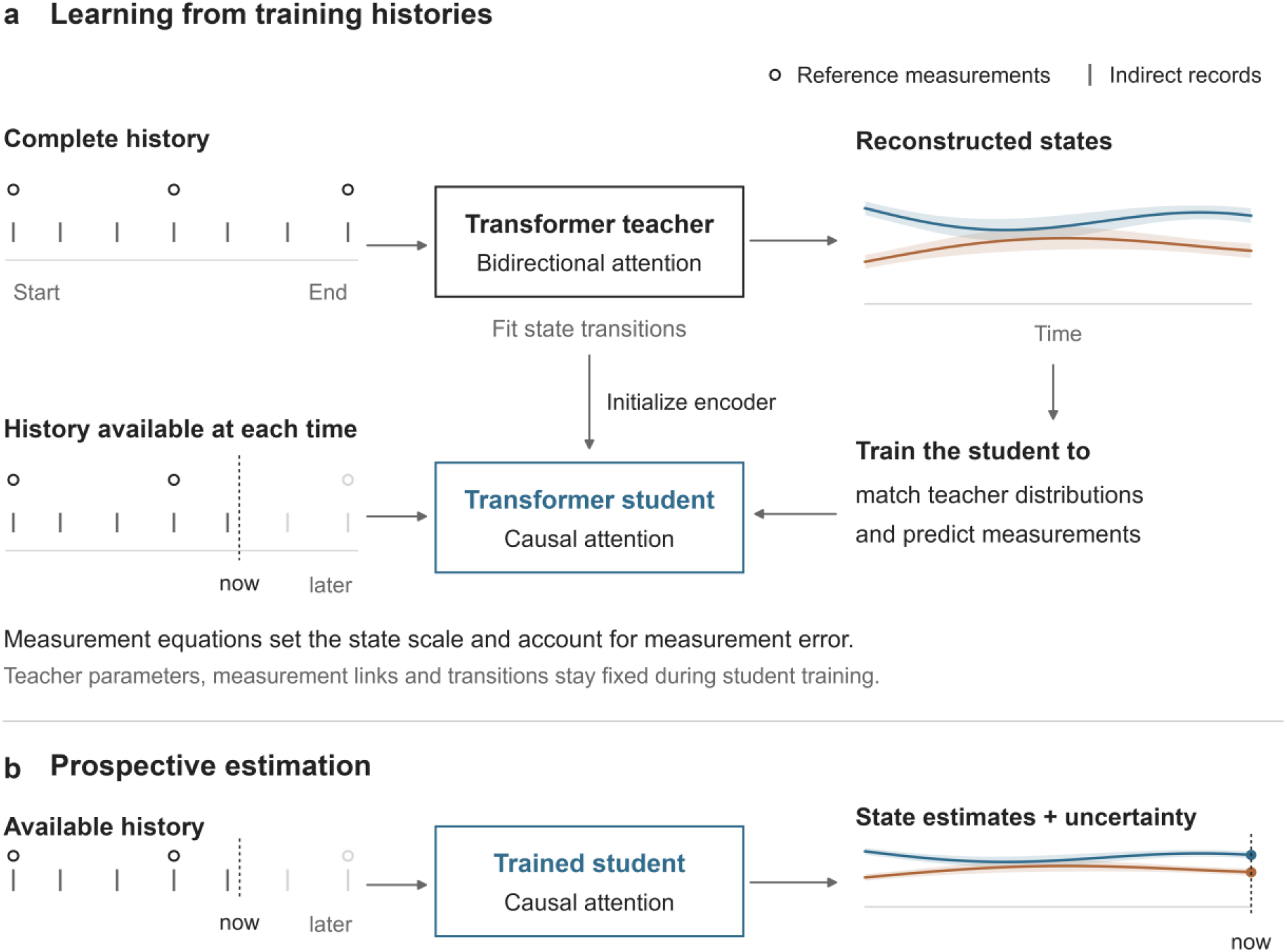
Learning interpretable states from complete histories for prospective estimation. **Notes**. a, Complete training histories provide sparse reference measurements and denser indirect records. A bidirectional transformer teacher reconstructs latent state distributions and is fitted jointly with state transitions. We copy the teacher parameters shared by the two architectures to initialize the forward-only student. We train the student using teacher-to-student Kullback– Leibler divergence between state distributions and the posterior-predictive likelihood of observed reference measurements. Instrument-specific measurement equations establish the state scale and account for measurement error. b, At each session, the trained student uses only available information. Faded symbols denote later observations, which are excluded. In the clinical application, blue and orange curves represent depression and anxiety, and questionnaires enter the history at the next session. Curves and uncertainty bands are schematic, not fitted results. The primary clinical model excludes RDoC scores. **Abbreviations**. BALL means Bayesian Anchored Latent Learning, and RDoC means Research Domain Criteria.

We evaluated BALL using simulations with known depression and anxiety trajectories and clinical records from patients receiving repetitive transcranial magnetic stimulation.^20–22^ The simulations assessed trajectory recovery, prediction of withheld questionnaires, and recovery of programmed relationships between Research Domain Criteria (RDoC) variables (transdiagnostic domains of mental illness with known brain-behavior relationships) and subsequent symptom change.^23–26^ Comparators included a direct transformer with the same forward architecture, a Gaussian process filter, a recurrent network, and linear-Gaussian and Markov state-space models.^9–11, 27–29^ Using ensemble, Bayesian, and conformal methods to quantify uncertainty,^30–35^ we evaluated clinical questionnaire prediction across four measurement schedules and examined whether uncertainty could guide additional questionnaire requests. A separate exploratory analysis tested whether earlier changes in RDoC content in clinical notes predicted subsequent BALL-estimated symptom change.

## Results

### Simulation evaluation - Recovery of known symptom trajectories and transition coefficients

#### Recovery of symptom trajectories and questionnaires

To test recovery of a changing state when the truth was known, we simulated depression and anxiety trajectories with sparse questionnaires, treatment histories, 24 partly observed clinical variables, and six noisy RDoC scores for 150 patients over 84 days. We compared six prospective models across five scenarios: linear RDoC effects on symptom change; treatment-dependent or nonlinear effects; effects that differed across patient subtypes; or increased missingness and measurement noise. Generating each scenario with weaker and stronger RDoC contributions, along with three random seeds, yielded 30 datasets. Using the mean of the two symptom states as a composite trajectory, BALL achieved the lowest mean root mean squared error (RMSE) among the six prospective methods, at 0.788 compared with 0.848 for the direct transformer and 1.419–2.074 for the other comparators. BALL had a lower composite error than the direct transformer across all datasets, reducing the mean RMSE by 7.0%. Improvements occurred in both channels, with depression-state RMSE of 0.888 versus 0.942 and anxiety-state RMSE of 0.885 versus 0.945. When predicting questionnaire scores withheld from these same datasets, BALL reduced RMSE from 1.122 to 1.070, with lower error in 29 of 30 datasets (Table 1, Figs. 2–3, and Supplementary Tables 1–4).

**Table 1.** Primary simulation performance across ten prespecified simulation settings.

| Model or component | Composite trajectory RMSE | Held-out questionnaire RMSE | Cosine similarity | Top-three F1 |
| --- | --- | --- | --- | --- |
| BALL student | 0.788 | 1.070 | — | — |
| BALL transition coefficient | — | — | 0.894 | 0.789 |
| Direct transformer | 0.848 | 1.122 | 0.914 | 0.800 |
| Exponential-decay gated recurrent unit | 1.419 | 1.161 | — | — |
| Causal Gaussian process | 1.856 | 1.684 | — | — |
| Linear Gaussian state-space model | 2.074 | 1.444 | 0.390 | 0.522 |
| Markov pattern-mixture model | 2.063 | 1.402 | 0.351 | 0.500 |

**Notes.** Values are descriptive means across 30 runs, spanning 10 prespecified settings and 3 seeds per setting. Lower RMSE indicates better prediction. All trajectory and questionnaire estimates use information available by the current session. Complete training records are used only to fit model parameters and train the teacher. The BALL teacher and student share a single transition coefficient, which is reported separately. Selecting three of the six domains gives an expected F1 of 0.50 by chance. Setting-specific results and Monte Carlo standard errors appear in Supplementary Table 2.
**Abbreviations.** Decay-GRU means exponential-decay gated recurrent unit, GP filter means Gaussian-process filter, BALL means Bayesian Anchored Latent Learning, F1 means the harmonic mean of precision and recall, RDoC means Research Domain Criteria, and RMSE means root mean squared error.

#### Comparison of BALL training conditions

To examine how student training influenced trajectory recovery, we reused the linear, nonlinear, and increased-missingness scenarios at the lower RDoC signal strength, each generated with three random seeds. Within these nine datasets, we compared students trained to match teacher distributions, predict questionnaires using fixed teacher-fitted dynamics, or do both through the full BALL procedure. Students trained to match the teacher, including full BALL students, started with teacher parameters. Students trained only on questionnaires started with random parameters. The full BALL training procedure achieved a composite trajectory RMSE of 0.731 and questionnaire RMSE of 1.076. Corresponding errors were 0.737 and 1.099 when students only matched the teacher’s distributions, and 0.770 and 1.111 when students learned from questionnaires with the teacher’s transitions held fixed. The full procedure, therefore, produced the lowest mean error for both targets (Supplementary Tables 5–6).

To assess whether the comparison depended on the optimization schedule, we trained BALL and the direct transformer using shared validation checks and stopping rules. These nine simulations spanned linear and nonlinear relationships, as well as increasing missingness, across three random seeds. BALL retained lower composite trajectory error in all nine runs and lower questionnaire error in six (Supplementary Table 4).

#### Recovery of programmed transition coefficients

We next asked whether the transition model could identify the variables programmed to influence subsequent changes in symptoms. In the primary simulations, three of the six RDoC domains contributed to that change, providing known coefficients against which to assess recovery. For the six RDoC coefficients, cosine similarity assessed alignment with the programmed direction of their effects on subsequent symptom change, and top-three F1 assessed identification of the three active domains. Cosine similarity and top-three F1 were 0.894 and 0.789 for BALL, 0.914 and 0.800 for the direct transformer, 0.390 and 0.522 for the linear Gaussian model, and 0.351 and 0.500 for the Markov model. Both transformers recovered the programmed direction and active domains more accurately than the classical models (Fig. 2 and Supplementary Table 7).

**Figure 2.**
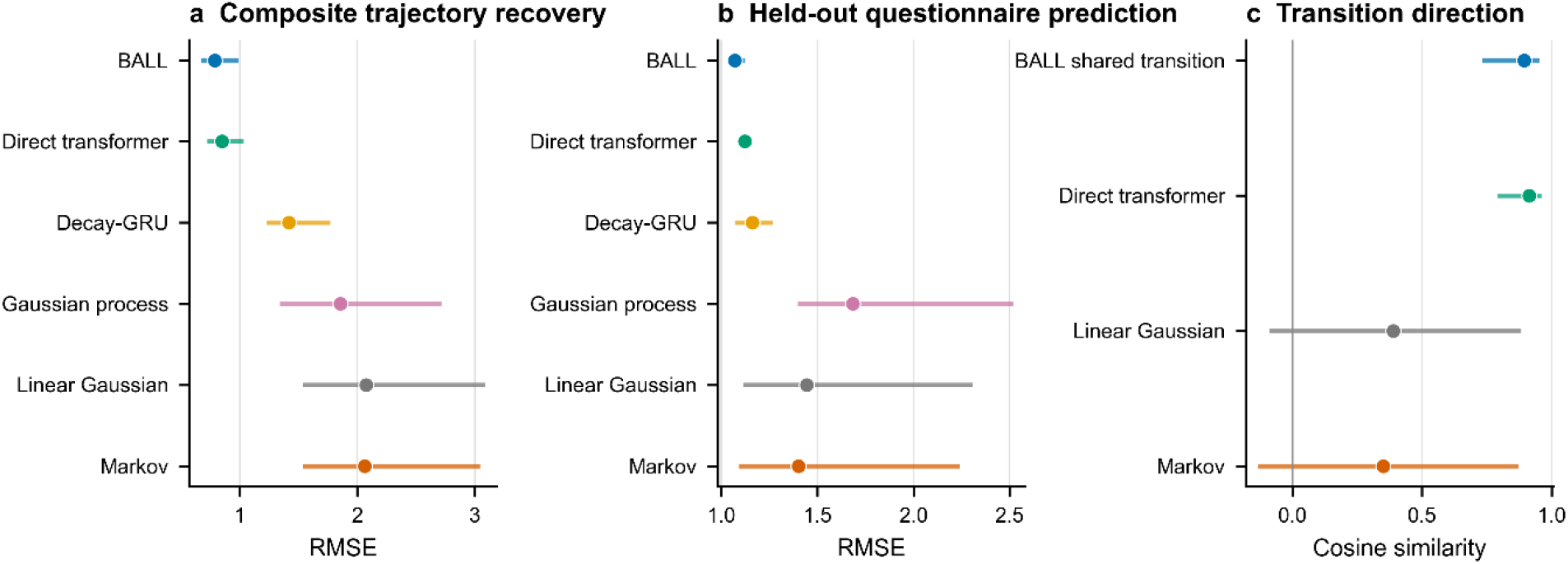
Primary simulation performance for known latent trajectories, withheld questionnaires, and the programmed RDoC coefficient direction. **Notes**. Panels a and b show mean RMSE across 30 simulation runs. Horizontal ranges show the ten setting-specific means and describe variation across settings. Panel a evaluates known composite trajectories, panel b evaluates withheld questionnaires, and panel c evaluates alignment with the programmed RDoC coefficient direction. All trajectory and questionnaire estimates use information available by the current session. Complete training records are used only to fit model parameters and train the teacher. **Abbreviations**. BALL means Bayesian Anchored Latent Learning, and RMSE means root mean squared error.

**Figure 3.**
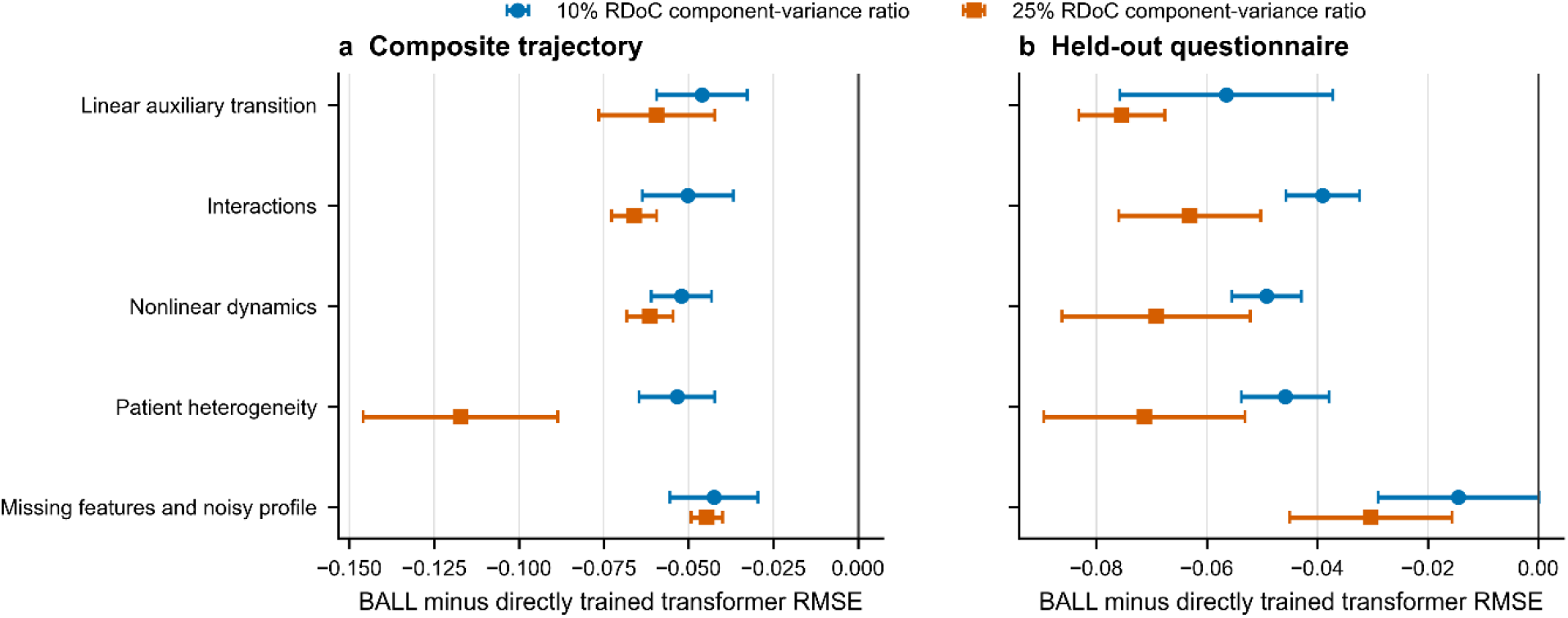
Paired simulation comparison of the BALL student and direct forward training. **Notes**. Each point is the mean of the paired differences across three seeds for one prespecified simulation setting and a programmed RDoC signal strength. Error bars show the Monte Carlo standard error across those three seeds. Values below zero indicate a lower error for the BALL student. The two deployable models use the same forward transformer architecture, observed inputs, ensemble size, transition parameterization, and uncertainty calculation. BALL learns from a retrospective teacher. The direct transformer learns its encoder and transition model jointly from questionnaires using the history available at each session. **Abbreviations**. BALL means Bayesian Anchored Latent Learning, RDoC means Research Domain Criteria, and RMSE means root mean squared error.

To investigate weaker coefficient recovery in the classical models, we tested them in simplified simulations with one active variable and nearly noise-free measurements. We also compared coefficient estimates obtained from true or estimated states and true or noisy RDoC scores to examine sensitivity to input error. The models identified the correct variable and sign in all 12 single-active-variable fits and showed greater sensitivity to trajectory-estimation error at the lower signal strength (Supplementary Table 7).

We tested whether recovery of explicit RDoC coefficients depended on other routes by which the models received RDoC information. We first removed RDoC scores from the encoders and then excluded correlated structured variables, ensuring that this information was entered only through the explicit transition coefficients. Restricting RDoC information to the explicit transition coefficient preserved directional recovery, and removing correlated structured variables further improved it. Estimated coefficients remained smaller than the programmed values (Supplementary Fig. 1 and Supplementary Table 8).

#### Sensitivity across settings and model choices

To assess robustness to different state dynamics and data quality, we compared BALL with direct transformer training across five simulation settings. These included linear relationships, treatment interactions, nonlinear relationships, differences among three patient subtypes, and more missing variables and noisier RDoC scores. Each setting used three random seeds, with RDoC contributions scaled to 10% and 25% of the sum of the component variances in mean symptom change. BALL had lower composite trajectory error than the direct transformer across all five settings, with coefficient recovery weakest in the setting with greater missingness and noisier scores (Supplementary Table 2).

We separately examined whether trajectory recovery depended on the assumed persistence of the state or the weight given to questionnaires during teacher training. Persistence determines how much of the current state carries into the next day, and the questionnaire weight controls the contribution of measurement fit to the teacher’s training objective. Errors changed little across the evaluated daily persistence values, and classical simulation models used a persistence of 0.6 selected using validation data. Teacher questionnaire weights of 20, 40, and 80 produced similar errors, with higher errors at 10 (Supplementary Fig. 2 and Supplementary Tables 9–11).

#### Calibration of uncertainty intervals

We tested whether uncertainty intervals contained known latent states and withheld questionnaire scores at their intended 95% rate. Using three additional samples of 1,000 simulated patients, we compared the original intervals with intervals rescaled using a calibration sample that gave each patient equal weight. Latent-state coverage increased from 92.0% to 94.1%, with an approximately 10% increase in mean width, and withheld-questionnaire coverage increased from 93.9% to 96.1% (Supplementary Fig. 3 and Supplementary Tables 12–13).

#### Negative controls for transition coefficients

We used negative controls to test whether estimated transition coefficients depended on the programmed relationship between RDoC profiles and symptom change. We repeated the analysis after removing that contribution, shuffling profiles across sessions, or replacing them with noise. These changes reduced the overall size (Euclidean norm) of BALL’s coefficients from 0.218 to 0.050 or less (Supplementary Fig. 4 and Supplementary Table 14).

We also tested whether penalizing weak coefficients more strongly improved the identification of the active RDoC domains. We refitted the model with larger penalties for coefficients with smaller initial estimates, using the same patients and inputs. Coefficient direction and active-domain identification were similar to those of the primary unpenalized model (Supplementary Table 15).

#### Alternative questionnaire representation

To test performance with a different representation of reference measurements, we replaced continuous questionnaire totals with a model of ordered response categories for individual items. This comparison evaluated both recovery of the underlying traits and prediction of the observed responses using the same item data and measurement parameters across models. BALL achieved a joint latent-trait RMSE of 0.271, compared with 0.373 for the linear Gaussian filter and 0.371 for the Markov filter. The classical filters achieved lower error in predicting item responses (negative log-likelihood) and expected total scores, indicating that model rankings depended on the evaluated target (Supplementary Table 16).

### Clinical evaluation

#### Prospective prediction of withheld questionnaires

We evaluated BALL in clinical records to test prospective questionnaire prediction under routine measurement patterns and missingness. Patients were separated by clinic so that training and model selection used different clinics from calibration and final testing. The primary dataset included 1,609 patients with major depressive disorder receiving repetitive transcranial magnetic stimulation during 56,939 sessions at 19 Discovery Behavioral Health clinics. Excluding 94 patients whose records crossed clinic partitions left 1,515 patients and 52,493 sessions. Three additional patients had no retained questionnaire under the evaluation schedules, giving a final modeled cohort of 1,512, including 1,168 patients from 13 clinics used for training and validation and 344 from six held-out clinics. We divided held-out patients into calibration and final-test groups (Supplementary Tables 17–18).

To test performance as reference measurements became less frequent, we retained the first questionnaire event for each patient-instrument pair in each 7-, 14-, 21-, or 28-day interval. All methods predicted the remaining events using the same information available at the target session. The schedules retained 32.2%, 19.1%, 14.0%, and 11.2% of recorded questionnaires, respectively. We scaled errors by each instrument’s standard deviation in training patients and gave each patient equal weight in RMSE. Instrument-specific measurement equations converted the estimated depression state into PHQ-9 and BDI-II predictions and the anxiety state into GAD-7 predictions.

We compared BALL with the direct forward transformer to assess the contribution of teacher-based training at each measurement schedule. We also compared both transformers with the linear Gaussian, Markov, Gaussian process, and recurrent models on the same set of withheld questionnaires. BALL’s advantage over direct transformer training increased as questionnaires became sparser. At 21 and 28 days, BALL achieved RMSEs of 0.669 and 0.718, compared with 0.689 and 0.743 for the direct transformer. The paired differences were −0.020 (95% confidence interval, −0.040 to −0.001) and −0.026 (−0.048 to −0.005). Direct training had lower error at seven days (0.577 versus 0.598), and the methods performed similarly at 14 days. BALL had lower estimated RMSE than the linear Gaussian, Markov, Gaussian-process, and recurrent comparators across all four schedules (Fig. 4 and Supplementary Table 19).

**Figure 4.**
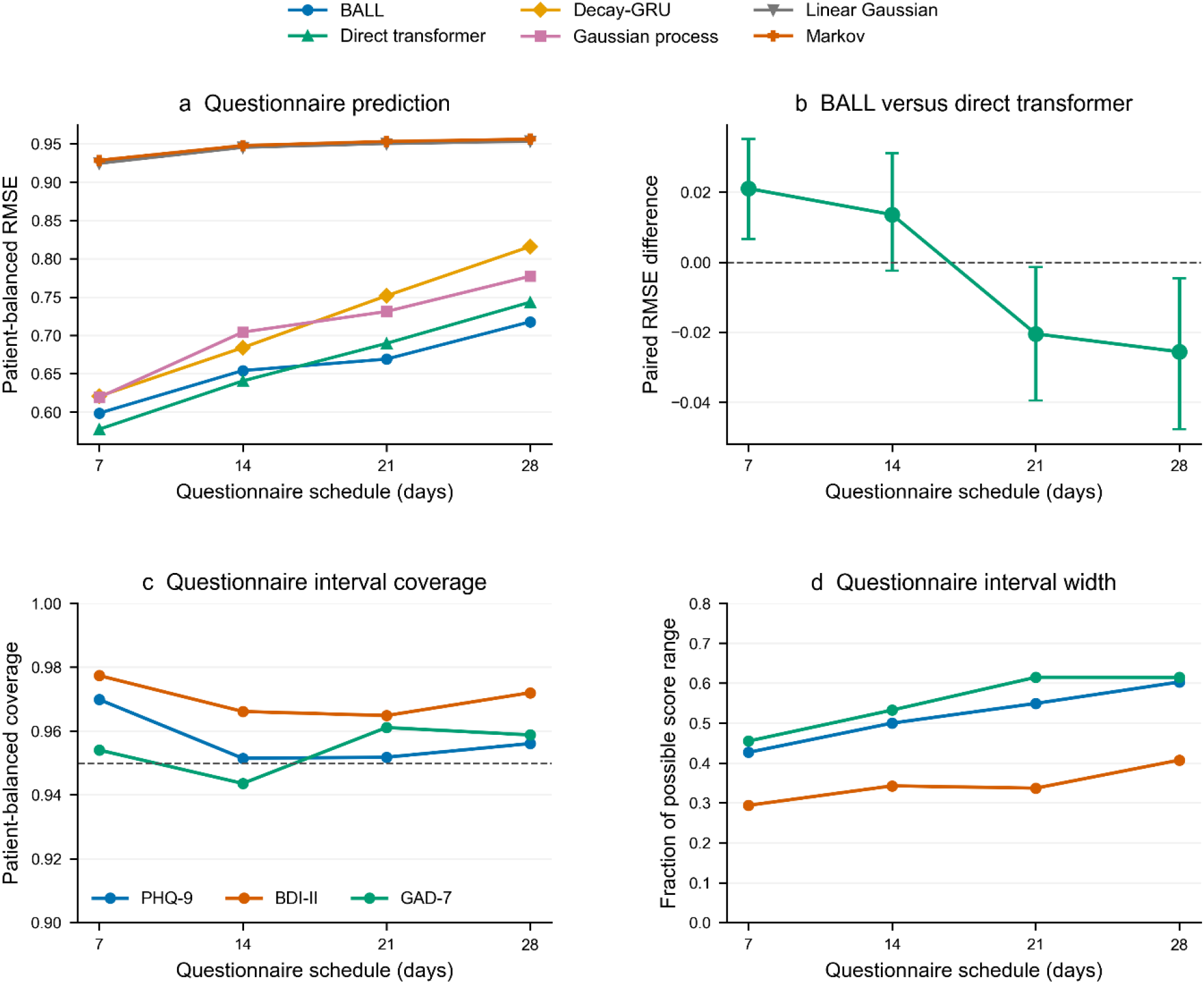
Questionnaire prediction and uncertainty across measurement schedules. **Notes**. Panel a shows questionnaire RMSE with each patient weighted equally for six forward-only methods under 7-, 14-, 21-, and 28-day retention schedules. Panel b shows paired RMSE differences between BALL and the direct transformer, with 95% confidence intervals from 2,000 patient bootstrap samples. Negative values favor BALL. Panel c shows coverage of BALL’s 95% questionnaire prediction intervals by instrument, and panel d shows mean interval width after restricting intervals to valid scores as a fraction of each instrument’s possible score range. Within each schedule, all methods predict the same withheld events. Interval procedures were selected using training and validation patients and calibrated in reserved patients. Decay-GRU denotes the exponential-decay gated recurrent unit. **Abbreviations**. BALL, Bayesian Anchored Latent Learning. BDI-II, Beck Depression Inventory-II. GAD-7, Generalized Anxiety Disorder-7. PHQ-9, Patient Health Questionnaire-9. RMSE, root mean squared error.

#### Information contributing to questionnaire prediction

To identify which records contributed to questionnaire prediction, we refitted BALL with progressively smaller sets of clinical inputs at the 14-day schedule. Separate comparisons changed the weighting of questionnaire events and the student’s training conditions.

Questionnaire history and session timing supplied most of the predictive information at the 14-day schedule. RMSE was 0.649 with questionnaires and timing, 0.648 after adding treatment, and 0.654 with the full feature set. Giving each patient-session equal total questionnaire weight reduced RMSE to 0.642, and teacher-based training configurations produced RMSEs of 0.654– 0.664 (Supplementary Table 20 and Supplementary Fig. 5).

#### Evaluation in a later patient cohort

We tested performance in a later calendar period by fitting models in patients whose records ended before 2025 and evaluating patients first observed in 2025. This analysis compared BALL with the linear Gaussian and Markov filters at the 14-day schedule, using separate later patients for calibration and testing. BALL achieved a questionnaire RMSE of 0.701, compared with 1.038 for the linear Gaussian filter and 1.040 for the Markov filter. Instrument-specific interval coverage was 97.7–99.7% (Supplementary Table 20 and Supplementary Fig. 5).

#### Uncertainty intervals and questionnaire requests

We evaluated the reliability and precision of questionnaire prediction intervals by measuring how often they contained withheld scores and how much of each instrument’s range they spanned. We also assessed coverage among patients whose first retained same-instrument score met the severe-symptom threshold. BALL’s 95% questionnaire prediction intervals achieved instrument-specific coverage of 94.4–97.7% across the four schedules. Mean widths after restricting intervals to valid scores ranged from 29% to 61% of the instrument’s possible score range. Coverage was 95.0–96.0% among events whose first retained same-instrument score met the severe-symptom threshold (Fig. 4 and Supplementary Tables 19–22).

To test whether uncertainty could guide useful additional measurement, we worked through recorded questionnaires in time order and compared predictions under different rules for requesting additional scores. The adaptive rule requested an available questionnaire when the expected reduction in latent-state variance reached 50% and at least seven days had passed since the latest available score from that instrument. Requested scores entered predictions from the next session. At the 28-day schedule, the rule requested 14.6% of the recorded questionnaires withheld under the original schedule and reduced RMSE from 0.718 to 0.658. Collecting every available questionnaire after seven days requested 21.3% and achieved an RMSE of 0.641 (Supplementary Table 23).

#### Clinical interpretation of the estimated trajectories

To assess whether BALL predictions captured clinically interpretable changes, we compared response and remission classifications from predicted and withheld questionnaire scores. Response was defined as a reduction of at least 50% from the first retained same-instrument score, and remission was defined as PHQ-9 and GAD-7 scores of 4 or lower and BDI-II scores of 13 or lower. BALL classified questionnaire-defined response with accuracy of 0.790–0.860 and remission with accuracy of 0.771–0.923 across schedules and instruments (Supplementary Table 19).

We examined whether BALL trajectories maintained differences between patients while varying smoothly within each patient’s treatment course. For each channel, we compared changes between consecutive sessions with differences between patients at the same treatment stage. At the 14-day schedule, between-patient differences exceeded within-patient changes for 98.8% of patients in depression and 99.7% in anxiety, with the same pattern appearing at all four schedules (Supplementary Fig. 6).

#### RDoC content in clinical notes

In a separate analysis, we examined whether earlier changes in RDoC note content predicted subsequent BALL-estimated symptom change across the four measurement schedules and two state channels. Two language models rated domain emphasis across 39,556 narrative fields, and we tested whether adding recent changes in these scores improved prediction beyond current severity, timing, and earlier note content. The raters showed moderate-to-strong agreement, and note-content scores varied more between patients than within patients. Across the eight schedule-by-channel analyses, adding the six note-content changes produced slightly higher prediction error, and none of the associations met the Holm-adjusted significance threshold. These scores, therefore, added negligible predictive information about subsequent trajectories estimated without them (Supplementary Tables 24–26 and Supplementary Fig. 7).

## Discussion

BALL provides a computational framework for following an interpretable, changing state when reference measurements are sparse and indirect records are more frequent. Its contribution is to use complete training histories to train a model that estimates the state prospectively as new observations arrive, with measurement equations preserving the meaning of those estimates over time. Across the evaluated simulations, this approach recovered known trajectories more accurately than the prospective statistical and machine-learning comparators. The psychiatric application then demonstrated how the same design operates with irregular clinical records, with gains over direct transformer training at the 21- and 28-day measurement schedules.

The teacher–student architecture addresses a broader learning problem created by incomplete observation of dynamic systems. A complete training record contains information about earlier states that is unavailable when those states must be estimated prospectively. We use a bidirectional teacher to convert that information into distributions over the evolving state, providing supervision between reference measurements. The forward-only student learns from those distributions and the observed measurements, allowing information learned retrospectively to improve estimation from an accumulating history. The comparison with the same architecture trained directly from measurements supports this combined strategy of teacher initialization, distribution matching, and measurement supervision.

The measurement model makes the recovered state interpretable by linking each reference instrument to a defined state scale. This separation between what the state means and which records inform its evolution provides a route to other scientific applications. For example, intermittent surveys, inspections, or assays could define the target state in environmental monitoring, infrastructure assessment, or longitudinal biology, with more frequent records informing change between measurements. Applying BALL in those settings would require appropriate measurement links and dynamics, which can be specified within the same learning design.

State estimation also provides a basis for deciding when additional information would be useful. BALL combines uncertainty in the latent state with a measurement model, allowing the expected reduction in state uncertainty to guide further data collection. In the analysis of recorded questionnaires, this rule improved subsequent questionnaire prediction at the sparsest schedule and requested fewer measurements than the rule that requested every available questionnaire after the same seven-day minimum. The same principle could support adaptive sampling in other settings where reference measurements are costly to obtain.

The transition model extends the framework from reconstructing a trajectory to examining prespecified relationships with later state change. In the simulations, RDoC variables served as a worked example of inputs with an explicit role in the transition equation. BALL recovered the direction of their programmed contribution and produced smaller coefficients when that contribution was removed or when the inputs were misaligned. More generally, this structure allows investigators to study selected variables alongside flexible neural modeling of the remaining dynamics.

### Limitations

Known latent states were available only in simulation, so empirical validation relied on predicting observed measurements. In the psychiatric application, questionnaires summarized the preceding 14 days, and retained measurements could overlap with the recall period of withheld targets. Questionnaire interval coverage, therefore, evaluates observed-score prediction, and the breadth of the clinical intervals requires further assessment of their usefulness for individual decisions.

Generalization beyond the evaluated setting remains an empirical question. All clinical data came from one provider network, and other populations and scientific domains will differ in what is measured, how observations are collected, and how states change. Independent evaluations should assess those differences as well as performance under shifts in observation frequency and data quality. The adaptive analysis evaluated requests at sessions with recorded questionnaires, so its effects on actual measurement burden and decisions require prospective evaluation.

The usefulness of indirect observations depends on how well they track the state of interest. The clinical RDoC scores measured domain emphasis in notes, with limited within-patient variation and no independent ratings of domain functioning. Their lack of incremental prediction concerns these note-based measurements. In simulation, the transition-coefficient direction was recovered more accurately than its magnitude, and its recovery became less reliable with measurement error or incorrect alignment, highlighting challenges that also arise when using noisy proxies in other domains.

Finally, the current implementation estimates two state channels at recorded observation times, uses fixed persistence, and represents reference measurements through specified observation links. In the clinical application, total questionnaire scores summarized depression and anxiety, leaving other symptom dimensions and item-level differences outside the primary model. Shared persistence and shared RDoC coefficients across simulation channels, together with transitions favoring gradual change, may be restrictive for systems with differing time scales or abrupt changes. These modeling choices should be reassessed when applying the framework to a new system.

BALL brings interpretable state estimation, learning from complete histories, and adaptive measurement into a common computational design. Connecting sparse reference measurements with denser longitudinal records enables the study of how partially observed systems evolve and the identification of when additional observations would be most informative.

## Methods

### Overview

BALL estimates a changing latent state by combining sparse reference measurements with more frequent indirect observations. Measurement equations define the state’s scale and account for measurement uncertainty. A bidirectional transformer teacher uses complete training histories to reconstruct state distributions and learn transitions between them. We train a forward-only transformer student to reproduce these distributions and predict reference measurements, holding measurement equations and teacher-fitted transitions fixed. Causal attention limits each estimate to the available history, allowing retrospective learning to support prospective estimation.^12, 19^

We evaluated the approach in 30 simulated datasets with known depression and anxiety trajectories, varying the relationships that drove state changes, signal strength, missingness, and measurement noise. These simulations allowed us to assess the recovery of the underlying trajectories, the prediction of withheld questionnaires, and the recovery of programmed relationships between indirect observations and subsequent state changes.

We applied BALL to longitudinal psychiatric records, estimating depression and anxiety as two latent-state components, called channels. PHQ-9 and BDI-II measured depression, and GAD-7 measured anxiety,^3–6^ with instrument-specific equations linking scores to the mean state over each questionnaire’s recall window. Clinical history, treatment, and timing informed changes between measurements. We evaluated questionnaire prediction, interval calibration, and additional requests based on uncertainty across increasingly sparse measurement schedules. A separate analysis related the emphasis on six Research Domain Criteria (RDoC) domains in clinical notes to subsequent estimated symptom change. These scores were excluded from the primary clinical model.

The Mass General Brigham Institutional Review Board approved this retrospective study under protocol 2023P001692 and waived informed consent.

### Data structure and timing

We index patients by *i*, sessions by *n*, and the day of the session *n* by *t*_*in*_, with elapsed time *Δt*_*in*_ = *t*_*i,n*+1_ − *t*_*in*_. Each record can contain questionnaires, structured variables *X*_*in*_, indicators of which variables are observed, treatment *A*_*in*_, and an optional six-domain RDoC profile *B*_*in*_. A questionnaire event is one administration of an instrument to a patient. Questionnaires administered at the same session remain separate events.

The encoder is the network that converts available records into a distribution over latent states. The transition model describes how those states change between sessions. Retained questionnaires entered the encoder at the next recorded session, excluding all questionnaires administered at the session being predicted. The encoder received the preceding session’s treatment, and the current treatment was entered into the transition model to estimate change by the next session. Diagnoses became available the next calendar day. In the separate RDoC association analysis, we used only note profiles recorded before the current day. We defined treatment categories in the training clinics and assigned unseen categories to “other.”

### Clinical cohort and questionnaire retention schedules

The cohort comprised 1,609 patients and 56,939 repetitive transcranial magnetic stimulation (rTMS) sessions at 19 clinics from 2 January 2023 to 27 June 2025. All three questionnaires used a 14-day recall.^3–6^ We assigned 10 clinics to training, three to validation for choosing model settings, and six to held-out evaluation. Excluding 94 patients attending clinics in different partitions left 1,170 patients across the training and validation clinics and 345 held-out patients. We divided the held-out patients into a calibration group for the final interval correction and a separate test group for performance evaluation.

To study less frequent measurement, we retained the first questionnaire event in each 7-, 14-, 21-, or 28-day interval for each patient and instrument. Retained events were available as model inputs, and the remaining events were withheld as prediction targets. Retention changed input frequency but preserved each instrument’s 14-day recall window.

We included earlier retained scores whose recall windows overlapped the target’s recall window because these scores were available when making the prediction.

We preserved questionnaires administered together as separate records for each patient, session, and instrument, rejecting duplicates, merged instruments, and events outside the designated input and evaluation sets. Two patients from the training and validation clinics and one held-out patient lacked retained questionnaires, leaving 1,168 patients across the training and validation clinics and 344 held-out patients for modeling. The modeled held-out cohort comprised 171 calibration patients and 173 final-test patients. Patients contributing withheld events numbered 168 in calibration at every schedule and 170, 170, 171, and 171 in final testing at 7, 14, 21, and 28 days, respectively.

### Empirical structured features and temporal ordering

The 14 structured features were treatment-session number, cumulative non-depression comorbidity count, and prior indicators for depression, anxiety, post-traumatic stress, bipolar, obsessive-compulsive, attention-deficit/hyperactivity, psychotic-spectrum, substance-use, eating, personality, autism-spectrum, and sleep disorders.

One indicator recorded whether earlier diagnosis information was available for the comorbidity and diagnosis variables. We retained features on their recorded scales, including raw counts for session number and elapsed days, and followed the timing rules above. We checked that inputs came from permitted source fields and fitted comparator preprocessing only in training patients.

### Latent states and transitions

We jointly modeled two related latent channels, depression 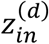 and anxiety 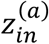, consistent with dimensional frameworks of psychopathology.^23–24^, ^36–37^ Joint modeling allowed information about either symptom channel to inform the other, with each questionnaire retaining its instrument-specific relationship with the relevant channel.

Composite simulation metrics were computed by averaging the two estimated channels, as given by Equation 1.

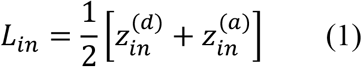

Here, *L*_*in*_ is the composite severity for the patient *i* at session *n*, calculated from depression 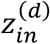 and anxiety 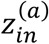. We used *L*_*in*_ only for composite metrics.

The transition model combines the persistence of the current state with expected change based on observed inputs and unexplained variation. For channel *c*, Equations 2 and 3 accumulate these components over the days between sessions *n* and *n* + 1.

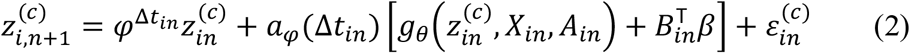

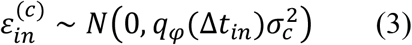

Here, *z* is severity, *c* identifies the channel, and *φ* is daily persistence, the fraction of the previous day’s state carried forward. Both channels shared a fixed persistence of 0.30 in primary BALL simulations, 0.60 in classical simulation comparators, and 0.90 in empirical BALL fits.

The neural network *g*_*θ*_, with parameters *θ*, estimates daily change from severity, structured variables, and treatment, with optional RDoC scores *B* contributing through coefficients *β* shared across simulation channels. The residual *ε* represents unexplained change and is normally distributed, with *q* accumulating its variance across the gap and *σ* specifying the channel-specific scale.

Equation 4 accumulates daily change and variance over an observed gap.

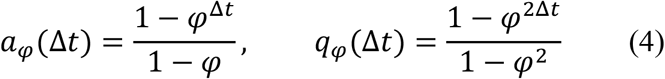

The factors *a*_*φ*_(*Δt*) and *q*_*φ*_(*Δt*) accumulate expected change and variance, respectively, and both equal *Δt* when *φ* = 1. Simulations used *Δt*_*in*_ = 1, and clinical transitions used observed calendar gaps. We fitted transitions only between sessions separated by a positive time gap, but included all sessions in the encoder and questionnaire likelihoods.

### Questionnaire measurement model

To put instruments on a comparable scale, we normalized questionnaire totals by subtracting and dividing by the midpoint of each instrument’s valid range, then reversed this transformation for reporting. For the normalized score *Y*_*ijm*_, *m* identifies the instrument, *i* the patient, and *j* the administration. Because questionnaires summarize a recall period, Equations 5 and 6 link the score to the mean state across recall-window sessions *W*_*ijm*_, with *c*(*m*) identifying the instrument’s depression or anxiety channel.

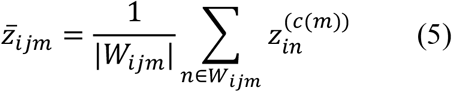

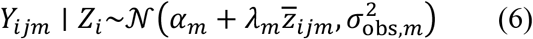

In Equation 6, *Z*_*i*_ is the patient’s state sequence, and the intercept *α*_*m*_ and loading (slope) *λ*_*m*_ convert its recall-window mean 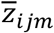 to the instrument’s expected score. The measurement-error standard deviation *σ*_obs,*m*_ describes variation around that expectation. We gave recall-window sessions equal weight and included each questionnaire administration separately, linking PHQ-9 and BDI-II to depression and GAD-7 to anxiety.

PHQ-9 defined the depression scale, and GAD-7 defined the anxiety scale through intercepts fixed at zero and loadings fixed at one. BDI-II measured depression through an estimated intercept and positive loading, with a separately estimated measurement-error standard deviation for each instrument.

We calibrated these links and measurement variances using all questionnaires from the 10 training clinics before fitting trajectories or applying retention schedules. We used temporary state-space models with Gaussian errors to jointly calibrate PHQ-9 and BDI-II and estimate GAD-7 measurement variance. These processes had an initial-state variance of one and the following transitions.

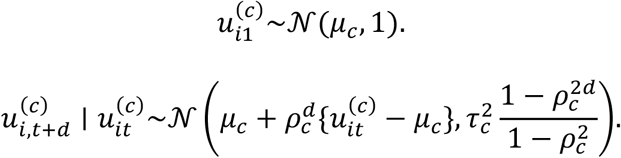

The calibration state *u* has mean *µ*, daily persistence *ρ*, and standard deviation of unexplained daily change *τ*. The second equation describes its change over *d* days.

We estimated the instrument links by averaging over possible trajectories under these Gaussian processes, accounting for uncertainty in the temporary trajectories. This gave the marginal likelihood below, which we maximized jointly over the instrument and temporary-process parameters.

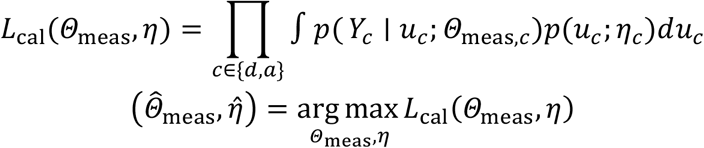

Here, *Y*_*c*_ contains questionnaires for the channel *c, u*_*c*_ its temporary calibration trajectories, and *Θ*_meas,*c*_ its instrument parameters. The temporary-process parameters *η*_*c*_ comprise the process mean, persistence, and standard deviation of unexplained daily change. We evaluated the integrated likelihood separately for depression and anxiety using Kalman updates, which successively predict and incorporate each questionnaire.

We maximized this marginal likelihood using bounded limited-memory Broyden–Fletcher– Goldfarb–Shanno optimization, initializing the means, persistence, and standard deviations at 0, 0.5, and 0.25, and the BDI-II intercept and loading at 0 and 1, respectively. Bounds were −3 to 3 for means, −4.6 to 4.6 for the log-odds of persistence, −5 to 1 for log standard deviations, and −2 to 2 for the BDI-II intercept and log loading. We allowed 160 iterations for depression and 120 for anxiety, with a tolerance of 0.0000000001.

After calibration, we transferred the instrument intercepts, loadings, and measurement variances to all trajectory models and held them fixed. At the 14-day schedule, we also gave each patient-session equal total questionnaire weight to assess the influence of administering both depression instruments together.

### Retrospective teacher and forward-only student

The two transformer encoders used attention to combine information across sessions. The teacher could attend to the complete training record, whereas a causal attention mask blocked the student from attending to later sessions. Both returned distributions over the same two latent channels, and only the student supplied prospective estimates.

At each session, both transformers represented depression and anxiety by a joint Gaussian distribution, with Equation 7 defining the teacher’s distribution.^12, 38–39^

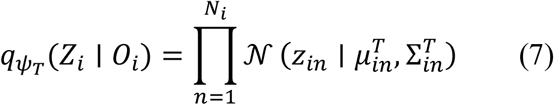

For patient *i, O*_*i*_ is the complete record of *N*_*i*_ sessions and *Z*_*i*_ the state sequence. The teacher distribution 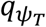 has parameters *ψ*_*T*_, and at the session *n*, 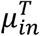 contains the channel means and 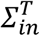 their two-by-two covariance. In the primary fits, the two state components represent depression and anxiety, with covariance between the channels fixed at zero within each independently initialized fit.

### Training the retrospective teacher

We fitted the teacher encoder and transition model jointly to the observed questionnaires, keeping instrument links fixed. In simulations, we also trained them to reconstruct observed structured variables from the latent states. Six additional Gaussian latent variables had their own transitions and a penalty drawing them toward zero. These auxiliary variables were excluded from the reported depression and anxiety states. Equation 8 combines these objectives.^38–39^

Each channel’s initial state follows 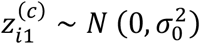, with learned initial-state scale *σ*_0_.

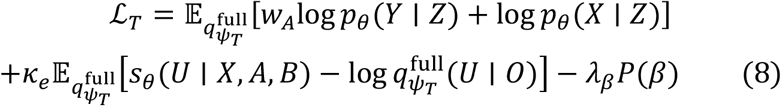

The complete encoder distribution 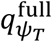 represents *U* = (*α, Z*), comprising six auxiliary trajectories *α* and the two reported trajectories *Z*. We averaged the objective under this distribution. Its likelihood terms assess agreement with questionnaires *Y*, weighted by *w*_*A*_, and simulated structured variables *X*. The score *s*_*θ*_ combines Gaussian log densities for the initial states and transitions of *U* across positive time gaps with zero-centered Laplace log densities for *α* at fixed rate 2. The Laplace terms draw the auxiliary variables toward zero. Subtracting the encoder log density gives a regularization term that balances agreement with the state model against overly concentrated state distributions. This variational regularization has weight *κ*_*e*_ at epoch *e*, where an epoch is one pass through the training patients. Parameters *ψ*_*T*_ and *θ* govern the teacher encoder and generative model, with instrument links fixed. Penalty *P*(*β*) acts on the transition coefficient *β* with strength *λ*_*β*_.

We set the questionnaire weights to 40 in the simulation and to 10 in clinical fitting, where clinical variables were inputs to the encoder and transition model. The primary coefficient analysis used *λ*_*β*_ equal to zero, with adaptive shrinkage assessed separately.

The first phase, called questionnaire warm-up, combined questionnaire fit and transitions to establish the direction and scale of each latent channel before the full objective was applied. The second phase added prediction of structured variables from inferred states in simulation, with *κ*_*e*_ increasing from 0 to 1 during each phase.

### Training the forward-only student

After teacher fitting, we used distillation to train the student to approximate the teacher’s session-level state distribution from the history available at that session.^19^ We initialized each student from its paired teacher’s input and session-position representations, transformer layers, normalization parameters, and layers that produce the state distributions, together with a copy of the learned numerical representation of treatment, called the treatment embedding.

During student training, we held the teacher network, its treatment embedding, its fitted transitions, and the questionnaire links fixed, and updated only the student encoder and its copied treatment embedding. This preserved the fitted symptom dynamics and questionnaire scale as the encoder adapted to information available at each session.

Student training balanced agreement with the teacher’s state distributions against prediction of observed questionnaires. Equation 9 uses Kullback–Leibler (KL) divergence to measure distribution disagreement across sessions. Negative log-likelihood penalizes low predicted likelihood for observed questionnaire scores and is averaged across administrations.

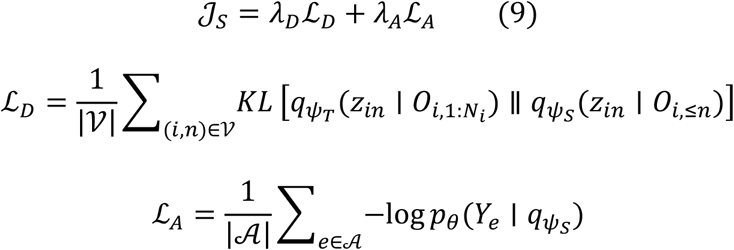

The total loss *J*_*S*_ combines distribution loss *L*_*D*_ and questionnaire loss *L*_*A*_ with weights *λ*_*D*_ and *λ*_*A*_. The set *V* contains |*V*| training sessions.

For patient *i* and session *n, z*_*in*_ is the joint state, represented by teacher and student distributions 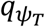 and 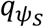 with parameters *ψ*_*T*_ and *ψ*_*S*_. The teacher uses all *N*_*i*_ sessions in 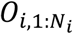, and the student uses information through *n* in *O*_*i*,≤*n*_. Divergence is averaged over *V*.

Teacher-to-student divergence penalizes low student probability where the teacher assigns substantial probability. This direction can accommodate a broader student distribution when the prospective record contains less information than the complete record used by the teacher. We used the full two-by-two covariance matrices, adding 0.000001 to their diagonals to stabilize the calculation.

The set *A* contains |*A*| questionnaire administrations. For event *e, Y*_*e*_ is the score and *p*_*θ*_ its probability under the fixed measurement parameters *θ*. The questionnaire loss *L*_*A*_ averages their negative log-likelihoods.

For Gaussian questionnaires, we averaged state means across the recall window and combined state variances using squared recall weights, then added measurement variance. In the ordinal sensitivity below, we integrated item-category probabilities over the student’s trait distribution using 15-point Gauss–Hermite quadrature, a weighted numerical average over possible trait values. Item log-likelihoods were averaged within administrations.

Using initial training and validation, we fixed the distribution-matching weight to 1 and the questionnaire weights to 5 in simulation and to 2 clinically, using patients and seeds separate from the definitive evaluation.

### Questionnaire prediction intervals

A questionnaire prediction interval gives a plausible range of observed scores, accounting for state and measurement uncertainty. We constructed these intervals in three stages.^30–35^ First, we calculated uncertainty from latent states and the final prediction layer over each recall window, combined predictions across five independently trained models, and added observation variance. Second, we estimated percentiles of prediction errors given available history in training and validation patients. Third, we used reserved calibration patients to correct the lower and upper interval limits.

We fitted five independently initialized teacher–student pairs and combined their predictions as an ensemble. For patient *i*, session *n*, and member *r*, Equations 10 and 11 combine means 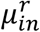 and variances 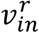.

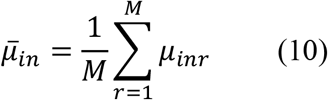

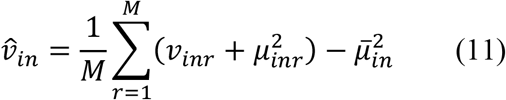

With *M* = 5 members, *µ*_*in*_ is the ensemble mean and *V*_*in*_ combines average within-member variance with variance between member means. Thus uncertainty increases when individual fits are uncertain or independently fitted models disagree.

We also accounted for uncertainty in the fitted weights of the final prediction layer. A diagonal Laplace approximation represented these weights by a Gaussian distribution around their fitted values, with separate variances for each weight, as shown in Equation 12.^31–32^

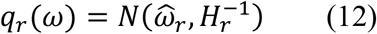

For member *r, q*_*r*_(*ω*) is centered on fitted weights *ω*^*r*^ with covariance 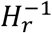, and diagonal precision *H*_*r*_ equals one plus the sum of squared values from the final hidden layer in training patients. Precision is the inverse of variance, so greater precision reduces uncertainty about a fitted weight. We used a multiplier of one for this weight uncertainty before combining ensemble members.

For each model, we averaged states through the target session over the empirical 14-day recall window before calculating disagreement between models. We combined session-specific state variances using squared recall weights and applied final-layer weight uncertainty to the average hidden-layer representation. Because sessions shared the same output intercept, its variance contributed once to the recall-window mean. Later sessions, including those on the same day, were excluded.

### Adapting and calibrating prediction intervals

For each instrument and retention schedule, we used histogram gradient boosting, a tree-based regression method, to estimate conditional quantiles of normalized questionnaire residuals. These quantiles are percentiles of observed minus predicted scores that vary with available history.

Inputs were the predicted mean and standard deviation, elapsed days, number of sessions in the recall window, time since the latest questionnaire, numbers of earlier questionnaires overall and by instrument and channel, and latest and first scores from the same instrument. More detailed histories added time since the latest administration of the same instrument, the latest change in score and that change divided by elapsed days, and the mean, standard deviation, range, and daily slope of the last three scores. They also included elapsed calendar days between the first and latest questionnaires from the same instrument, numbers of questionnaires from the same instrument in the preceding 14 and 28 days, and the fraction of overlap between current and latest recall windows. All inputs were available when predicting the target score, and quantile estimation left latent-state estimates unchanged.

We converted residual quantiles to lower and upper limits on the normalized questionnaire scale by adding the BALL-predicted mean. After sorting and applying the valid score bounds, these endpoints supplied the lower and upper functions in Equations 13–14.

We compared four interval procedures, with the first three estimating the 2.5th and 97.5th percentiles. The first used prediction errors from patients used to fit BALL. The second used errors from patients excluded from that fit, and the third added the more detailed histories described above. The fourth used the third procedure’s errors and histories, selecting the narrowest of five intervals with lower percentiles 0.5, 1.5, 2.5, 3.5, and 4.5 and upper percentiles 95 percentage points higher. Before calibration, we ordered the quantiles from lower to upper and restricted them to valid scores. To obtain these errors, we divided training and validation patients into two groups with representation from each clinic. For each group and schedule, we fitted five BALL pairs using 60 warm-up, 120 teacher, and 120 student epochs, with measurement links fitted only to the patients used to train that model. Each patient’s residual came from the model trained in the other group, with reserved calibration and test patients excluded throughout.

Within the 10 training clinics, we randomly assigned 80% of patients to interval fitting and 20% to initial interval calibration. In the three validation clinics, we compared four procedures and three configurations using an interval score that gives patients equal weight and combines width with penalties for scores outside the interval. We used predictions for patients excluded from BALL fitting to fit, calibrate, and compare the interval procedures. Regression trees had up to 7 terminal groups (leaves) with 100 or 200 iterations, or 15 leaves with 100 iterations. All used learning rate 0.05, at least 100 observations per leaf, and quadratic penalty 1. We refitted the selected procedure in all 13 training and validation clinics, using the prediction errors specified by that procedure and giving each patient equal weight within the instrument. We used reserved calibration patients for the final correction to the interval limits, and temporal evaluation repeated selection within pre-2025 data. Alternative interval procedures remained sensitivities (Supplementary Tables 19–20).^35^

We then used reserved calibration patients to correct the interval endpoints. Equation 13 measures the larger signed deviation beyond the fitted quantiles, weighting each patient equally within the instrument. Scores between the lower and upper quantiles have negative calibration residuals.

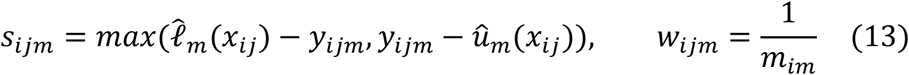

Patient, event, and instrument are indexed by *i, j*, and *m*. The observed normalized score *y*_*ijm*_ is used to compute the calibration residual. Prior information *x*_*ij*_ alone determines the predicted quantiles 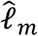 and *û*_*m*_. The residual is *s*_*ijm*_, and weight *w*_*ijm*_ gives each patient equal total influence across *m*_*im*_ questionnaires included in calibration. The correction 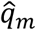is found by sorting residuals from smallest to largest and accumulating their patient weights. We select the first residual whose cumulative weight reaches 95% of one plus the number of calibration patients, rounded upward. We fitted this correction separately for each instrument and schedule using reserved calibration patients.

Equation 14 applies this correction, restricts the interval to valid scores, and permits conversion back to questionnaire points.

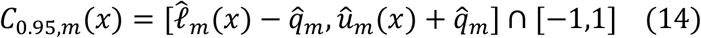

For a new history *x, C*_0.95,*m*_ is the calibrated interval for instrument *m*. Coverage is the fraction of observed scores falling inside their intervals, with each patient weighted equally. We assessed coverage and width by instrument, earlier questionnaire availability, elapsed time, treatment week, demographics, and clinic. High-severity groups were defined before prediction using the first retained score from the same instrument, with cutoffs PHQ-9 ≥20, BDI-II ≥29, and GAD-7 ≥15 (Supplementary Table 22). We excluded events lacking an earlier score from the same instrument from these groups.

### Comparison models

We compared BALL with two classical state-space models, a direct transformer, a Gaussian-process filter, and a smaller recurrent network. We fitted parameters in training data, and every trajectory and questionnaire comparison used estimates based only on information available at that session. For auxiliary profiles, we used the most recent earlier observation, or zero before the first observation. Questionnaire-history groups likewise used only earlier observations.

### Shared information across models

All models used the same patient partitions, retained questionnaires, observed inputs, session times, and withheld evaluation targets. Predictions followed the timing rules above. We fitted preprocessing in training data and chose model settings using validation data. Simulation inputs included structured variables, their availability indicators, treatment histories, and RDoC profiles. Training likelihoods assigned questionnaires to their administration sessions, and the classical models’ regressions for estimating states from structured variables included both feature values and availability indicators.

Clinical models shared 14 structured features, availability indicators, previous treatment, and elapsed time, and statistical filters used earlier questionnaires without revising previously issued states. We verified that inputs and targets matched across models and stopped the analysis if they differed.

### Linear Gaussian state-space model

The linear Gaussian benchmark linked continuous depression and anxiety states to observed measurements through linear equations with Gaussian errors.^9–11^ Simulation channels had separate intercepts, noise scales, questionnaire links, and regressions estimating states from structured variables, with shared persistence and RDoC coefficients. We alternated state estimation with regression for transitions, using a ridge penalty to limit large coefficients.^40^

Equation 15 describes how the estimated state changes between two observed sessions.

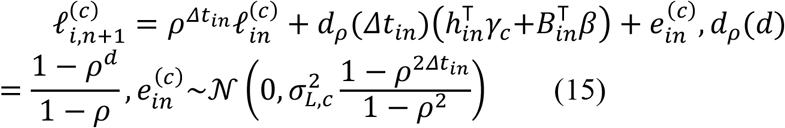

In Equation 15, 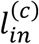 is the channel state and *ρ* its daily persistence. The vector *h*_*in*_ contains the other transition predictors, including an intercept and treatment history, with coefficients *γ*_*c*_. The factor *d*_*ρ*_ scales change over the elapsed gap, and residual 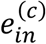 represents unexplained change with daily error standard deviation *σ*_*L,c*_. We linked the questionnaires using Equation 6 and replaced missing structured-feature values with means calculated from training patients. We compared persistence values of 0.1, 0.3, 0.6, and 0.9 using validation data and chose 0.6 for both classical simulation models.

For the clinical data, we used separate random-walk models for depression and anxiety, adding baseline drift and treatment contributions over the elapsed days. Linear regression on fourteen structured features provided another estimate of each state, with missing values replaced by means calculated from training patients according to availability indicators. We fitted parameters using complete training records, then held them fixed when estimating states prospectively.

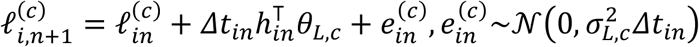

Here, *θ*_*L,c*_ contains the baseline and treatment coefficients. The equation scales their contribution and residual variance by elapsed days. We used ridge penalties of 1 for structured-feature regression and 10 for transitions, an initial-state standard deviation of 5, a daily transition standard deviation of 0.75, and five alternating updates.

### Markov pattern-mixture model

The Markov pattern-mixture model allowed dynamics to depend on questionnaire availability.^28^ Two cutoffs fixed using training data divided sessions into three groups according to the number of retained questionnaires recorded earlier. Simulation groups had separate intercepts, with channel-specific dynamics, persistence of 0.6, and a shared RDoC coefficient vector.

Equations 16 and 17 describe the simulation transition.

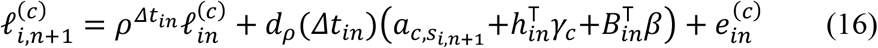

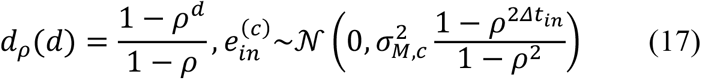

Symbols follow Equation 15. The intercept 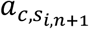 depends on channel *c* and patient *i*’s group *s*_*i,n*+1_ at the later session, and *σ*_*M,c*_ is the standard deviation of unexplained change. Primary Gaussian fitting used 3–25 alternating updates, stopping when relative changes in states, coefficients, and noise scales were at most 0.0001. Classical control and ordinal sensitivities allowed 300 updates with tolerance 0.002.

Clinical group boundaries were fixed based on prior questionnaire counts across sessions for the 1,168 patients used for final training. Each channel and group had a rate governing persistence of the earlier state and accumulation of baseline and treatment contributions, with variance proportional to elapsed days and the same regression for estimating states from structured variables as the linear Gaussian model.

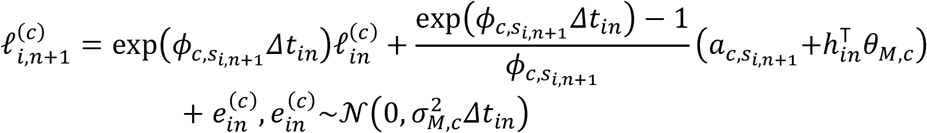

Here, 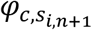 is the rate governing persistence of the earlier state for the channel *c* and patient *i*’s group at the later session. The group intercept and treatment coefficients *θ*_*M,c*_ contribute through the elapsed-time factor shown above. We used four alternating updates, a ridge penalty of 10, and rates bounded between −0.95 and 0.05.

### Direct forward transformer

To test learning without a retrospective teacher, we used a direct forward transformer that shared BALL’s forward architecture, inputs, ensemble size, transition model, and uncertainty calculation. We jointly trained its randomly initialized encoder and transition model against questionnaires using fixed instrument links.

Equation 18 describes the distribution produced by this transformer.

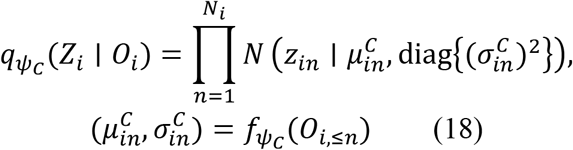

For patient *i, Z*_*i*_ is the state sequence and *O*_*i*,≤*n*_ the record through the session *n*. Encoder 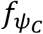 produces mean 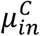 and standard deviation 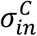, defining 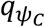. Training used the teacher’s questionnaire warm-up and subsequent schedule, with a separate simulation comparison under the stopping rules described below.

### Prospective Gaussian-process filter

The Gaussian-process comparator estimated states by treating sessions with similar times and clinical inputs as more strongly related to one another.^29^ Separate processes for depression and anxiety combined smooth and rapid temporal components with similarity in structured features and treatment.

Equations 19 and 20 define these relationships.

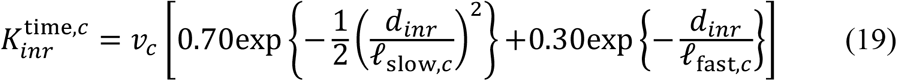

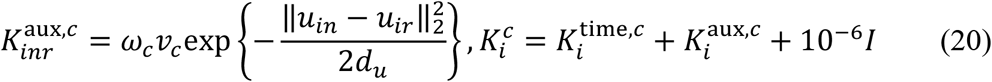

For patient *i*, sessions *n* and *r*, and channel *c, d*_*inr*_ is the absolute difference in calendar days and *V*_*c*_ the channel variance. Time scales *l*_*slow,c*_ and *l*_*fast,c*_ control how quickly relationships between sessions weaken as their time gap grows, with the rapid component using *d*_*inr*_/*l*_*fast,c*_. Vectors *u*_*in*_ and *u*_*ir*_ contain features, availability indicators, and preceding treatment, and have dimension *d*_*u*_. The weight *ω*_*c*_ controls clinical similarity. For numerical stability, we added 0.000001 times the identity matrix *I* to the covariance matrix.

We fitted scaling, mean functions, and channel variances in the 10 training clinics. We then used the three validation clinics to select slow time scales of 14, 28, or 56 days and fast time scales of 1, 3, or 7 days based on questionnaire negative log-likelihood, keeping the clinical similarity weight fixed at 0.15. After selection, we refitted the model in all 13 training and validation clinics. In the simulation, we selected from the same candidate time scales separately for each channel, using the questionnaire marginal likelihood in training patients.

At each session, the Gaussian-process estimate used questionnaires recorded during strictly earlier sessions, as expressed in Equation 21.

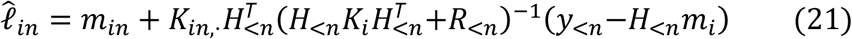

Here, *m*_*in*_ is the mean state predicted from clinical inputs for the current session, *m*_*i*_ collects these means across sessions, and *K*_*ic*_ describes covariance between latent states at different sessions. Equation 21 omits channel superscripts for readability. Matrix *H*_<*n*_ maps earlier states to questionnaire recall windows and scales, *y*_<*n*_ contains earlier scores after subtracting their instrument intercepts, and *R*_<*n*_ their measurement variances. Earlier prediction errors update the current estimate.

### Exponential-decay gated recurrent unit

The exponential-decay gated recurrent unit (GRU) served as a smaller neural comparator that summarized previous sessions into a running numerical representation.^27^ It reduced the contribution of that earlier representation as the time gap grew, then combined it with current inputs.

Equations 22 and 23 describe these two steps.

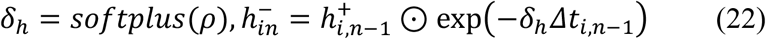

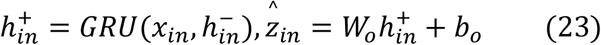

Learned positive rates *δ*_*h*_ reduce the previous representation 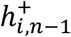 to 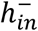 across gap *Δt*_*i,n*−1_. The recurrent unit combines 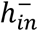 with input *x*_*in*_ to produce 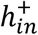, from which output weights *W*_*o*_ and intercept *b*_*o*_ yield the two states 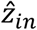. Softplus maps unrestricted rate parameters *ρ* to positive values, and *⊙* denotes multiplication of corresponding vector entries.

Equation 24 combines the questionnaire squared error with a penalty for abrupt state changes over short time gaps.

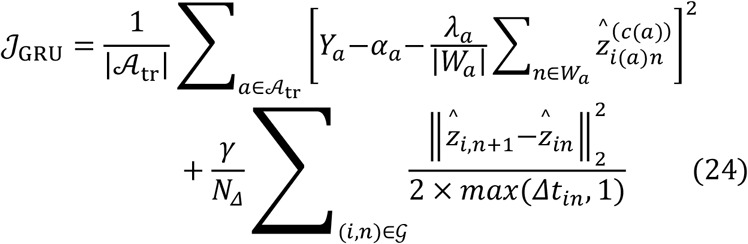

The questionnaires used for training form the set *A*_tr_. For questionnaire *a, Y*_*a*_ is its score, *W*_*a*_ its recall window, *λ*_*a*_ its loading, and *c*(*a*) its channel, with intercept *α*_*a*_ and patient index *i*(*a*). The *N*_*Δ*_ adjacent pairs in *G* contribute a penalty for abrupt changes over short time gaps weighted by *γ*. We set *γ* to 0.01 in simulation and 0.02 clinically, using one initialization per simulation replicate and five independent initializations clinically.

### Model architecture, tuning, and optimization

The teacher, student, and direct transformer used three layers with 96 internal units and four attention heads, which combine information across sessions in parallel. During training, dropout randomly removed internal activations with probability 0.10, treatments were represented by 8 learned numbers, and the transition network had 32 units per hidden layer. Adam learning rates were 0.002 during questionnaire warm-up and 0.001 afterward,^41^ with gradients rescaled when their norm exceeded 5 to limit large updates.^42^

Primary simulations used 120 warm-up, 300 teacher, and 300 student epochs, a gradual increase in the variational regularization weight over 80 epochs, and five ensemble members. Warm-up used the full training cohort, followed by batches of 32 patients for teacher and student training. The recurrent comparator was trained for 300 epochs.

Clinical fits used 60 warm-up, 120 teacher, and 120 student epochs, a gradual increase in the same weight over 60 epochs, and five members, with the full training cohort used for warm-up and batches of 32 patients thereafter.

To choose model settings at the 14-day schedule, we trained in 10 clinics and selected settings in 3 clinics using questionnaire error averaged equally across patients. We tested all combinations of persistence (0.1, 0.3, 0.6, and 0.9) and questionnaire weight during teacher training (10, 20, 40, and 80), then student questionnaire weights (0.5, 1, 2, 5, and 10). Among student weights whose squared error was within one standard error of the minimum, we selected the weight with the lowest questionnaire negative log-likelihood. The selected persistence, teacher weight, and student weight were 0.9, 10, and 2.

We carried these selections forward, refitting the primary model in all 13 training and validation clinics and fixing these settings across schedules, held-out clinics, temporal evaluation, and focused analyses. The recurrent model’s smoothness penalty, statistical-model penalties, and Markov group cutoffs were fixed before held-out evaluation. Calibration and test patients had no role in fitting shared model parameters or selecting settings.

### Simulation design

Simulations allowed us to evaluate the recovery of known latent states and prediction of noisy questionnaire scores. Each primary dataset contained 150 patients followed for 84 days, two latent channels, 24 partly observed variables, 12 treatment types, six RDoC scores, and sparse questionnaires. Treatments lasted 10–15 days, and we assigned 60% of patients to training, 10% to model selection, 10% to interval calibration, and 20% to testing.

Six groups of four structured variables reflected current severity, severity during the preceding seven days, the underlying RDoC process, rapidly changing residual symptoms, recent and cumulative treatment, and unrelated noise, with primary missingness of 30%. Treatment assignment depended weakly on recent severity.

We scheduled both simulated questionnaires weekly, using a 14-day recall, an observation probability of 0.90, a loading of 1, and a measurement-error standard deviation of 0.95 for depression. Anxiety used a seven-day recall, probability 0.80, loading 0.75, and standard deviation 0.85, with all methods sharing these fixed measurement parameters.

Three of six RDoC dimensions affected subsequent change in both channels. Their coefficient vector *β*^*\**^ was rescaled to length one (Euclidean norm), and their contribution was scaled to 10% or 25% of the sum of the component variances in Equation 25.

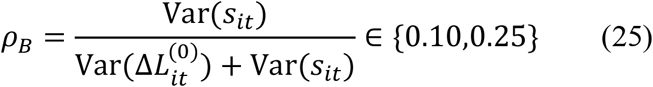

Here, *ρ*_*B*_ is the designated component-variance ratio, and 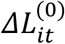 is a change in the mean of the two latent channels before adding the RDoC contribution. We computed within-patient variances and summed them across patients in the heterogeneous setting, using the sum of the component variances as the denominator. The weighted sum *r*_*it*_ of domain scores using coefficients *β*^*\**^ has patient mean 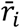 for patient *i*, and constant *κ* scales the centered contribution *s*_*it*_ to the designated variance share.

Five settings varied the RDoC contribution. It entered directly into the linear setting and was multiplied by 1 + 1.5*R*_*it*_ for the recent-treatment indicator *R*_*it*_ in the interaction setting. The nonlinear setting added 0.75 *tanh*(*C*_*it*_)^*T*^*β*^*\**^ to the linear projection, where *C*_*it*_ is the underlying six-domain profile before measurement noise. The hyperbolic tangent is applied to each coordinate before projection, and both projections are centered within the patient. In the heterogeneous setting, we applied subtype multipliers of 0.5, 1.0, and 1.5 after within-patient normalization. Then we used a common rescaling to preserve the pooled variance ratio while allowing differing ratios across subtypes.

The fifth setting had at least 55% structured-feature missingness, missingness dependent on underlying severity with strength 0.7, and a baseline RDoC measurement-error standard deviation of 0.40. To increase error when profiles were sparse, we multiplied this standard deviation by one plus 1.5 times the fraction of days without a profile over the most recent seven days. Five settings, two signal strengths, and seeds 2027, 2028, and 2029 produced 30 datasets.

In a separate 1,000-patient uncertainty experiment over 84 days, we used three seeds and five ensemble members to assess interval calibration separately for latent states and questionnaire outcomes. Calibration multiplied model standard deviations by a factor estimated with equal patient weights. We also grouped uncalibrated latent-state intervals by feature missingness and distance to the nearest observed questionnaire. Questionnaire intervals that adapt to earlier history were fitted only in the clinical analysis.

### Simulation evaluation

We evaluated latent-state recovery and questionnaire prediction separately using root mean squared error (RMSE). For explicit RDoC coefficients, we assessed their direction using cosine similarity and identified the three active dimensions using the F1 score, which combines precision and recall.

Equation 26 gives the cosine similarity between an estimated coefficient vector and the programmed vector.

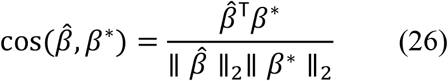

Equation 26 normalizes the inner product of the estimated *β* and programmed *β*^*\**^ coefficients, with values near one indicating aligned directions. F1 used the three largest absolute coefficients, with a chance expectation of 0.50. The teacher and student shared one coefficient estimate, reported once. Twelve classical-model checks placed one positive coefficient in each of six positions, using daily one-day recall questionnaires with an error standard deviation of 0.02, noise-free RDoC profiles, and no state estimate from structured variables. Further diagnostics compared true states with true or noisy profiles and fitted states with noisy profiles across three seeds and both signal strengths (Supplementary Table 7).

We evaluated latent RMSE at test-patient sessions and scored questionnaires at the final eligible event per test patient and instrument, excluding retained questionnaires whose recall windows overlapped the target window. We selected models using questionnaire targets from separate validation patients. We calculated standard errors across three replicate seeds (Monte Carlo standard errors), treating the ten conditions as distinct settings.^43^

### Training-condition comparisons

We evaluated alternative training conditions by comparing BALL with a randomly initialized student trained on questionnaires with the teacher’s transitions fixed, a student initialized from the teacher and trained only to match its distributions, and a direct transformer that jointly learned its encoder and transitions. These comparisons change the initialization and updated parameters, as well as the loss terms. We evaluated the training conditions across nine simulations, spanning three settings and three seeds, using 600 student-stage epochs after the common 120-epoch warm-up and 300-epoch teacher fit. The focused 14-day clinical analysis retained its 120-epoch student schedule. Results appear in Supplementary Tables 5 and 20, with configurations in Supplementary Table 6.

A simulation sensitivity compared BALL and the direct transformer under shared stopping rules across three seeds in linear, nonlinear, and missing-feature settings with 10% signal. Training was allowed to run 600–1,500 epochs, checking the validation questionnaire error every 25 epochs. Four consecutive checks without at least a 0.0001 improvement halved the learning rate to a minimum of 0.00001, and 12 consecutive checks ended training. For each member, we retained the saved parameters with the lowest validation error. BALL retained a separate 300-epoch teacher fit (Supplementary Table 4).

### Negative controls for coefficient recovery

To test whether coefficient recovery depended on the relationship between RDoC scores and subsequent state changes,^44^ we compared a linear 25% signal positive control with zero signal, complete six-domain profiles shuffled across patient sessions, and independent standard-normal profile noise. Shuffling kept the six scores within each profile together.

Linear Gaussian, Markov, BALL, and direct-transformer coefficients were evaluated by cosine similarity and F1 against the positive-control vector. Four procedures, four controls, and three matched seeds produced 48 fits.

### Simulation sensitivity analyses

Across three seeds, we generated data and fitted models at persistence values of 0.1, 0.3, 0.6, and 0.9, testing all combinations in the linear setting and the setting with more missing variables and noisier RDoC scores. We selected fitted persistence by validation questionnaire error. The same settings were used to compare questionnaire weights of 10, 20, 40, and 80 during teacher training.

To isolate explicit RDoC coefficients, one sensitivity removed the six RDoC scores from the encoder but retained four correlated structured variables. A stricter version also removed those variables from encoders, the flexible part of the transition model, reconstruction of structured variables, and comparator regressions using those variables, leaving the explicit coefficient as the only RDoC pathway.

### Ordinal item-response measurement

An item-response sensitivity replaced normally distributed questionnaire totals with two nine-item questionnaires, each with four response categories, using a 14-day recall for depression and a 7-day recall for anxiety. Equations 27 and 28 map the latent trait to category probabilities using a graded-response model.^45^

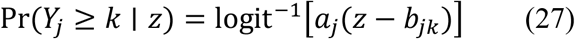

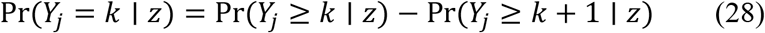

For item *j*, response *Y*_*j*_ takes values 0–3. Trait *z*, category boundary *k*, sensitivity to the trait (discrimination) *a*_*j*_, and trait threshold *b*_*jk*_ for boundary *k* determine response probabilities, with *P*(*Y*_*j*_ ≥ 0│*z*) = 1 and *P*(*Y*_*j*_ ≥ 4│*z*) = 0. The inverse-logit transformation converts the item’s latent response scale to a probability, and differences between adjacent cumulative probabilities yield category probabilities.

All methods used the same item responses, training and evaluation events, item parameters, and mapping to the latent trait scale. Classical models fitted parameters using item responses from training patients and updated state estimates using item likelihoods as earlier questionnaires became available during evaluation. BALL combined item likelihood with its variational and reconstruction objectives. Outcomes were channel-specific and joint trajectory error, item negative log-likelihood, error in expected total questionnaire scores, and coefficient recovery.

### Adaptive coefficient shrinkage

We tested whether applying larger penalties to coefficients with smaller initial estimates improved identification of the domains programmed to affect symptom change. We first fitted a teacher with its coefficient penalty set to zero, then used adaptive Lasso^46–47^ to set the penalties for a second fit. Equation 29 converts the initial coefficient estimates into penalty weights and rescales these weights to average one.

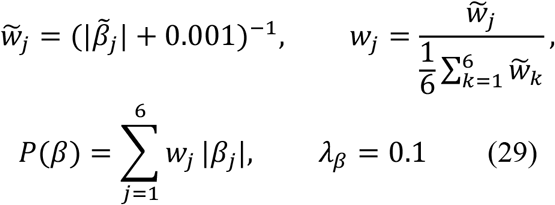

For domain *j*, initial coefficient estimate 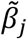 gives unnormalized weight 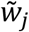 and normalized weight *w*_*j*_. We refitted with a penalty *P*(*β*) at strength *λ*_*β*_ of 0.1, holding the weights fixed and using the same patients and inputs.

### Empirical held-out questionnaire evaluation

All six prospective methods predicted the same withheld events using shared partitions and instrument links. Questionnaire predictions averaged previously issued channel estimates over the recall window, preserving those estimates when later observations became available.

Equation 30 averages the squared questionnaire errors within each patient before averaging across patients, giving each patient equal weight.

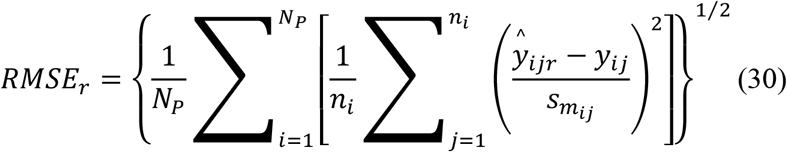

In Equation 30, *N*_*P*_ is the number of evaluated patients and *n*_*i*_ is the number of withheld questionnaires for the patient *i*. For event *j, ŷ_ijr_* is the prediction from the method *r* and *y*_*ij*_ the observed score. Dividing by 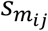, the standard deviation of that instrument’s scores in the training clinics, places errors on a comparable scale. We resampled patients with replacement 2,000 times to estimate uncertainty in paired differences between methods. We also examined BALL performance by questionnaire, amount of earlier questionnaire history, severity, treatment week, age, recorded sex, and clinic. We separately identified predictions made with fewer than two earlier questionnaires available (Supplementary Table 21).

### Performance in a later calendar period

For evaluation in a later calendar period at the 14-day schedule, we trained using patients from the 13 training and validation clinics whose records ended before 1 January 2025. Patients first observed on or after that date were assigned to separate calibration and test groups, excluding records spanning the cutoff. We refitted measurement links in pre-2025 training clinics and fitted BALL and both classical comparators using only pre-2025 training patients, retaining selected settings. Excluding 96 patients whose records spanned the cutoff left 847 patients for training, of whom 845 had usable retained questionnaires, and 227 later patients. We assigned 113 later patients to calibration and 114 to testing, with 113 contributing questionnaires for evaluation.

### Clinical sensitivity analyses

To assess the contribution of clinical history at 14 days, we refitted BALL after removing diagnoses and comorbidity, retaining questionnaires, session number, treatment, and elapsed time. A second version also removed treatment, leaving questionnaires, session number, and elapsed time.

For each schedule, we reported the fraction of questionnaires retained, questionnaire RMSE before restricting predictions to valid scores, and interval width as a fraction of the instrument’s range after applying those bounds.

We defined response as a ≥50% reduction from the first retained same-instrument score and remission as PHQ-9/GAD-7 ≤4 or BDI-II ≤13. Classification used predictions restricted to valid scores, equal patient weights, and 2,000 patient bootstrap samples.

### Adaptive questionnaire reassessment

We tested whether the expected information from an additional questionnaire could guide when to request it. We worked through each patient’s recorded questionnaires in chronological order, keeping the trained BALL models fixed and making scores available according to the original monitoring schedule. Before revealing a withheld score, we estimated how much it would reduce uncertainty about the average latent state over the questionnaire’s recall window. Under the Gaussian approximation, this proportional reduction is the latent variance on the questionnaire scale divided by the sum of latent and measurement variances.

We requested questionnaires when the expected reduction reached 25%, 50%, or 75%, requiring at least seven days since the latest available score from the same instrument. We used 50% as the representative threshold and added requested scores to the next session’s inputs. For comparison, we followed the original monitoring schedule or added every recorded questionnaire administered at least seven days after the latest available score from the same instrument.

We compared all collection policies at the same withheld questionnaires, predicting each score before revealing it, including scores requested by a policy. For each policy, we fitted prediction intervals using training patients and recalibrated them using reserved patients. We estimated uncertainty in paired RMSE differences using 2,000 patient bootstraps (Supplementary Table 23).

### Exploratory RDoC association analysis

RDoC defines six domains covering negative and positive valence, cognition, social processes, arousal/regulation, and sensorimotor functions.^26^ Scores described the attention devoted to each domain in the notes. After estimating clinical trajectories without these scores, we tested whether emphasis on domains in earlier notes was associated with subsequent estimated symptom change.

We used large language models (LLMs) to summarize clinical concepts in narrative fields, applying gemma-4-31B-it and Qwen3-Next-80B-A3B-Instruct to text outside structured diagnosis lists with a prompt adapted from McCoy and Perlis.^26^ Each model rated domain emphasis as 0, 0.5, or 1, after which we averaged across models and then across same-day fields and reported scorer agreement and score distributions. Positive valence includes reward processes relevant to anhedonia.^48^

From the 344 held-out patients, we included those with at least two retained questionnaires in the analyzed channel. We joined adjacent sessions on different calendar days, using the latest profile strictly before the current day and counting distinct daily profiles once in historical averages. We recorded which patients and pairs of sessions were excluded.

For each domain, we subtracted the average of all earlier distinct profiles from the latest profile, so positive differences indicated greater emphasis in the latest note than in that patient’s earlier notes. A sensitivity used the complete-record mean for retrospective association analysis, separately from prospective trajectory comparisons.

Equation 31 relates subsequent estimated change to current severity, elapsed days, time since the latest note, averages of earlier note scores, and differences between the latest scores and those averages, separately by channel.

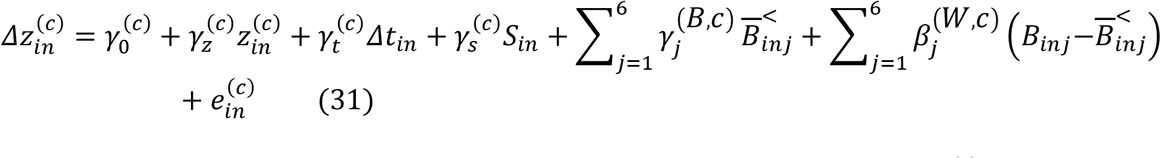

Indices *i, n, c*, and *j* denote patient, session, channel, and domain. Outcome 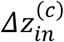 is a subsequent change, 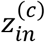 current severity, *Δt*_*in*_ elapsed days, and *S*_*in*_ note age. Latest score *B*_*inj*_ is compared with the prior mean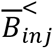, with gamma terms providing an intercept and adjustment for severity, timing, and earlier profiles. Coefficient 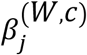 measures the association of the difference in domain *j*’s note score with subsequent symptom change, with residual 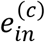.

We fitted separate channel regressions with a ridge penalty of 1, validating across five groups with each patient assigned to only one. Predictor scaling was fitted in the training groups. We compared the base model, using severity, elapsed days, time since the latest note, and averages of earlier note scores, with an extended model adding the six differences between the latest note scores and their earlier averages.

Using predictions for patients excluded from each fitted regression, we measured added value as the reduction in squared error after adding these differences in note scores. Dividing this reduction by the base-model error gave incremental *R*^2^. To assess whether this improvement depended on the timing of the notes, we repeated the comparison two hundred times after shifting these differences along each patient’s session sequence by a randomly chosen number of positions. Values moved past the end returned to the beginning, with all six domains shifted together and the earlier averages held fixed. We reported unadjusted probabilities and probabilities corrected by Holm’s method across the eight combinations of schedule and channel.

### Language-model scoring prompt

The complete scoring prompt used for both LLMs was the following. “You are a skilled psychiatrist scoring clinical notes in terms of how the notes reflect the 6 NIMH Research Domain Criteria (RDoC): negative valence systems, positive valence systems, cognitive systems, social processes, arousal and regulatory systems, and sensorimotor systems. Score 0 if concepts in a domain are not addressed, 0.5 if they are mentioned but not a focus, and 1 if they are a focus. A given note can have more than 1 focus. Please score explicit references to RDoC domains or symptoms or behaviors loading in that domain. Remember that substance use and addiction are reflected in the positive valence domain. Return your answer in the following format. DO NOT GIVE ANY EXPLANATION: negative valence systems: label positive valence systems: label cognitive systems: label social processes: label arousal and regulatory systems: label sensorimotor systems: label The optional list for label is [0, 0.5, 1].”

### Within-patient stability and between-patient variation

To distinguish changes within a patient from differences in patients’ overall severity, we compared within-patient change with between-patient differences using separate channel trajectories during days 0–84. The primary 14-day schedule grouped sessions into seven-day treatment weeks to compare patients at similar stages.

Equation 32 averages absolute changes between consecutive sessions on different calendar days, separately for each patient and channel. We assigned each pair to the treatment week of its earlier session.

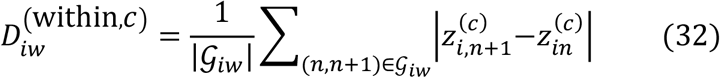

For patient *i*, week *w*, and channel *c, G*_*iw*_ contains the consecutive session pairs on different days for patient *i* in week *w*. Session *n* has severity 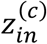. Smaller 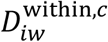 indicates less within-patient change.

Equation 33 compares patients’ weekly mean severity using the median absolute difference. We calculated each weekly mean from the state at the earlier session in each pair defined in Equation 32.

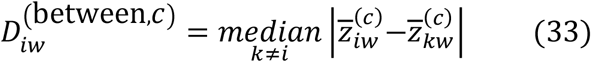

Here, 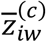 is patient *i*’s mean state at the earlier session in each pair assigned to week *w*, and 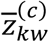 is the corresponding mean for patient *k*. Larger 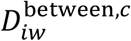 indicates greater between-patient separation.

We averaged each measure across the included weeks separately for each patient and channel and reported median within-patient change, between-patient difference, paired difference, and the proportion with greater between-patient difference. Weeks required ≥10 patients, and each patient needed ≥2 consecutive session pairs on different calendar days across the included weeks. We estimated uncertainty using 5,000 patient bootstraps and repeated the analysis at 7, 21, and 28 days to assess sensitivity.

## Data Availability

The repetitive transcranial magnetic stimulation cohort is governed as protected health information within Mass General Brigham. Requests for access may be directed to the corresponding author and require institutional and data-custodian approval, an executed data-use agreement, and analysis in an approved secure environment. The released code generates the simulation data deterministically from the replicate seeds listed in the benchmark parameter table.

## Code Availability

The public repository contains source code for BALL and the reported comparison models. It includes the directly trained forward-transformer comparisons under the standard and shared validation-based training rules, the student training-condition analysis, two-channel classical state-space comparators, channel-specific prospective Gaussian-process filters, and the exponential-decay gated recurrent unit. The repository also contains deterministic simulation generators, definitions of the clinical input variables, checks that predictions use only available information, and validation code that links each analysis to its manuscript text and display. Code is available at https://github.com/jbgibbons093/ball-paper-code.

### Acknowledgements

This work was supported by the National Institute on Drug Abuse under award 1K01DA061975 and the National Institute of Mental Health under award 1UG3MH141096-01. The funders had no role in study design, analysis, or the decision to submit.

## Author Contributions

JBG conceived and designed the study, developed and trained the BALL model, did the statistical analyses, and drafted the manuscript. NL, BPM, and ADM contributed to model development and analysis. RWo, MR, SS, SL, and DH provided the source clinical data and clinical operations expertise. All authors contributed to data interpretation and clinical or methodological expertise. PSW supervised the study as a co-senior author. JBG and PSW directly accessed and verified the underlying data. All authors critically revised the manuscript for important intellectual content, approved the final version, and accept responsibility for the decision to submit for publication.

## References

1. Fortney, J. C. et al. A tipping point for measurement-based care. Psychiatr. Serv. 68, 179–188 (2017). 10.1176/appi.ps.201500439.

2. Trivedi, M. H. et al. Evaluation of outcomes with citalopram for depression using measurement-based care in STAR*D: implications for clinical practice. Am. J. Psychiatry 163, 28–40 (2006). 10.1176/appi.ajp.163.1.28.

3. Kroenke, K., Spitzer, R. L. & Williams, J. B. The PHQ-9: validity of a brief depression severity measure. J. Gen. Intern. Med. 16, 606–613 (2001). 10.1046/j.1525-1497.2001.016009606.x.

4. Löwe, B., Unützer, J., Callahan, C. M., Perkins, A. J. & Kroenke, K. Monitoring depression treatment outcomes with the Patient Health Questionnaire-9. Med. Care 42, 1194–1201 (2004). 10.1097/00005650-200412000-00006.

5. Spitzer, R. L., Kroenke, K., Williams, J. B. & Löwe, B. A brief measure for assessing generalized anxiety disorder: the GAD-7. Arch. Intern. Med. 166, 1092–1097 (2006). 10.1001/archinte.166.10.1092.

6. Beck, A. T., Steer, R. A. & Brown, G. K. Beck Depression Inventory-II (BDI-II). APA PsycTests (1996). 10.1037/t00742-000.

7. Weiskopf, N. G. & Weng, C. Methods and dimensions of electronic health record data quality assessment: enabling reuse for clinical research. J. Am. Med. Inform. Assoc. 20, 144–151 (2013). 10.1136/amiajnl-2011-000681.

8. Rajkomar, A. et al. Scalable and accurate deep learning with electronic health records. npj Digit. Med. 1, 18 (2018). 10.1038/s41746-018-0029-1.

9. Durbin, J. & Koopman, S. J. Time Series Analysis by State Space Methods 2nd edn (Oxford University Press, 2012).

10. Kalman, R. E. A new approach to linear filtering and prediction problems. J. Basic Eng. 82, 35–45 (1960).

11. Rauch, H. E., Tung, F. & Striebel, C. T. Maximum likelihood estimates of linear dynamic systems. AIAA J. 3, 1445–1450 (1965).

12. Vaswani, A. et al. Attention is all you need. In Advances in Neural Information Processing Systems 30 (2017).

13. Li, Y. et al. BEHRT: transformer for electronic health records. Sci. Rep. 10, 7155 (2020). 10.1038/s41598-020-62922-y.

14. Shmatko, A., Jung, A. W., Gaurav, K. et al. Learning the natural history of human disease with generative transformers. Nature 647, 248–256 (2025). 10.1038/s41586-025-09529-3.

15. Krishnan, R. G., Shalit, U. & Sontag, D. Structured inference networks for nonlinear state-space models. Proc. AAAI Conf. Artif. Intell. 31 (2017). 10.1609/aaai.v31i1.10779.

16. Rubanova, Y., Chen, R. T. Q. & Duvenaud, D. Latent ordinary differential equations for irregularly sampled time series. In Advances in Neural Information Processing Systems 32 (2019).

17. Kidger, P., Morrill, J., Foster, J. & Lyons, T. Neural controlled differential equations for irregular time series. In Advances in Neural Information Processing Systems 33, 6696–6707 (2020).

18. Fortuin, V., Baranchuk, D., Rätsch, G. & Mandt, S. GP-VAE: deep probabilistic time-series imputation. Proc. 23rd Int. Conf. Artificial Intelligence and Statistics 108, 1651–1661 (PMLR, 2020).

19. Hinton, G., Vinyals, O. & Dean, J. Distilling the knowledge in a neural network. Preprint at https://arxiv.org/abs/1503.02531 (2015).

20. O’Reardon, J. P. et al. Efficacy and safety of transcranial magnetic stimulation in the acute treatment of major depression: a multisite randomized controlled trial. Biol. Psychiatry 62, 1208–1216 (2007). 10.1016/j.biopsych.2007.01.018.

21. George, M. S. et al. Daily left prefrontal transcranial magnetic stimulation therapy for major depressive disorder: a sham-controlled randomized trial. Arch. Gen. Psychiatry 67, 507–516 (2010). 10.1001/archgenpsychiatry.2010.46.

22. Blumberger, D. M. et al. Effectiveness of theta burst versus high-frequency repetitive transcranial magnetic stimulation in patients with depression (THREE-D): a randomised non-inferiority trial. Lancet 391, 1683–1692 (2018). 10.1016/S0140-6736(18)30295-2.

23. Insel, T. et al. Research Domain Criteria (RDoC): toward a new classification framework for research on mental disorders. Am. J. Psychiatry 167, 748–751 (2010). 10.1176/appi.ajp.2010.09091379.

24. Cuthbert, B. N. & Insel, T. R. Toward the future of psychiatric diagnosis: the seven pillars of RDoC. BMC Med. 11, 126 (2013). 10.1186/1741-7015-11-126.

25. McCoy, T. H. et al. A clinical perspective on the relevance of Research Domain Criteria in electronic health records. Am. J. Psychiatry 172, 316–320 (2015). 10.1176/appi.ajp.2014.14091177.

26. McCoy, T. H. & Perlis, R. H. Characterizing Research Domain Criteria symptoms among psychiatric inpatients using large language models. J. Mood Anxiety Disord. 8, 100079 (2024). 10.1016/j.xjmad.2024.100079.

27. Che, Z., Purushotham, S., Cho, K. et al. Recurrent neural networks for multivariate time series with missing values. Sci. Rep. 8, 6085 (2018). 10.1038/s41598-018-24271-9.

28. Little, R. J. A. Pattern-mixture models for multivariate incomplete data. J. Am. Stat. Assoc. 88, 125–134 (1993). 10.2307/2290705.

29. Rasmussen, C. E. & Williams, C. K. I. Gaussian Processes for Machine Learning (MIT Press, 2006).

30. Lakshminarayanan, B., Pritzel, A. & Blundell, C. Simple and scalable predictive uncertainty estimation using deep ensembles. In Advances in Neural Information Processing Systems 30 (2017).

31. MacKay, D. J. C. A practical Bayesian framework for backpropagation networks. Neural Comput. 4, 448–472 (1992). 10.1162/neco.1992.4.3.448.

32. Daxberger, E. et al. Laplace redux: effortless Bayesian deep learning. In Advances in Neural Information Processing Systems 34 (2021).

33. Vovk, V., Gammerman, A. & Shafer, G. Algorithmic Learning in a Random World (Springer, 2005). 10.1007/b106715.

34. Lei, J., G’Sell, M., Rinaldo, A., Tibshirani, R. J. & Wasserman, L. Distribution-free predictive inference for regression. J. Am. Stat. Assoc. 113, 1094–1111 (2018). 10.1080/01621459.2017.1307116.

35. Romano, Y., Patterson, E. & Candès, E. J. Conformalized quantile regression. In Advances in Neural Information Processing Systems 32, 3543–3553 (2019).

36. Caspi, A. et al. The p factor: one general psychopathology factor in the structure of psychiatric disorders? Clin. Psychol. Sci. 2, 119–137 (2014). 10.1177/2167702613497473.

37. Kotov, R. et al. The Hierarchical Taxonomy of Psychopathology (HiTOP): a dimensional alternative to traditional nosologies. J. Abnorm. Psychol. 126, 454–477 (2017). 10.1037/abn0000258.

38. Jordan, M. I., Ghahramani, Z., Jaakkola, T. S. & Saul, L. K. An introduction to variational methods for graphical models. Mach. Learn. 37, 183–233 (1999).

39. Kingma, D. P. & Welling, M. Auto-encoding variational Bayes. Preprint at https://arxiv.org/abs/1312.6114 (2014).

40. Hoerl, A. E. & Kennard, R. W. Ridge regression: biased estimation for nonorthogonal problems. Technometrics 12, 55–67 (1970).

41. Kingma, D. P. & Ba, J. Adam: a method for stochastic optimization. In Proc. 3rd International Conference on Learning Representations (2015).

42. Pascanu, R., Mikolov, T. & Bengio, Y. On the difficulty of training recurrent neural networks. Proc. 30th Int. Conf. Machine Learning 28, 1310–1318 (PMLR, 2013).

43. Morris, T. P., White, I. R. & Crowther, M. J. Using simulation studies to evaluate statistical methods. Stat. Med. 38, 2074–2102 (2019). 10.1002/sim.8086.

44. Lipsitch, M., Tchetgen Tchetgen, E. & Cohen, T. Negative controls: a tool for detecting confounding and bias in observational studies. Epidemiology 21, 383–388 (2010). 10.1097/EDE.0b013e3181d61eeb.

45. Samejima, F. Estimation of Latent Ability Using a Response Pattern of Graded Scores. Psychometric Monograph No. 17 (Psychometric Society, 1969).

46. Tibshirani, R. Regression shrinkage and selection via the lasso. J. R. Stat. Soc. Series B Stat. Methodol. 58, 267–288 (1996). 10.1111/j.2517-6161.1996.tb02080.x.

47. Zou, H. The adaptive lasso and its oracle properties. J. Am. Stat. Assoc. 101, 1418–1429 (2006). 10.1198/016214506000000735.

48. Pizzagalli, D. A. Depression, stress, and anhedonia: toward a synthesis and integrated model. Annu. Rev. Clin. Psychol. 10, 393–423 (2014). 10.1146/annurev-clinpsy-050212-185606.

