## Supplementary Information for "Bayesian Anchored Latent Learning for Estimating State Trajectories"

**Supplementary Table 1 | Primary simulation design, model settings, and computing environment.**

| Parameter | Value | Role |
|---|---|---|
| Patients per replicate | 150 | Cohort size in the simulation with a known RDoC coefficient |
| Follow-up length | 84 days | Length of each simulated longitudinal record |
| RDoC auxiliary scores | 6 | Number of auxiliary scores supplied at each modeled day |
| RDoC domains programmed to contribute to change | 3 | Scores assigned nonzero coefficients in the data-generating transition |
| RDoC component-variance ratio | 0.10 and 0.25 | RDoC contribution variance divided by the sum of symptom-change component variances, calculated within patients and pooled when patient subtypes differ |
| Simulation conditions | Linear, interaction, nonlinear, heterogeneous, and missingness with proxy noise | Five forms of data-generating dynamics |
| Replicate seeds | 2027, 2028, and 2029 | Three matched replicates for each simulation condition |
| Shared transformer architecture | 3 layers, width 96, 4 attention heads, dropout 0.10, an 8-dimensional treatment embedding, and transition-network width 32 | Architecture used by the teacher, student, and direct forward transformer |
| BALL teacher | Bidirectional attention | Uses the complete training record to estimate the trajectory at each session during training |
| BALL student | Forward-only attention | Estimates states from available history, learning from the teacher's depression and anxiety distributions and observed questionnaires |
| Direct transformer | Forward-only attention | Learns from observed questionnaires and the state-transition model using available history |
| Independent ensemble members | 5 | Independently initialized and optimized members |
| Trainable parameters per joint BALL member | 705,430 | Generative model, teacher, and student within one ensemble member |
| Trainable parameters per deployable BALL member | 353,758 | Generative model and forward-only student |
| Trainable parameters for the direct forward transformer | 353,758 per member and 1,768,790 across five members | Generative model and forward encoder |
| Trainable parameters across the BALL ensemble | 3,527,150 during joint training and 1,768,790 for prospective inference | Five joint members during training and five forward-only student members for prospective use |

| Parameter | Value | Role |
|---|---|---|
| Retrospective-teacher and forward-student training | 300 teacher epochs followed by 300 student epochs in the primary benchmark. Up to 1,500 student epochs in the convergence-controlled comparison | Forward-only student training follows fitting of its paired teacher member |
| Direct transformer training | 120 questionnaire-focused epochs at learning rate 0.002 and 300 variational epochs at learning rate 0.001 | Standard direct training uses the same number of training epochs as the teacher. The convergence-controlled comparison uses the same validation and stopping rule as the BALL student |
| Initial questionnaire-focused training | 120 epochs | Establishes latent scale before the full teacher or direct objective |
| Gradual Kullback-Leibler weighting | 80 epochs | Increases the contribution of the transition prior during the full objective |
| Forward-student learning rate | 0.001 | Optimization during prospective-student training |
| Student distribution-matching weight | 1 | Weight on the mean teacher-to-student divergence over valid patient-session positions |
| Student questionnaire-likelihood weight | 5 in simulation and 2 in the empirical analysis | Fixed during initial training and validation and held constant throughout each definitive evaluation |
| Small addition to the covariance diagonal | 0.000001 | Numerical stabilization for depression–anxiety covariance calculations |
| Student questionnaire objective | Posterior-predictive likelihood | Scores the questionnaire estimate and the predictive uncertainty over each recall window |
| Gradient-norm limit | 5 | Maximum gradient norm during neural-model optimization |
| Batch size | Full training cohort during questionnaire warm-up; 32 patients afterward | Batching for teacher, student, and direct-transformer optimization |
| Daily persistence of the fast latent component | 0.30 | Retention from one simulated day to the next |
| Penalty on the explicit RDoC coefficient | 0 | The primary benchmark estimates the explicit coefficient freely |
| Gaussian-process covariance allocation | 85 percent temporal and 15 percent auxiliary | Fixed allocation between time and auxiliary-input covariance |
| Gaussian-process temporal kernel | 70 percent squared-exponential with length selected from 14, 28, or 56 days and 30 percent Ornstein–Uhlenbeck with length selected from 1, 3, or 7 days | Channel-specific lengths selected by questionnaire marginal likelihood in training patients |

| Parameter | Value | Role |
|---|---|---|
| Gaussian-process auxiliary kernel | Squared-exponential kernel over RDoC, treatment, recent treatment, and treatment burden after standardization in training patients | Covariance induced by similarity in auxiliary inputs |
| Decay-GRU | Width 96, 300 epochs, learning rate 0.001, and smoothness weight 0.01 | Reduces the contribution of earlier information as time passes, then combines it with current inputs |
| Decay-GRU trainable parameters | 49,538 | Single forward-only neural comparator in each simulation replicate |
| Linear Gaussian | At least 3 and at most 25 alternating updates, convergence tolerance 0.0001, transition standard deviation 0.75, prior standard deviation 5.0, ridge penalty 10, and daily persistence 0.6 | Classical two-channel forward state-space filter |
| Markov | 3 groups based on prior questionnaire counts, 3 to 25 alternating training updates, convergence tolerance 0.0001, ridge penalty 10, and daily persistence 0.6 | Two-channel forward filter with group boundaries fitted in training patients |
| Patient partitions | 0.60, 0.10, 0.10, and 0.20 | Fractions used for training, validation, calibration of prediction intervals, and testing |
| Compute hardware | NVIDIA GeForce RTX 3070 Ti with 8 GB memory | Hardware used for the reported simulation runs |
| Mean runtime per replicate | Prospective reruns: linear Gaussian 16.80 s, Markov 17.28 s, Gaussian process 14.59 s | Recorded wall time with CPU thread environment limits set to two and sequential analysis jobs; not a comparison of computational efficiency |
| Software | Python 3.13.7, PyTorch 2.10.0 with CUDA 12.8, NumPy 2.3.4, pandas 2.3.3, and SciPy 1.16.3 | Software versions recorded for the simulation run |

**Notes**. The table lists the simulation design, model settings, optimization schedule, and computing environment. The teacher used complete training sequences during training. The BALL student and direct transformer used only information available by the session being estimated.

**Abbreviations**. BALL, Bayesian Anchored Latent Learning. CUDA, Compute Unified Device Architecture. Decay-GRU, exponential-decay gated recurrent unit. LGSSM, linear Gaussian state-space model. RDoC, Research Domain Criteria.

**Supplementary Table 2 | Simulation performance by setting and strength of the RDoC contribution.**

| Data-generating condition | Programmed auxiliary variance share | Model or component | Seeds | Composite trajectory RMSE | Held-out questionnaire RMSE | Cosine similarity | Top-three F1 |
|---|---|---|---|---|---|---|---|
| Linear auxiliary transition | 10% | BALL student | 3 | 0.715 (0.010) | 1.055 (0.074) | — | — |
| Linear auxiliary transition | 10% | BALL shared generative transition | 3 | — | — | 0.944 (0.008) | 0.778 (0.111) |
| Linear auxiliary transition | 10% | Direct transformer | 3 | 0.761 (0.012) | 1.111 (0.074) | 0.940 (0.024) | 0.889 (0.111) |
| Linear auxiliary transition | 10% | Exponential-decay gated recurrent unit | 3 | 1.244 (0.052) | 1.082 (0.080) | — | — |
| Linear auxiliary transition | 10% | Linear Gaussian state-space model | 3 | 1.580 (0.009) | 1.274 (0.091) | 0.534 (0.120) | 0.556 (0.111) |
| Linear auxiliary transition | 10% | Markov pattern-mixture model | 3 | 1.568 (0.012) | 1.242 (0.070) | 0.552 (0.116) | 0.556 (0.111) |
| Linear auxiliary transition | 10% | Causal Gaussian process | 3 | 1.430 (0.086) | 1.500 (0.081) | — | — |
| Linear auxiliary transition | 25% | BALL student | 3 | 0.836 (0.016) | 1.064 (0.051) | — | — |
| Linear auxiliary transition | 25% | BALL shared generative transition | 3 | — | — | 0.912 (0.023) | 0.889 (0.111) |
| Linear auxiliary transition | 25% | Direct transformer | 3 | 0.896 (0.018) | 1.139 (0.053) | 0.941 (0.020) | 0.778 (0.111) |
| Linear auxiliary transition | 25% | Exponential-decay gated recurrent unit | 3 | 1.502 (0.134) | 1.122 (0.004) | — | — |
| Linear auxiliary transition | 25% | Linear Gaussian state-space model | 3 | 2.411 (0.025) | 1.684 (0.270) | 0.873 (0.029) | 0.778 (0.111) |
| Linear auxiliary transition | 25% | Markov pattern-mixture model | 3 | 2.390 (0.016) | 1.656 (0.251) | 0.863 (0.027) | 0.667 (0.000) |
| Linear auxiliary transition | 25% | Causal Gaussian process | 3 | 2.210 (0.079) | 2.049 (0.152) | — | — |
| Interactions | 10% | BALL student | 3 | 0.698 (0.017) | 1.064 (0.068) | — | — |
| Interactions | 10% | BALL shared generative transition | 3 | — | — | 0.933 (0.012) | 0.889 (0.111) |
| Interactions | 10% | Direct transformer | 3 | 0.748 (0.017) | 1.103 (0.072) | 0.943 (0.032) | 0.889 (0.111) |

| Data-generating condition | Programmed auxiliary variance share | Model or component | Seeds | Composite trajectory RMSE | Held-out questionnaire RMSE | Cosine similarity | Top-three F1 |
|---|---|---|---|---|---|---|---|
| Interactions | 10% | Exponential-decay gated recurrent unit | 3 | 1.262 (0.024) | 1.167 (0.076) | — | — |
| Interactions | 10% | Linear Gaussian state-space model | 3 | 1.556 (0.015) | 1.208 (0.053) | -0.080 (0.246) | 0.444 (0.111) |
| Interactions | 10% | Markov pattern-mixture model | 3 | 1.554 (0.016) | 1.172 (0.025) | -0.124 (0.229) | 0.444 (0.111) |
| Interactions | 10% | Causal Gaussian process | 3 | 1.360 (0.066) | 1.416 (0.074) | — | — |
| Interactions | 25% | BALL student | 3 | 0.800 (0.015) | 1.055 (0.055) | — | — |
| Interactions | 25% | BALL shared generative transition | 3 | — | — | 0.941 (0.001) | 0.889 (0.111) |
| Interactions | 25% | Direct transformer | 3 | 0.866 (0.015) | 1.118 (0.055) | 0.952 (0.022) | 0.889 (0.111) |
| Interactions | 25% | Exponential-decay gated recurrent unit | 3 | 1.421 (0.066) | 1.256 (0.044) | — | — |
| Interactions | 25% | Linear Gaussian state-space model | 3 | 2.308 (0.052) | 1.420 (0.170) | 0.057 (0.280) | 0.444 (0.111) |
| Interactions | 25% | Markov pattern-mixture model | 3 | 2.303 (0.054) | 1.351 (0.124) | -0.015 (0.245) | 0.444 (0.111) |
| Interactions | 25% | Causal Gaussian process | 3 | 2.039 (0.104) | 1.616 (0.075) | — | — |
| Nonlinear dynamics | 10% | BALL student | 3 | 0.691 (0.015) | 1.068 (0.073) | — | — |
| Nonlinear dynamics | 10% | BALL shared generative transition | 3 | — | — | 0.930 (0.015) | 0.889 (0.111) |
| Nonlinear dynamics | 10% | Direct transformer | 3 | 0.744 (0.014) | 1.117 (0.072) | 0.938 (0.034) | 0.889 (0.111) |
| Nonlinear dynamics | 10% | Exponential-decay gated recurrent unit | 3 | 1.270 (0.064) | 1.200 (0.041) | — | — |
| Nonlinear dynamics | 10% | Linear Gaussian state-space model | 3 | 1.574 (0.025) | 1.207 (0.047) | 0.231 (0.102) | 0.222 (0.222) |
| Nonlinear dynamics | 10% | Markov pattern-mixture model | 3 | 1.573 (0.025) | 1.180 (0.025) | 0.170 (0.069) | 0.222 (0.222) |
| Nonlinear dynamics | 10% | Causal Gaussian process | 3 | 1.392 (0.071) | 1.429 (0.076) | — | — |

| **Data-generating condition** | **Programmed auxiliary variance share** | **Model or component** | **Seeds** | **Composite trajectory RMSE** | **Held-out questionnaire RMSE** | **Cosine similarity** | **Top-three F1** |
|---|---|---|---|---|---|---|---|
| Nonlinear dynamics | 25% | BALL student | 3 | 0.812 (0.020) | 1.072 (0.060) | — | — |
| Nonlinear dynamics | 25% | BALL shared generative transition | 3 | — | — | 0.939 (0.007) | 0.778 (0.111) |
| Nonlinear dynamics | 25% | Direct transformer | 3 | 0.873 (0.025) | 1.141 (0.062) | 0.944 (0.027) | 0.778 (0.111) |
| Nonlinear dynamics | 25% | Exponential-decay gated recurrent unit | 3 | 1.460 (0.043) | 1.201 (0.051) | — | — |
| Nonlinear dynamics | 25% | Linear Gaussian state-space model | 3 | 2.351 (0.068) | 1.425 (0.162) | 0.373 (0.095) | 0.444 (0.294) |
| Nonlinear dynamics | 25% | Markov pattern-mixture model | 3 | 2.348 (0.069) | 1.369 (0.124) | 0.308 (0.038) | 0.556 (0.222) |
| Nonlinear dynamics | 25% | Causal Gaussian process | 3 | 2.102 (0.116) | 1.635 (0.094) | — | — |
| Patient heterogeneity | 10% | BALL student | 3 | 0.709 (0.027) | 1.072 (0.066) | — | — |
| Patient heterogeneity | 10% | BALL shared generative transition | 3 | — | — | 0.885 (0.030) | 0.667 (0.192) |
| Patient heterogeneity | 10% | Direct transformer | 3 | 0.762 (0.023) | 1.118 (0.070) | 0.898 (0.036) | 0.778 (0.111) |
| Patient heterogeneity | 10% | Exponential-decay gated recurrent unit | 3 | 1.289 (0.025) | 1.147 (0.025) | — | — |
| Patient heterogeneity | 10% | Linear Gaussian state-space model | 3 | 1.587 (0.039) | 1.124 (0.012) | 0.321 (0.154) | 0.556 (0.111) |
| Patient heterogeneity | 10% | Markov pattern-mixture model | 3 | 1.585 (0.037) | 1.103 (0.023) | 0.256 (0.174) | 0.444 (0.111) |
| Patient heterogeneity | 10% | Causal Gaussian process | 3 | 1.427 (0.040) | 1.409 (0.056) | — | — |
| Patient heterogeneity | 25% | BALL student | 3 | 0.867 (0.032) | 1.058 (0.052) | — | — |
| Patient heterogeneity | 25% | BALL shared generative transition | 3 | — | — | 0.879 (0.058) | 0.778 (0.222) |

| **Data-generating condition** | **Programmed auxiliary variance share** | **Model or component** | **Seeds** | **Composite trajectory RMSE** | **Held-out questionnaire RMSE** | **Cosine similarity** | **Top-three F1** |
|---|---|---|---|---|---|---|---|
| Patient heterogeneity | 25% | Direct transformer | 3 | 0.984 (0.034) | 1.129 (0.058) | 0.897 (0.044) | 0.778 (0.111) |
| Patient heterogeneity | 25% | Exponential-decay gated recurrent unit | 3 | 1.515 (0.028) | 1.143 (0.018) | — | — |
| Patient heterogeneity | 25% | Linear Gaussian state-space model | 3 | 2.392 (0.064) | 1.295 (0.047) | 0.483 (0.126) | 0.556 (0.111) |
| Patient heterogeneity | 25% | Markov pattern-mixture model | 3 | 2.387 (0.062) | 1.248 (0.029) | 0.396 (0.146) | 0.556 (0.111) |
| Patient heterogeneity | 25% | Causal Gaussian process | 3 | 2.173 (0.062) | 1.581 (0.064) | — | — |
| Missing features and noisy profile | 10% | BALL student | 3 | 0.785 (0.004) | 1.085 (0.067) | — | — |
| Missing features and noisy profile | 10% | BALL shared generative transition | 3 | — | — | 0.840 (0.062) | 0.667 (0.000) |
| Missing features and noisy profile | 10% | Direct transformer | 3 | 0.828 (0.016) | 1.100 (0.067) | 0.800 (0.070) | 0.556 (0.111) |
| Missing features and noisy profile | 10% | Exponential-decay gated recurrent unit | 3 | 1.478 (0.045) | 1.113 (0.108) | — | — |
| Missing features and noisy profile | 10% | Linear Gaussian state-space model | 3 | 1.916 (0.037) | 1.508 (0.146) | 0.321 (0.195) | 0.667 (0.000) |
| Missing features and noisy profile | 10% | Markov pattern-mixture model | 3 | 1.896 (0.041) | 1.466 (0.120) | 0.322 (0.198) | 0.556 (0.111) |
| Missing features and noisy profile | 10% | Causal Gaussian process | 3 | 1.736 (0.102) | 1.699 (0.177) | — | — |
| Missing features and noisy profile | 25% | BALL student | 3 | 0.970 (0.009) | 1.111 (0.062) | — | — |
| Missing features and noisy profile | 25% | BALL shared generative transition | 3 | — | — | 0.741 (0.078) | 0.667 (0.000) |
| Missing features and noisy profile | 25% | Direct transformer | 3 | 1.014 (0.012) | 1.141 (0.048) | 0.881 (0.059) | 0.778 (0.111) |
| Missing features and noisy profile | 25% | Exponential-decay gated recurrent unit | 3 | 1.749 (0.193) | 1.177 (0.066) | — | — |

| Data-generating condition | Programmed auxiliary variance share | Model or component | Seeds | Composite trajectory RMSE | Held-out questionnaire RMSE | Cosine similarity | Top-three F1 |
|---|---|---|---|---|---|---|---|
| Missing features and noisy profile | 25% | Linear Gaussian state-space model | 3 | 3.067 (0.021) | 2.297 (0.370) | 0.786 (0.048) | 0.556 (0.111) |
| Missing features and noisy profile | 25% | Markov pattern-mixture model | 3 | 3.023 (0.023) | 2.229 (0.334) | 0.784 (0.043) | 0.556 (0.111) |
| Missing features and noisy profile | 25% | Causal Gaussian process | 3 | 2.695 (0.179) | 2.507 (0.348) | — | — |

**Notes**. Each cell gives the mean and, in parentheses, the Monte Carlo standard error across three seeds. Lower root mean squared error indicates better prediction. Higher cosine similarity indicates closer alignment with the programmed coefficient direction, and top-three F1 measures whether the three largest coefficients identify the three programmed domains. Selecting three of six domains gives an expected F1 of 0.50 by chance. A dash means the metric was not estimated for that row.

**Abbreviations**. BALL, Bayesian Anchored Latent Learning. Decay-GRU, exponential-decay gated recurrent unit. F1, harmonic mean of precision and recall. GP, Gaussian process. RDoC, Research Domain Criteria. RMSE, root mean squared error.

**Supplementary Table 3 | Depression- and anxiety-specific simulation performance pooled across prespecified conditions.**

| Model or component | Outcome | Descriptive mean | Descriptive SE across 30 simulation runs |
|---|---|---|---|
| BALL student | Depression latent RMSE | 0.888 | 0.016 |
| BALL student | Anxiety latent RMSE | 0.885 | 0.015 |
| BALL student | Depression-questionnaire RMSE | 1.195 | 0.019 |
| BALL student | Anxiety-questionnaire RMSE | 0.928 | 0.016 |
| Direct transformer | Depression latent RMSE | 0.942 | 0.016 |
| Direct transformer | Anxiety latent RMSE | 0.945 | 0.017 |
| Direct transformer | Depression-questionnaire RMSE | 1.264 | 0.021 |
| Direct transformer | Anxiety-questionnaire RMSE | 0.956 | 0.016 |
| Exponential-decay gated recurrent unit | Depression latent RMSE | 1.515 | 0.037 |
| Exponential-decay gated recurrent unit | Anxiety latent RMSE | 1.448 | 0.033 |
| Exponential-decay gated recurrent unit | Depression-questionnaire RMSE | 1.210 | 0.019 |
| Exponential-decay gated recurrent unit | Anxiety-questionnaire RMSE | 1.102 | 0.029 |
| Linear Gaussian state-space model | Depression latent RMSE | 2.186 | 0.092 |
| Linear Gaussian state-space model | Anxiety latent RMSE | 2.030 | 0.088 |
| Linear Gaussian state-space model | Depression-questionnaire RMSE | 1.568 | 0.075 |
| Linear Gaussian state-space model | Anxiety-questionnaire RMSE | 1.306 | 0.078 |
| Markov pattern-mixture model | Depression latent RMSE | 2.174 | 0.091 |
| Markov pattern-mixture model | Anxiety latent RMSE | 2.019 | 0.086 |
| Markov pattern-mixture model | Depression-questionnaire RMSE | 1.519 | 0.070 |
| Markov pattern-mixture model | Anxiety-questionnaire RMSE | 1.270 | 0.074 |
| Causal Gaussian process | Depression latent RMSE | 2.010 | 0.085 |
| Causal Gaussian process | Anxiety latent RMSE | 1.923 | 0.081 |
| Causal Gaussian process | Depression-questionnaire RMSE | 1.900 | 0.083 |

| Model or component | Outcome | Descriptive mean | Descriptive SE across 30 simulation runs |
|---|---|---|---|
| Causal Gaussian process | Anxiety-questionnaire RMSE | 1.435 | 0.061 |

**Notes**. Values are means and descriptive standard errors across 30 simulations. The first questionnaire measures depression, and the second measures anxiety. Lower root mean squared error indicates better prediction.

**Abbreviations**. BALL, Bayesian Anchored Latent Learning. Decay-GRU, exponential-decay gated recurrent unit. GP, Gaussian process. RMSE, root mean squared error. SE, standard error.

**Supplementary Table 4 | Paired simulation differences between the BALL student and comparison methods.**

| Comparison method | Evaluation target | Paired difference: BALL minus comparison | SE of paired difference | Runs with lower BALL error | Total paired runs | Relative error reduction for BALL, % |
|---|---|---|---|---|---|---|
| Direct transformer | Latent trajectory RMSE | -0.059 | 0.005 | 30 | 30 | 6.996 |
| Direct transformer | Held-out questionnaire RMSE | -0.051 | 0.005 | 29 | 30 | 4.583 |
| Exponential-decay gated recurrent unit | Latent trajectory RMSE | -0.631 | 0.027 | 30 | 30 | 44.444 |
| Exponential-decay gated recurrent unit | Held-out questionnaire RMSE | -0.090 | 0.017 | 26 | 30 | 7.789 |
| Linear Gaussian state-space model | Latent trajectory RMSE | -1.286 | 0.075 | 30 | 30 | 61.997 |
| Linear Gaussian state-space model | Held-out questionnaire RMSE | -0.374 | 0.082 | 29 | 30 | 25.889 |
| Markov pattern-mixture model | Latent trajectory RMSE | -1.274 | 0.074 | 30 | 30 | 61.782 |
| Markov pattern-mixture model | Held-out questionnaire RMSE | -0.331 | 0.076 | 29 | 30 | 23.632 |
| Causal Gaussian process | Latent trajectory RMSE | -1.068 | 0.069 | 30 | 30 | 57.533 |
| Causal Gaussian process | Held-out questionnaire RMSE | -0.614 | 0.067 | 30 | 30 | 36.442 |
| Direct transformer, shared validation stopping | Composite trajectory RMSE | -0.101 | 0.022 | 9 | 9 | 12.103 |

| Comparison method | Evaluation target | Paired difference: BALL minus comparison | SE of paired difference | Runs with lower BALL error | Total paired runs | Relative error reduction for BALL, % |
|---|---|---|---|---|---|---|
| Direct transformer, shared validation stopping | Held-out questionnaire RMSE | -0.026 | 0.019 | 6 | 9 | 2.422 |

**Notes**. Each difference compares BALL with the named method in matched simulations. Negative differences indicate lower error for BALL. Primary comparisons use 30 matched runs, and the comparison using the same stopping rule uses nine matched runs. All evaluated trajectories and questionnaire predictions use prospective information.

**Abbreviations**. BALL, Bayesian Anchored Latent Learning. Decay-GRU, exponential-decay gated recurrent unit. GP, Gaussian process. RMSE, root mean squared error. SE, standard error.

**Supplementary Table 5 | Performance of the different approaches to training the forward-only student.**

| **Forward-training configuration** | **Runs** | **Composite trajectory RMSE** | **SE** | **Questionnaire RMSE** | **SE** |
|---|---|---|---|---|---|
| Student trained on questionnaires with teacher transitions fixed | 9 | 0.770 | 0.022 | 1.111 | 0.033 |
| Student trained only to match teacher distributions | 9 | 0.737 | 0.013 | 1.099 | 0.037 |
| BALL: teacher initialization, posterior matching, and questionnaire supervision | 9 | 0.731 | 0.015 | 1.076 | 0.037 |

**Notes**. Values are means and standard errors across nine matched simulations. Each configuration used the same 120-epoch questionnaire warm-up, 300-epoch retrospective teacher fit, and 600 epochs of student training. The configurations differ in student initialization and training targets.

**Abbreviations**. BALL, Bayesian Anchored Latent Learning. RMSE, root mean squared error. SE, standard error.

**Supplementary Table 6 | Training configurations used to evaluate teacher-based forward learning.**

| Training configuration | Forward encoder initialization | Generative-model status | Teacher-posterior matching | Questionnaire objective | Updated parameters | Training schedule |
|---|---|---|---|---|---|---|
| Direct transformer | Random | Jointly fitted | None | Forward variational objective and questionnaire likelihood | Forward encoder and generative model | Questionnaire warm-up and variational schedule used for the teacher |
| Student trained on questionnaires with teacher transitions fixed | Random | Teacher-fitted generative model held fixed | None | Forward variational objective and questionnaire likelihood | Student encoder and its treatment embedding | 600 epochs in simulation and 120 clinically, after the common teacher fit |
| Student trained only to match teacher distributions | Initialized from paired retrospective teacher | Teacher-fitted generative model held fixed | Difference between teacher and student distributions over depression and anxiety | None | Student encoder and its treatment embedding | 600 epochs in simulation and 120 clinically, after the common teacher fit |
| BALL with teacher matching and questionnaire supervision | Initialized from paired retrospective teacher | Teacher-fitted generative model held fixed | Difference between teacher and student distributions over depression and anxiety | Posterior-predictive questionnaire likelihood | Student encoder and its treatment embedding | 600 epochs in simulation and 120 clinically, after the common teacher fit |

| Training configuration | Forward encoder initialization | Generative-model status | Teacher-posterior matching | Questionnaire objective | Updated parameters | Training schedule |
|---|---|---|---|---|---|---|
| Direct transformer with shared validation stopping | Random | Jointly fitted | None | Forward variational objective and questionnaire likelihood | Forward encoder and generative model | Up to 1,500 forward-training epochs with the same validation and stopping rules |

**Notes**. The three teacher-based configurations use the same fitted retrospective teacher and number of student-training epochs within each setting. The simulation training-condition analysis used 120 warm-up, 300 teacher, and 600 student-stage epochs, and the clinical analysis used 60, 120, and 120 epochs, respectively. The rows describe student initialization, the status of the generative model, training targets, updated parameters, and training schedule. The simulation comparison with shared stopping rules uses the same validation target, learning-rate schedule, minimum training duration, stopping rule, and rule for choosing saved model parameters for BALL and the direct transformer. Questionnaire instrument links were held fixed in every configuration.

**Abbreviations**. BALL, Bayesian Anchored Latent Learning.

**Supplementary Table 7 | Classical-model coefficient diagnostics and the consequences of measurement error.**

**Panel a. Simulation with one variable programmed to affect change**

| Programmed active coefficient | Model or component | Programmed position | Cosine similarity | Largest estimated position | Correct sign | Correct position | Converged |
|---|---|---|---|---|---|---|---|
| Coefficient 1 | Linear Gaussian | 0 | 1.0 | 0 | Yes | Yes | Yes |
| Coefficient 1 | Markov | 0 | 1.0 | 0 | Yes | Yes | Yes |
| Coefficient 2 | Linear Gaussian | 1 | 1.0 | 1 | Yes | Yes | Yes |
| Coefficient 2 | Markov | 1 | 1.0 | 1 | Yes | Yes | Yes |
| Coefficient 3 | Linear Gaussian | 2 | 1.0 | 2 | Yes | Yes | Yes |
| Coefficient 3 | Markov | 2 | 1.0 | 2 | Yes | Yes | Yes |
| Coefficient 4 | Linear Gaussian | 3 | 1.0 | 3 | Yes | Yes | Yes |
| Coefficient 4 | Markov | 3 | 1.0 | 3 | Yes | Yes | Yes |
| Coefficient 5 | Linear Gaussian | 4 | 1.0 | 4 | Yes | Yes | Yes |
| Coefficient 5 | Markov | 4 | 1.0 | 4 | Yes | Yes | Yes |
| Coefficient 6 | Linear Gaussian | 5 | 1.0 | 5 | Yes | Yes | Yes |
| Coefficient 6 | Markov | 5 | 1.0 | 5 | Yes | Yes | Yes |

**Panel b. Coefficient direction with error in RDoC scores and trajectories**

| Condition | Programmed variance share | State and auxiliary-profile inputs | Seeds | Mean cosine similarity | SE | Minimum | Maximum | Fraction with positive cosine |
|---|---|---|---|---|---|---|---|---|
| Linear auxiliary transition | 0.1 | Linear Gaussian estimated severity and observed profile | 3 | 0.534 | 0.120 | 0.296 | 0.685 | 1.000 |
| Linear auxiliary transition | 0.1 | Markov estimated severity and observed profile | 3 | 0.552 | 0.116 | 0.320 | 0.676 | 1.000 |
| Linear auxiliary transition | 0.1 | True severity and observed noisy RDoC profile | 3 | 0.691 | 0.091 | 0.509 | 0.795 | 1.000 |
| Linear auxiliary transition | 0.1 | True severity and true RDoC profile | 3 | 0.703 | 0.052 | 0.629 | 0.803 | 1.000 |
| Linear auxiliary transition | 0.25 | Linear Gaussian estimated severity and observed profile | 3 | 0.873 | 0.029 | 0.835 | 0.929 | 1.000 |
| Linear auxiliary transition | 0.25 | Markov estimated severity and observed profile | 3 | 0.863 | 0.027 | 0.829 | 0.917 | 1.000 |
| Linear auxiliary transition | 0.25 | True severity and observed noisy RDoC profile | 3 | 0.890 | 0.024 | 0.859 | 0.937 | 1.000 |
| Linear auxiliary transition | 0.25 | True severity and true RDoC profile | 3 | 0.863 | 0.018 | 0.828 | 0.890 | 1.000 |
| Missing features and noisy profile | 0.1 | True severity and observed noisy RDoC profile | 3 | 0.460 | 0.184 | 0.096 | 0.679 | 1.000 |
| Missing features and noisy profile | 0.1 | True severity and true RDoC profile | 3 | 0.703 | 0.052 | 0.629 | 0.803 | 1.000 |

| Condition | Programmed variance share | State and auxiliary-profile inputs | Seeds | Mean cosine similarity | SE | Minimum | Maximum | Fraction with positive cosine |
|---|---|---|---|---|---|---|---|---|
| Missing features and noisy profile | 0.25 | True severity and observed noisy RDoC profile | 3 | 0.803 | 0.042 | 0.744 | 0.883 | 1.000 |
| Missing features and noisy profile | 0.25 | True severity and true RDoC profile | 3 | 0.863 | 0.018 | 0.828 | 0.890 | 1.000 |

**Notes**. Panel a programs one active coefficient at a time. Panel b compares coefficient recovery using true or reconstructed trajectories and true or noisy auxiliary profiles. Panel b reports means and Monte Carlo standard errors across three seeds. Higher cosine similarity indicates closer directional recovery.

**Abbreviations**. SE, standard error.

**Supplementary Table 8 | Coefficient recovery when RDoC scores enter only through their transition coefficients.**

| Data-generating condition | Allowed auxiliary-information pathway | Seeds | Composite trajectory RMSE | Held-out questionnaire RMSE | Cosine similarity | Top-three F1 | Estimated coefficient norm |
|---|---|---|---|---|---|---|---|
| Linear transition | Observed profile enters only through the explicit coefficient; correlated proxies remain available | 3 | 0.845 (0.022) | 1.053 (0.073) | 0.911 (0.024) | 0.889 (0.111) | 0.214 (0.025) |
| Linear transition | Observed profile enters only through the explicit coefficient; correlated proxies removed from flexible pathways | 3 | 0.824 (0.025) | 1.042 (0.071) | 0.952 (0.012) | 0.889 (0.111) | 0.268 (0.011) |
| Missing features and noisy profile | Observed profile enters only through the explicit coefficient; correlated proxies remain available | 3 | 0.954 (0.006) | 1.081 (0.067) | 0.723 (0.081) | 0.667 (0.000) | 0.094 (0.016) |
| Missing features and noisy profile | Observed profile enters only through the explicit coefficient; correlated proxies removed from flexible pathways | 3 | 0.961 (0.001) | 1.085 (0.075) | 0.863 (0.050) | 0.889 (0.111) | 0.119 (0.010) |

**Notes**. The first restriction removes observed RDoC scores from the encoder and flexible part of the transition model but retains correlated structured variables. The second restriction also removes those correlated variables. Values are means and Monte Carlo standard errors across three seeds. Lower root mean squared error and higher cosine similarity and F1 indicate better performance. Selecting three of six domains gives an expected F1 of 0.50 by chance.

**Abbreviations**. BALL, Bayesian Anchored Latent Learning. F1, harmonic mean of precision and recall. RDoC, Research Domain Criteria. RMSE, root mean squared error.

**Supplementary Table 9 | Sensitivity of BALL to generating and fitted daily persistence.**

| Data-generating condition | Generating daily persistence | Fitted daily persistence | Seeds | Composite trajectory RMSE | Held-out questionnaire RMSE | Cosine similarity | Top-three F1 |
|---|---|---|---|---|---|---|---|
| Linear transition | 0.1 | 0.1 | 3 | 0.882 (0.017) | 1.081 (0.050) | 0.846 (0.040) | 0.667 (0.000) |
| Linear transition | 0.1 | 0.3 | 3 | 0.872 (0.023) | 1.080 (0.057) | 0.884 (0.026) | 0.778 (0.111) |
| Linear transition | 0.1 | 0.6 | 3 | 0.899 (0.020) | 1.084 (0.053) | 0.918 (0.011) | 0.889 (0.111) |
| Linear transition | 0.1 | 0.9 | 3 | 0.887 (0.028) | 1.078 (0.052) | 0.939 (0.009) | 0.889 (0.111) |
| Linear transition | 0.3 | 0.1 | 3 | 0.855 (0.014) | 1.080 (0.054) | 0.887 (0.036) | 0.889 (0.111) |
| Linear transition | 0.3 | 0.3 | 3 | 0.836 (0.016) | 1.064 (0.051) | 0.912 (0.023) | 0.889 (0.111) |
| Linear transition | 0.3 | 0.6 | 3 | 0.832 (0.023) | 1.057 (0.048) | 0.938 (0.011) | 0.889 (0.111) |
| Linear transition | 0.3 | 0.9 | 3 | 0.845 (0.013) | 1.067 (0.054) | 0.953 (0.005) | 1.000 (0.000) |
| Linear transition | 0.6 | 0.1 | 3 | 0.805 (0.017) | 1.064 (0.059) | 0.784 (0.082) | 0.667 (0.000) |
| Linear transition | 0.6 | 0.3 | 3 | 0.815 (0.025) | 1.069 (0.068) | 0.817 (0.062) | 0.667 (0.000) |
| Linear transition | 0.6 | 0.6 | 3 | 0.820 (0.020) | 1.064 (0.071) | 0.879 (0.027) | 0.778 (0.111) |
| Linear transition | 0.6 | 0.9 | 3 | 0.811 (0.016) | 1.080 (0.074) | 0.919 (0.012) | 0.889 (0.111) |
| Linear transition | 0.9 | 0.1 | 3 | 0.783 (0.023) | 1.064 (0.038) | 0.805 (0.095) | 0.778 (0.111) |
| Linear transition | 0.9 | 0.3 | 3 | 0.794 (0.014) | 1.061 (0.047) | 0.823 (0.081) | 0.778 (0.111) |
| Linear transition | 0.9 | 0.6 | 3 | 0.800 (0.018) | 1.063 (0.046) | 0.860 (0.060) | 0.889 (0.111) |
| Linear transition | 0.9 | 0.9 | 3 | 0.781 (0.014) | 1.060 (0.037) | 0.884 (0.047) | 0.889 (0.111) |

| **Data-generating condition** | **Generating daily persistence** | **Fitted daily persistence** | **Seeds** | **Composite trajectory RMSE** | **Held-out questionnaire RMSE** | **Cosine similarity** | **Top-three F1** |
|---|---|---|---|---|---|---|---|
| Missing features and noisy profile | 0.1 | 0.1 | 3 | 0.996 (0.016) | 1.100 (0.058) | 0.531 (0.040) | 0.556 (0.111) |
| Missing features and noisy profile | 0.1 | 0.3 | 3 | 0.991 (0.015) | 1.106 (0.056) | 0.622 (0.041) | 0.556 (0.111) |
| Missing features and noisy profile | 0.1 | 0.6 | 3 | 1.003 (0.012) | 1.109 (0.055) | 0.742 (0.047) | 0.667 (0.000) |
| Missing features and noisy profile | 0.1 | 0.9 | 3 | 0.980 (0.016) | 1.102 (0.057) | 0.797 (0.049) | 0.667 (0.000) |
| Missing features and noisy profile | 0.3 | 0.1 | 3 | 0.967 (0.011) | 1.100 (0.054) | 0.683 (0.095) | 0.667 (0.000) |
| Missing features and noisy profile | 0.3 | 0.3 | 3 | 0.970 (0.009) | 1.111 (0.062) | 0.741 (0.078) | 0.667 (0.000) |
| Missing features and noisy profile | 0.3 | 0.6 | 3 | 0.955 (0.001) | 1.096 (0.056) | 0.817 (0.041) | 0.667 (0.000) |
| Missing features and noisy profile | 0.3 | 0.9 | 3 | 0.956 (0.010) | 1.093 (0.065) | 0.841 (0.014) | 0.667 (0.000) |
| Missing features and noisy profile | 0.6 | 0.1 | 3 | 0.919 (0.011) | 1.090 (0.072) | 0.493 (0.137) | 0.444 (0.111) |
| Missing features and noisy profile | 0.6 | 0.3 | 3 | 0.917 (0.016) | 1.084 (0.072) | 0.571 (0.094) | 0.556 (0.111) |
| Missing features and noisy profile | 0.6 | 0.6 | 3 | 0.918 (0.009) | 1.078 (0.073) | 0.699 (0.041) | 0.667 (0.000) |

| Data-generating condition | Generating daily persistence | Fitted daily persistence | Seeds | Composite trajectory RMSE | Held-out questionnaire RMSE | Cosine similarity | Top-three F1 |
|---|---|---|---|---|---|---|---|
| Missing features and noisy profile | 0.6 | 0.9 | 3 | 0.929 (0.011) | 1.086 (0.071) | 0.773 (0.032) | 0.667 (0.000) |
| Missing features and noisy profile | 0.9 | 0.1 | 3 | 0.885 (0.016) | 1.063 (0.047) | 0.439 (0.205) | 0.333 (0.192) |
| Missing features and noisy profile | 0.9 | 0.3 | 3 | 0.884 (0.027) | 1.071 (0.038) | 0.473 (0.176) | 0.333 (0.192) |
| Missing features and noisy profile | 0.9 | 0.6 | 3 | 0.901 (0.028) | 1.079 (0.044) | 0.536 (0.137) | 0.556 (0.111) |
| Missing features and noisy profile | 0.9 | 0.9 | 3 | 0.890 (0.028) | 1.069 (0.037) | 0.571 (0.111) | 0.444 (0.111) |

**Notes**. Each row combines one persistence value used to generate the data with one used to fit the model in either the linear condition or the condition with greater missingness and noisier auxiliary scores. Values are means and Monte Carlo standard errors across three seeds. Lower root mean squared error and higher cosine similarity and F1 indicate better performance.

**Abbreviations**. BALL, Bayesian Anchored Latent Learning. F1, harmonic mean of precision and recall. RMSE, root mean squared error.

**Supplementary Table 10 | Validation-based selection of daily persistence for the classical simulation comparators.**

| Candidate daily persistence | Mean validation questionnaire RMSE | Median validation questionnaire RMSE | SD of validation questionnaire RMSE | Converged fits | Candidate fits | Selection result |
|---|---|---|---|---|---|---|
| 0.1 | 2.007 | 1.84 | 0.755 | 12 | 24 | |
| 0.3 | 1.787 | 1.647 | 0.58 | 18 | 24 | |
| 0.6 | 1.7 | 1.528 | 0.508 | 24 | 24 | Selected |
| 0.9 | 1.743 | 1.616 | 0.493 | 0 | 24 | |

**Notes.** Daily persistence was selected using questionnaire root mean squared error with equal weight per patient in validation simulations, with convergence required in all 24 fits. The selected value of 0.6 was fixed for the prospective classical comparisons.

**Abbreviations**. RMSE, root mean squared error. SD, standard deviation.

**Supplementary Table 11 | Sensitivity to the teacher questionnaire-likelihood weight.**

| Teacher questionnaire-likelihood weight | Runs | Composite trajectory RMSE | Held-out questionnaire RMSE | Cosine similarity | Top-three F1 |
|---|---|---|---|---|---|
| 10.0 | 6 | 0.968 (0.029) | 1.138 (0.055) | 0.847 (0.032) | 0.833 (0.075) |
| 20.0 | 6 | 0.919 (0.030) | 1.100 (0.041) | 0.854 (0.037) | 0.778 (0.070) |
| 40.0 | 6 | 0.903 (0.031) | 1.087 (0.038) | 0.826 (0.053) | 0.778 (0.070) |
| 80.0 | 6 | 0.929 (0.031) | 1.078 (0.030) | 0.805 (0.065) | 0.778 (0.070) |

**Notes**. Each row summarizes six simulations, including three with a linear transition and three with greater missingness and noisier auxiliary scores. Parentheses contain standard errors. The varied weight controls how strongly questionnaires contribute to the retrospective teacher's training objective.

**Abbreviations**. BALL, Bayesian Anchored Latent Learning. F1, harmonic mean of precision and recall. RMSE, root mean squared error. SE, standard error.

**Supplementary Table 12 | Simulation calibration and width of latent-state and questionnaire intervals.**

| Interval procedure | Seeds | Latent-state coverage | Latent interval width / latent-state SD | Questionnaire coverage | Questionnaire interval width / questionnaire SD |
|---|---|---|---|---|---|
| BALL calibrated with equal patient weights | 3 | 0.941 (0.002) | 0.447 (0.014) | 0.961 (0.004) | 0.904 (0.031) |
| BALL calibrated using known latent states | 3 | 0.950 (0.004) | 0.462 (0.008) | — | — |
| BALL before calibration | 3 | 0.920 (0.001) | 0.408 (0.013) | 0.939 (0.002) | 0.825 (0.029) |

**Notes**. Values are means and Monte Carlo standard errors across three seeds. Latent-state intervals are evaluated against known simulated trajectories, and questionnaire intervals are evaluated against held-out questionnaire scores. Relative width divides mean interval width by the target's standard deviation. Target coverage is 0.95. A dash means that the procedure did not produce a questionnaire interval.

**Abbreviations**. BALL, Bayesian Anchored Latent Learning. SD, standard deviation. SE, standard error.

**Supplementary Table 13 | Performance of BALL latent-state intervals before calibration by feature missingness and distance to an observed questionnaire.**

| Grouping variable | Group | Latent-state coverage before calibration | Width before calibration / latent-state SD | Evaluated latent states |
|---|---|---|---|---|
| Daily structured-feature missingness | 0.00-0.50 | 0.921 (0.001) | 0.397 (0.013) | 34004 |
| Daily structured-feature missingness | 0.50-0.70 | 0.907 (0.004) | 0.424 (0.012) | 12895 |
| Daily structured-feature missingness | 0.70-0.85 | 0.896 (0.012) | 0.453 (0.014) | 3307 |
| Daily structured-feature missingness | 0.85-1.01 | 0.932 (0.013) | 0.481 (0.008) | 194 |
| Calendar days to nearest observed questionnaire | 0-1 | 0.921 (0.002) | 0.399 (0.012) | 7054 |
| Calendar days to nearest observed questionnaire | 1-2 | 0.920 (0.003) | 0.405 (0.013) | 13518 |
| Calendar days to nearest observed questionnaire | 2-4 | 0.923 (0.001) | 0.413 (0.012) | 27036 |
| Calendar days to nearest observed questionnaire | 4-7 | 0.891 (0.003) | 0.402 (0.018) | 2568 |
| Calendar days to nearest observed questionnaire | 7-1000 | 0.879 (0.034) | 0.399 (0.021) | 224 |

**Notes**. Values describe BALL intervals before conformal calibration, with Monte Carlo standard errors across three seeds in parentheses. Structured-feature missingness is the fraction of unavailable structured inputs. Questionnaire distance is the number of calendar days to the nearest observed questionnaire. Counts pool the three seeds.

**Abbreviations**. BALL, Bayesian Anchored Latent Learning. SD, standard deviation. SE, standard error.

**Supplementary Table 14 | Positive and negative controls for the programmed auxiliary transition coefficient.**

| Control condition | Model or component | Cosine similarity | Top-three F1 | Estimated coefficient norm | Composite trajectory RMSE | Held-out questionnaire RMSE |
|---|---|---|---|---|---|---|
| Valid profile | BALL | 0.912 (0.023) | 0.889 (0.111) | 0.218 (0.025) | 0.836 (0.016) | 1.064 (0.051) |
| Valid profile | Direct transformer | 0.941 (0.020) | 0.778 (0.111) | 0.223 (0.006) | 0.896 (0.018) | 1.139 (0.053) |
| Valid profile | Linear Gaussian | 0.873 (0.029) | 0.778 (0.111) | 1.394 (0.183) | 2.411 (0.025) | 1.684 (0.270) |
| Valid profile | Markov | 0.863 (0.027) | 0.667 (0.000) | 1.405 (0.185) | 2.390 (0.016) | 1.656 (0.251) |
| No direct signal | BALL | -0.321 (0.134) | 0.667 (0.192) | 0.050 (0.008) | 0.529 (0.014) | 1.027 (0.055) |
| No direct signal | Direct transformer | -0.274 (0.145) | 0.444 (0.111) | 0.046 (0.003) | 0.556 (0.010) | 1.030 (0.052) |
| No direct signal | Linear Gaussian | -0.156 (0.145) | 0.444 (0.111) | 0.217 (0.007) | 0.931 (0.022) | 1.214 (0.085) |
| No direct signal | Markov | -0.119 (0.141) | 0.556 (0.111) | 0.209 (0.013) | 0.932 (0.023) | 1.210 (0.084) |
| Permuted profile | BALL | 0.163 (0.299) | 0.333 (0.192) | 0.041 (0.008) | 0.857 (0.026) | 1.069 (0.059) |
| Permuted profile | Direct transformer | -0.353 (0.125) | 0.333 (0.192) | 0.041 (0.003) | 0.902 (0.017) | 1.134 (0.065) |
| Permuted profile | Linear Gaussian | 0.302 (0.271) | 0.444 (0.111) | 0.398 (0.035) | 2.507 (0.048) | 1.834 (0.335) |
| Permuted profile | Markov | 0.325 (0.266) | 0.444 (0.111) | 0.395 (0.053) | 2.466 (0.019) | 1.731 (0.278) |
| Noise profile | BALL | 0.219 (0.262) | 0.667 (0.192) | 0.024 (0.003) | 0.852 (0.024) | 1.059 (0.066) |
| Noise profile | Direct transformer | 0.174 (0.323) | 0.667 (0.192) | 0.029 (0.007) | 0.921 (0.022) | 1.118 (0.055) |
| Noise profile | Linear Gaussian | 0.362 (0.251) | 0.556 (0.222) | 0.113 (0.016) | 2.503 (0.049) | 1.835 (0.337) |

| Control condition | Model or component | Cosine similarity | Top-three F1 | Estimated coefficient norm | Composite trajectory RMSE | Held-out questionnaire RMSE |
|---|---|---|---|---|---|---|
| Noise profile | Markov | 0.361 (0.277) | 0.556 (0.222) | 0.115 (0.016) | 2.462 (0.020) | 1.732 (0.278) |

**Notes**. Values are means and Monte Carlo standard errors across three matched seeds. The positive control retains an RDoC contribution scaled to 25% of the sum of the component variances in mean symptom change. The negative controls set this contribution to zero, shuffle complete profiles across sessions, or replace the profiles with independent noise. Coefficient metrics use the programmed positive-control coefficient as the reference. Selecting three of six domains gives an expected F1 of 0.50 by chance.

**Abbreviations**. BALL, Bayesian Anchored Latent Learning. F1, harmonic mean of precision and recall. RDoC, Research Domain Criteria. RMSE, root mean squared error.

**Supplementary Table 15 | Sensitivity of BALL to adaptive shrinkage of the auxiliary transition coefficient.**

| Coefficient specification | Model component | Composite trajectory RMSE | Held-out questionnaire RMSE | Cosine similarity | Top-three F1 |
|---|---|---|---|---|---|
| Primary unpenalized coefficient | BALL student | 0.788 (0.016) | 1.070 (0.017) | — | — |
| Primary unpenalized coefficient | BALL shared transition coefficient | — | — | 0.894 (0.015) | 0.789 (0.037) |
| Adaptive Lasso coefficient | BALL student | 0.793 (0.017) | 1.068 (0.017) | — | — |
| Adaptive Lasso coefficient | BALL shared transition coefficient | — | — | 0.895 (0.015) | 0.778 (0.037) |

**Notes**. The table compares the primary unpenalized BALL model with adaptive-Lasso refits of the same 30 simulations. Parentheses contain descriptive standard errors. Smaller initial coefficient estimates receive larger penalties. The weights use exponent 1, a stabilizing constant of 0.001, and a penalty of 0.1.

**Abbreviations**. BALL, Bayesian Anchored Latent Learning. F1, harmonic mean of precision and recall. RDoC, Research Domain Criteria. RMSE, root mean squared error.

**Supplementary Table 16 | Sensitivity using models of ordered questionnaire item responses.**

**Panel a. Overall performance across all data-generating conditions**

| Model or component | Depression-state RMSE | Anxiety-state RMSE | Joint latent-trait RMSE | Item negative log-likelihood | Expected-total RMSE | Cosine similarity | Top-three F1 |
|---|---|---|---|---|---|---|---|
| BALL student | 1.974 (0.159) | 1.814 (0.136) | 0.271 (0.018) | 0.996 (0.003) | 2.706 (0.048) | — | — |
| Linear Gaussian state-space model | 2.811 (0.086) | 2.500 (0.070) | 0.373 (0.011) | 0.993 (0.004) | 2.604 (0.072) | 0.322 (0.070) | 0.533 (0.038) |
| Markov pattern-mixture model | 2.798 (0.087) | 2.487 (0.069) | 0.371 (0.011) | 0.990 (0.004) | 2.565 (0.068) | 0.263 (0.075) | 0.500 (0.031) |
| BALL shared RDoC coefficient | — | — | — | — | — | 0.821 (0.043) | 0.778 (0.043) |

**Panel b. Joint latent-trait RMSE by data-generating condition**

| Data-generating condition | BALL student | Linear Gaussian | Markov |
|---|---|---|---|
| Linear auxiliary transition | 0.281 | 0.361 | 0.357 |
| Interactions | 0.275 | 0.359 | 0.358 |
| Nonlinear dynamics | 0.285 | 0.361 | 0.360 |
| Patient heterogeneity | 0.288 | 0.362 | 0.361 |
| Missing features and noisy profile | 0.226 | 0.422 | 0.418 |

**Notes**. This analysis replaces Gaussian total scores with two questionnaires with fixed models for ordered item responses, each containing nine items with four response categories. The depression instrument uses a 14-day recall window, and the anxiety instrument uses a 7-day window. State root mean squared error remains on the original simulated severity scale. Joint latent-trait error and the item metrics evaluate reproduction of questionnaire responses. Values are means and descriptive standard errors across 30 simulations.

**Abbreviations**. BALL, Bayesian Anchored Latent Learning. F1, harmonic mean of precision and recall. IRT, item response theory. RDoC, Research Domain Criteria. RMSE, root mean squared error.

**Supplementary Table 17 | Empirical cohort, clinic partitions, structured-feature availability, and note-score coverage.**

| Item | Value |
|---|---|
| **Source cohort** | |
| Patients | 1,609 |
| Sessions | 56,939 |
| Source network clinics | 25 |
| Distinct facility names in the analysis data | 23 |
| Clinic groups after reconciling facility names | 19 |
| Calendar window | 2 January 2023 to 27 June 2025 |
| Original source training partition | 914 patients from 10 clinics |
| Original source validation partition | 325 patients from 3 clinics |
| Original source test partition | 370 patients from 6 clinics |
| **Cohort after clinic exclusions** | |
| Patients with sessions spanning source clinic partitions | 94 |
| Primary analytic cohort after excluding records that crossed clinic partitions | 1,515 patients and 52,493 sessions |
| Patients used for training and validation | 1,170 patients from 13 clinics |
| Six clinic groups reserved for calibration and testing | 345 patients from 6 clinics, allocated to 172 calibration and 173 final test |
| **Modeled cohort** | |
| Patients without a retained questionnaire | 3 patients (2 from training and validation clinics, 1 calibration) |
| Final modeled patients | 1,512 |
| Patients used for final model training | 1,168 from 13 clinics |
| Final modeled calibration patients | 171 from the held-out clinics |
| Final modeled test patients | 173 from the held-out clinics |
| Calibration patients with withheld events, 7 / 14 / 21 / 28 days | 168 / 168 / 168 / 168 |
| Test patients with withheld events, 7 / 14 / 21 / 28 days | 170 / 170 / 171 / 171 |
| **Demographic and clinical summaries: cohort after clinic exclusions** | |
| Age in cohort after clinic exclusions, mean (SD), years | 39.0 (13.7) |
| Female sex in cohort after clinic exclusions, number (%) | 985 (65.0%) |
| Sessions per patient in analytic cohort, median (IQR) | 35 (29 to 37) |
| Baseline PHQ-9 in analytic cohort, mean (SD) | 12.4 (9.5), based on 1,511 patients |
| Baseline BDI-II in analytic cohort, mean (SD) | 23.0 (11.6), based on 1,471 patients |

| Item | Value |
|---|---|
| Baseline GAD-7 in analytic cohort, mean (SD) | 10.7 (5.3), based on 1,428 patients |
| Standard treatment protocol | 49,804 sessions among 1,465 patients |
| Theta-burst treatment protocol | 2,688 sessions among 346 patients |
| Bilateral treatment | 30,968 sessions among 1,206 patients |
| Unilateral treatment | 21,525 sessions among 1,020 patients |
| Patients and sessions with an earlier diagnosis list available by the final modeled session | 949 patients (62.6%) and 29,874 sessions (56.9%) |
| Non-depression psychiatric comorbidity count at final session, median (IQR) | 0 (0 to 1) |
| Recorded depression diagnosis by final session | 924 patients (61.0%) |
| Recorded anxiety diagnosis by final session | 454 patients (30.0%) |
| Recorded post-traumatic stress disorder diagnosis by final session | 100 patients (6.6%) |
| Recorded bipolar disorder diagnosis by final session | 9 patients (0.6%) |
| Recorded obsessive-compulsive disorder diagnosis by final session | 47 patients (3.1%) |
| Recorded attention-deficit/hyperactivity disorder diagnosis by final session | 74 patients (4.9%) |
| Recorded psychotic-spectrum disorder diagnosis by final session | 3 patients (0.2%) |
| Recorded substance-use disorder diagnosis by final session | 18 patients (1.2%) |
| Recorded eating disorder diagnosis by final session | 5 patients (0.3%) |
| Recorded personality disorder diagnosis by final session | 13 patients (0.9%) |
| Recorded autism-spectrum disorder diagnosis by final session | 5 patients (0.3%) |
| Recorded sleep disorder diagnosis by final session | 41 patients (2.7%) |
| **Note-score coverage: source cohort** | |
| Fraction of source-cohort sessions with an RDoC note-content emphasis score | 0.949 |
| Source-cohort patients with at least one RDoC note-content emphasis score | 1,524 |
| Mean within-patient RDoC note-content emphasis score SD | 0.080 |
| Mean between-patient RDoC note-content emphasis score SD | 0.205 |
| RDoC domains | Negative valence systems, Positive valence systems, Cognitive systems, Social processes, Arousal and regulatory systems, Sensorimotor systems |

| Item | Value |
| --- | --- |
| Empirical validation target | Recorded questionnaire scores withheld under each monitoring schedule |

**Notes.** Source-cohort counts describe all available patients and sessions. The cohort after clinic exclusions removes patients whose records cross the training, validation, or held-out clinic groups. The modeled cohort additionally requires a retained questionnaire under the evaluation schedules. Counts of patients contributing withheld events can be smaller than modeled counts and are reported separately by schedule. Demographic, treatment, and diagnosis summaries describe the cohort after clinic exclusions, and RDoC note-score coverage describes the source cohort. Treatment-protocol and laterality counts can overlap when treatment changes during a course.

**Abbreviations**. IQR, interquartile range. RDoC, Research Domain Criteria. rTMS, repetitive transcranial magnetic stimulation. SD, standard deviation.

**Supplementary Table 18 | Clinical severity bands on the fixed transformed questionnaire scales.**

| Channel | Instrument | Conventional severity band | Native score range | Fixed transformed range |
|---|---|---|---|---|
| Depression | PHQ-9 | Minimal or none | 0-4 | -1.00 to -0.70 |
| Depression | PHQ-9 | Mild | 5-9 | -0.63 to -0.33 |
| Depression | PHQ-9 | Moderate | 10-14 | -0.26 to 0.04 |
| Depression | PHQ-9 | Moderately severe | 15-19 | 0.11 to 0.41 |
| Depression | PHQ-9 | Severe | 20-27 | 0.48 to 1.00 |
| Depression | BDI-II | Minimal or none | 0-13 | -1.00 to -0.59 |
| Depression | BDI-II | Mild | 14-19 | -0.56 to -0.40 |
| Depression | BDI-II | Moderate | 20-28 | -0.37 to -0.11 |
| Depression | BDI-II | Severe | 29-63 | -0.08 to 1.00 |
| Anxiety | GAD-7 | Minimal or none | 0-4 | -1.00 to -0.62 |
| Anxiety | GAD-7 | Mild | 5-9 | -0.52 to -0.14 |
| Anxiety | GAD-7 | Moderate | 10-14 | -0.05 to 0.33 |
| Anxiety | GAD-7 | Severe | 15-21 | 0.43 to 1.00 |

**Notes**. Each questionnaire is transformed using its full possible score range. PHQ-9 uses $(score - 13.5) / 13.5$, GAD-7 uses $(score - 10.5) / 10.5$, and BDI-II uses $(score - 31.5) / 31.5$. Each transformation maps the minimum possible score to $-1$ and the maximum to 1.

**Abbreviations**. BDI-II, Beck Depression Inventory-II. GAD-7, Generalized Anxiety Disorder-7. PHQ-9, Patient Health Questionnaire-9.

**Supplementary Table 19 | Prospective questionnaire prediction and interval performance across four retention schedules.**

| Analysis or method | 7 days | 14 days | 21 days | 28 days | Information setting or metric |
|---|---|---|---|---|---|
| Panel a. RMSE with equal patient weights for withheld questionnaires | | | | | |
| BALL student | 0.598 | 0.654 | 0.669 | 0.718 | Prospective, with no later observations |
| Direct transformer | 0.577 | 0.640 | 0.689 | 0.743 | Prospective, with no later observations |
| Decay-GRU | 0.620 | 0.684 | 0.752 | 0.816 | Prospective, with no later observations |
| GP filter | 0.619 | 0.704 | 0.731 | 0.777 | Prospective, with no later observations |
| Linear Gaussian filter | 0.924 | 0.945 | 0.950 | 0.953 | Prospective, with no later observations |
| Markov filter | 0.928 | 0.948 | 0.953 | 0.956 | Prospective, with no later observations |
| Panel b. Paired RMSE differences, BALL minus comparison method | | | | | |
| Direct transformer | 0.021 [0.007, 0.035] | 0.014 [-0.002, 0.031] | -0.020 [-0.040, -0.001] | -0.026 [-0.048, -0.005] | Patient bootstrap, 2,000 samples |
| Decay-GRU | -0.022 [-0.048, 0.002] | -0.030 [-0.056, -0.004] | -0.083 [-0.111, -0.054] | -0.098 [-0.136, -0.059] | Patient bootstrap, 2,000 samples |
| GP filter | -0.021 [-0.056, 0.010] | -0.050 [-0.094, -0.011] | -0.062 [-0.102, -0.027] | -0.059 [-0.106, -0.018] | Patient bootstrap, 2,000 samples |
| Linear Gaussian filter | -0.326 [-0.378, -0.279] | -0.291 [-0.348, -0.242] | -0.281 [-0.333, -0.229] | -0.236 [-0.288, -0.185] | Patient bootstrap, 2,000 samples |
| Markov filter | -0.330 [-0.383, -0.283] | -0.294 [-0.351, -0.244] | -0.284 [-0.337, -0.232] | -0.238 [-0.291, -0.187] | Patient bootstrap, 2,000 samples |
| Panel c. Marginal questionnaire interval coverage | | | | | Calibration with equal patient weights |
| PHQ-9 | 0.970 | 0.951 | 0.952 | 0.956 | Marginal coverage |
| BDI-II | 0.977 | 0.966 | 0.965 | 0.972 | Marginal coverage |

| **Analysis or method** | **7 days** | **14 days** | **21 days** | **28 days** | **Information setting or metric** |
|---|---|---|---|---|---|
| GAD-7 | 0.954 | 0.944 | 0.961 | 0.959 | Marginal coverage |
| Panel d. Mean bounded interval width as a fraction of the instrument range | | | | | After applying valid score bounds |
| PHQ-9 | 0.426 | 0.500 | 0.549 | 0.603 | Width / possible score range |
| BDI-II | 0.294 | 0.343 | 0.337 | 0.407 | Width / possible score range |
| GAD-7 | 0.455 | 0.532 | 0.615 | 0.614 | Width / possible score range |
| Panel e. Questionnaire-retention fraction and threshold classification accuracy | | | | | |
| Fraction of genuine questionnaire events retained as model history | 0.322 | 0.191 | 0.140 | 0.112 | Fraction of available events |
| Response classification accuracy from BALL predictions | 0.839 to 0.860 | 0.827 to 0.834 | 0.809 to 0.836 | 0.790 to 0.831 | Range across eligible instruments |
| Remission classification accuracy from BALL predictions | 0.810 to 0.923 | 0.781 to 0.919 | 0.776 to 0.902 | 0.771 to 0.897 | Range across eligible instruments |

**Supplementary Table 19 (continued) | Sensitivity to the questionnaire-interval procedure.**

| **Analysis or method** | **7 days** | **14 days** | **21 days** | **28 days** | **Information setting or metric** |
|---|---|---|---|---|---|
| Panel f. Interval-procedure sensitivities | | | | | Coverage / bounded width fraction |
| Original conditional-quantile specification | 0.971 / 0.456 | 0.960 / 0.508 | 0.961 / 0.530 | 0.964 / 0.583 | All common withheld questionnaire events |
| Central intervals from fitted-patient residuals | 0.971 / 0.452 | 0.961 / 0.510 | 0.959 / 0.528 | 0.963 / 0.583 | All common withheld questionnaire events |
| Central intervals from excluded-patient residuals | 0.969 / 0.392 | 0.952 / 0.464 | 0.959 / 0.498 | 0.958 / 0.535 | All common withheld questionnaire events |
| Excluded-patient residuals with richer history | 0.970 / 0.400 | 0.955 / 0.459 | 0.959 / 0.497 | 0.958 / 0.532 | All common withheld questionnaire events |
| Richer history with shortest quantile intervals | 0.968 / 0.380 | 0.954 / 0.448 | 0.958 / 0.491 | 0.958 / 0.529 | All common withheld questionnaire events |
| Selected interval procedure | 0.970 / 0.390 | 0.954 / 0.457 | 0.958 / 0.498 | 0.961 / 0.541 | All common withheld questionnaire events |

**Notes.** Within each schedule, all methods use the same prospective information and predict the same withheld questionnaire events. Root mean squared error uses each instrument's standard deviation in training patients to put errors on a common scale and gives each patient equal weight. Retained questionnaires enter at the next recorded session. Linear Gaussian and Markov estimates use sequential updates using available history, with Markov groups based on earlier questionnaire counts. Paired differences use 2,000 patient bootstrap samples, and negative values favor BALL. Interval width after restricting limits to valid scores is divided by the instrument's possible score range. Panels c and d use the interval procedure selected using training and validation patients. The separate continuation compares the original conditional-quantile specification with four candidate procedures and the selected procedure. Selection used training and validation patients, followed by calibration in reserved patients and evaluation on common test events. Pooled coverage weights each patient equally, and instrument-specific coverage is calculated within instrument. BALL daily persistence, teacher questionnaire weight, and student questionnaire weight were fixed at 0.9, 10, and 2. Focused 14-day analyses and temporal results appear in Supplementary Table 20.

**Abbreviations**. BALL, Bayesian Anchored Latent Learning. BDI-II, Beck Depression Inventory-II. GAD-7, Generalized Anxiety Disorder-7. GP, Gaussian process. PHQ-9, Patient Health Questionnaire-9. RMSE, root mean squared error.

**Supplementary Table 20 | Focused analyses and temporal evaluation at the 14-day questionnaire schedule.**

| Analysis or model | Estimate | Metric |
|---|---|---|
| **Panel a. Focused BALL analyses at 14 days** | | |
| Full feature set | 0.654 | Questionnaire RMSE with equal patient weights |
| Questionnaires, timing, and treatment | 0.648 | Questionnaire RMSE with equal patient weights |
| Questionnaires and timing | 0.649 | Questionnaire RMSE with equal patient weights |
| Equal total questionnaire weight within a patient-session | 0.642 | Questionnaire RMSE with equal patient weights |
| Student trained on questionnaires with teacher transitions fixed | 0.654 | Questionnaire RMSE with equal patient weights |
| Student trained only to match teacher distributions | 0.664 | Questionnaire RMSE with equal patient weights |
| **Panel b. Prospective evaluation in patients first observed in 2025** | | |
| BALL student | 0.701 | Questionnaire RMSE with equal patient weights |
| Linear Gaussian filter | 1.038 | Questionnaire RMSE with equal patient weights |
| Markov filter | 1.040 | Questionnaire RMSE with equal patient weights |
| BALL minus linear Gaussian | -0.338 [-0.411, -0.265] | Paired RMSE difference [95% confidence interval] |
| BALL minus Markov | -0.340 [-0.413, -0.267] | Paired RMSE difference [95% confidence interval] |
| PHQ-9 interval coverage / width | 0.981 / 0.593 | Coverage / bounded mean width as a fraction of the instrument range |
| BDI-II interval coverage / width | 0.977 / 0.353 | Coverage / bounded mean width as a fraction of the instrument range |
| GAD-7 interval coverage / width | 0.997 / 0.752 | Coverage / bounded mean width as a fraction of the instrument range |
| **Panel c. Temporal interval-procedure sensitivities** | | |
| Original conditional-quantile specification | 0.983 / 0.567 | Coverage / bounded width fraction |
| Central intervals from fitted-patient residuals | 0.983 / 0.568 | Coverage / bounded width fraction |
| Central intervals from excluded-patient residuals | 0.983 / 0.510 | Coverage / bounded width fraction |
| Excluded-patient residuals with richer history | 0.980 / 0.507 | Coverage / bounded width fraction |
| Richer history with shortest quantile intervals | 0.977 / 0.503 | Coverage / bounded width fraction |

| Analysis or model | Estimate | Metric |
|---|---|---|
| Selected interval procedure | 0.978 / 0.511 | Coverage / bounded width fraction |

**Notes.** These analyses were specified for the 14-day schedule. Panel a reports the focused feature, weighting, and student training-condition analyses. Panel b evaluates the same 113 final-test patients first observed in 2025 using parameters fitted exclusively in pre-2025 training patients. All three models predict from prospective information and are scored on identical withheld questionnaire events. Paired differences use 2,000 patient bootstrap samples. Questionnaire interval coverage and widths apply to BALL. Panel c compares interval procedures fitted and selected in pre-2025 training and validation patients, calibrated in reserved later patients, and evaluated on the same final-test events. Coverage gives each patient equal weight within the evaluated instrument or pooled event set. Width is the width averaged across questionnaires after restricting intervals to valid scores divided by the instrument's possible score range. Central intervals use the 2.5th and 97.5th percentiles. The shortest-interval procedure selects among five predicted quantile pairs spanning 95 percentage points.

**Supplementary Table 21 | BALL questionnaire-prediction error across instruments and clinical subgroups.**

| Grouping variable | Group | 7 days | 14 days | 21 days | 28 days |
|---|---|---|---|---|---|
| Instrument | BDI-II | 0.620 | 0.661 | 0.671 | 0.706 |
| Instrument | GAD-7 | 0.659 | 0.684 | 0.669 | 0.693 |
| Instrument | PHQ-9 | 0.551 | 0.629 | 0.660 | 0.716 |
| Availability of earlier questionnaires | Two or more earlier questionnaires | 0.605 | 0.655 | 0.663 | 0.715 |
| Availability of earlier questionnaires | Fewer than two earlier questionnaires | 0.658 | 0.754 | 0.714 | 0.721 |
| Number of earlier questionnaires | One earlier questionnaire | 0.658 | 0.754 | 0.714 | 0.721 |
| Number of earlier questionnaires | Two earlier questionnaires | 0.663 | 0.723 | 0.700 | 0.733 |
| Number of earlier questionnaires | Three or more earlier questionnaires | 0.603 | 0.620 | 0.629 | 0.675 |
| First retained score on the same instrument | Below severe-symptom cutoff | 0.564 | 0.614 | 0.622 | 0.663 |
| First retained score on the same instrument | At or above severe-symptom cutoff | 0.675 | 0.734 | 0.756 | 0.792 |
| Treatment week | Week 1 | 0.591 | 0.680 | 0.680 | 0.693 |
| Treatment week | Week 2 | 0.720 | 0.778 | 0.796 | 0.795 |
| Treatment week | Week 3 | 0.611 | 0.659 | 0.707 | 0.735 |
| Treatment week | Week 4 | 0.590 | 0.670 | 0.654 | 0.764 |
| Treatment week | Week 5 or later | 0.577 | 0.612 | 0.624 | 0.689 |
| Recorded sex | Female | 0.610 | 0.674 | 0.685 | 0.731 |
| Recorded sex | Male | 0.573 | 0.610 | 0.635 | 0.689 |
| Age | 18 to 29 years | 0.605 | 0.660 | 0.663 | 0.725 |
| Age | 30 to 44 years | 0.623 | 0.678 | 0.690 | 0.735 |
| Age | 45 years or older | 0.560 | 0.617 | 0.647 | 0.689 |
| Held-out clinic group | Clinic 01 | 0.593 | 0.630 | 0.652 | 0.696 |
| Held-out clinic group | Clinic 02 | 0.602 | 0.658 | 0.668 | 0.701 |
| Held-out clinic group | Clinic 03 | 0.648 | 0.710 | 0.736 | 0.869 |
| Held-out clinic group | Clinic 04 | 0.605 | 0.676 | 0.688 | 0.735 |
| Held-out clinic group | Clinic 05 | 0.590 | 0.624 | 0.640 | 0.671 |

| Grouping variable | Group | 7 days | 14 days | 21 days | 28 days |
|---|---|---|---|---|---|
| Held-out clinic group | Clinic 06 | 0.384 | 0.521 | 0.528 | 0.512 |

**Notes.** Cells report root mean squared error for BALL, giving each patient equal weight and dividing errors by each instrument's standard deviation in training patients. Lower values indicate better prediction. Instrument rows pool standardized errors, and subgroup rows describe final-test patients from the six held-out clinics. Severity groups use the first retained score on the same instrument, with severe-symptom cutoffs of PHQ-9 ≥20, BDI-II ≥29, and GAD-7 ≥15. Events with no earlier score on the same instrument are excluded from these two rows. A patient can contribute different instruments to different severity groups.

**Abbreviations**. BALL, Bayesian Anchored Latent Learning. BDI-II, Beck Depression Inventory-II. GAD-7, Generalized Anxiety Disorder-7. PHQ-9, Patient Health Questionnaire-9. RMSE, root mean squared error.

**Supplementary Table 22 | Conditional coverage and width of BALL questionnaire prediction intervals.**

| Grouping variable | Group | 7 days (coverage / bounded width fraction) | 14 days (coverage / bounded width fraction) | 21 days (coverage / bounded width fraction) | 28 days (coverage / bounded width fraction) |
|---|---|---|---|---|---|
| Instrument | BDI-II | 0.977 / 0.294 | 0.966 / 0.343 | 0.965 / 0.337 | 0.972 / 0.407 |
| Instrument | GAD-7 | 0.954 / 0.455 | 0.944 / 0.532 | 0.961 / 0.615 | 0.959 / 0.614 |
| Instrument | PHQ-9 | 0.970 / 0.426 | 0.951 / 0.500 | 0.952 / 0.549 | 0.956 / 0.603 |
| Days since previous retained questionnaire | 0 to 1 day | 0.981 / 0.380 | 0.979 / 0.418 | 0.975 / 0.447 | 0.989 / 0.474 |
| Days since previous retained questionnaire | More than 1 to 2 days | 0.975 / 0.382 | 0.975 / 0.427 | 0.978 / 0.461 | 0.970 / 0.498 |
| Days since previous retained questionnaire | More than 2 to 4 days | 0.971 / 0.390 | 0.970 / 0.433 | 0.967 / 0.462 | 0.975 / 0.504 |
| Days since previous retained questionnaire | More than 4 to 7 days | 0.965 / 0.396 | 0.961 / 0.449 | 0.967 / 0.489 | 0.979 / 0.524 |
| Days since previous retained questionnaire | More than 7 days | — | 0.936 / 0.479 | 0.947 / 0.513 | 0.951 / 0.554 |
| Availability of earlier questionnaires | Two or more earlier questionnaires | 0.968 / 0.386 | 0.951 / 0.449 | 0.955 / 0.483 | 0.959 / 0.528 |
| Availability of earlier questionnaires | Fewer than two earlier questionnaires | 0.985 / 0.485 | 0.971 / 0.569 | 0.975 / 0.630 | 0.970 / 0.624 |
| Number of earlier questionnaires | One earlier questionnaire | 0.985 / 0.485 | 0.971 / 0.569 | 0.975 / 0.630 | 0.970 / 0.624 |
| Number of earlier questionnaires | Two earlier questionnaires | 0.953 / 0.466 | 0.945 / 0.510 | 0.960 / 0.541 | 0.956 / 0.559 |
| Number of earlier questionnaires | Three or more earlier questionnaires | 0.969 / 0.379 | 0.958 / 0.437 | 0.958 / 0.460 | 0.967 / 0.508 |
| First retained score on the same instrument | Below severe-symptom cutoff | 0.972 / 0.359 | 0.950 / 0.427 | 0.959 / 0.468 | 0.956 / 0.504 |
| First retained score on the same instrument | At or above severe-symptom cutoff | 0.956 / 0.440 | 0.950 / 0.507 | 0.953 / 0.547 | 0.960 / 0.603 |
| Treatment week | Week 1 | 0.956 / 0.440 | 0.960 / 0.457 | 0.963 / 0.476 | 0.964 / 0.490 |
| Treatment week | Week 2 | 0.986 / 0.450 | 0.963 / 0.543 | 0.974 / 0.566 | 0.974 / 0.570 |
| Treatment week | Week 3 | 0.974 / 0.413 | 0.978 / 0.452 | 0.966 / 0.560 | 0.971 / 0.576 |
| Treatment week | Week 4 | 0.969 / 0.383 | 0.953 / 0.491 | 0.967 / 0.449 | 0.952 / 0.585 |
| Treatment week | Week 5 or later | 0.967 / 0.375 | 0.951 / 0.437 | 0.954 / 0.484 | 0.961 / 0.525 |

| Grouping variable | Group | 7 days (coverage / bounded width fraction) | 14 days (coverage / bounded width fraction) | 21 days (coverage / bounded width fraction) | 28 days (coverage / bounded width fraction) |
|---|---|---|---|---|---|
| Recorded sex | Female | 0.974 / 0.397 | 0.957 / 0.464 | 0.958 / 0.503 | 0.965 / 0.548 |
| Recorded sex | Male | 0.960 / 0.370 | 0.947 / 0.437 | 0.960 / 0.481 | 0.954 / 0.520 |
| Age | 18 to 29 years | 0.972 / 0.403 | 0.963 / 0.467 | 0.973 / 0.508 | 0.969 / 0.556 |
| Age | 30 to 44 years | 0.961 / 0.391 | 0.946 / 0.462 | 0.945 / 0.502 | 0.958 / 0.540 |
| Age | 45 years or older | 0.979 / 0.374 | 0.955 / 0.438 | 0.963 / 0.480 | 0.958 / 0.525 |
| Held-out clinic group | Clinic 01 | 0.969 / 0.360 | 0.959 / 0.424 | 0.968 / 0.460 | 0.977 / 0.510 |
| Held-out clinic group | Clinic 02 | 0.966 / 0.393 | 0.955 / 0.460 | 0.960 / 0.498 | 0.966 / 0.537 |
| Held-out clinic group | Clinic 03 | 0.973 / 0.400 | 0.949 / 0.465 | 0.937 / 0.498 | 0.944 / 0.564 |
| Held-out clinic group | Clinic 04 | 0.970 / 0.400 | 0.949 / 0.470 | 0.949 / 0.512 | 0.947 / 0.549 |
| Held-out clinic group | Clinic 05 | 0.969 / 0.387 | 0.950 / 0.451 | 0.965 / 0.499 | 0.963 / 0.543 |
| Held-out clinic group | Clinic 06 | 0.996 / 0.408 | 0.991 / 0.484 | 1.000 / 0.515 | 1.000 / 0.575 |

**Notes.** Each cell reports coverage with each patient weighted equally, followed by mean interval width after restricting limits to valid scores as a fraction of the instrument's possible score range. Target overall coverage is 0.95. A dash means that the group and schedule contained no predictions meeting the analysis criteria. Severity groups use the first retained score on the same instrument, with severe-symptom cutoffs of PHQ-9 ≥20, BDI-II ≥29, and GAD-7 ≥15. This information is available before prediction. Events with no earlier score on the same instrument are excluded from these two rows. A patient can contribute different instruments to different severity groups.

**Abbreviations**. BALL, Bayesian Anchored Latent Learning. BDI-II, Beck Depression Inventory-II. GAD-7, Generalized Anxiety Disorder-7. PHQ-9, Patient Health Questionnaire-9.

**Supplementary Table 23 | Adaptive questionnaire collection and its effect on later predictions.**

| Questionnaire-retention schedule | Collection policy | Required reduction in state uncertainty | Fraction of additional questionnaires requested | Extra questionnaires per patient | RMSE on all evaluation events | Coverage on all evaluation events |
|---|---|---|---|---|---|---|
| 7 days | Original schedule | — | 0.000 | 0.00 | 0.598 | 0.970 |
| 7 days | Expected variance reduction | 25% | 0.000 | 0.00 | 0.598 | 0.970 |
| 7 days | Expected variance reduction | 50% | 0.000 | 0.00 | 0.598 | 0.970 |
| 7 days | Expected variance reduction | 75% | 0.000 | 0.00 | 0.598 | 0.970 |
| 7 days | Next available after seven days | — | 0.000 | 0.00 | 0.598 | 0.970 |
| 14 days | Original schedule | — | 0.000 | 0.00 | 0.654 | 0.954 |
| 14 days | Expected variance reduction | 25% | 0.107 | 7.80 | 0.623 | 0.961 |
| 14 days | Expected variance reduction | 50% | 0.101 | 7.31 | 0.626 | 0.961 |
| 14 days | Expected variance reduction | 75% | 0.055 | 3.98 | 0.631 | 0.960 |
| 14 days | Next available after seven days | — | 0.149 | 10.82 | 0.616 | 0.964 |
| 21 days | Original schedule | — | 0.000 | 0.00 | 0.669 | 0.958 |
| 21 days | Expected variance reduction | 25% | 0.141 | 10.80 | 0.634 | 0.960 |
| 21 days | Expected variance reduction | 50% | 0.129 | 9.88 | 0.637 | 0.960 |
| 21 days | Expected variance reduction | 75% | 0.071 | 5.42 | 0.644 | 0.960 |
| 21 days | Next available after seven days | — | 0.192 | 14.73 | 0.625 | 0.961 |
| 28 days | Original schedule | — | 0.000 | 0.00 | 0.718 | 0.961 |
| 28 days | Expected variance reduction | 25% | 0.156 | 12.31 | 0.655 | 0.949 |
| 28 days | Expected variance reduction | 50% | 0.146 | 11.50 | 0.658 | 0.950 |

| **Questionnaire-retention schedule** | **Collection policy** | **Required reduction in state uncertainty** | **Fraction of additional questionnaires requested** | **Extra questionnaires per patient** | **RMSE on all evaluation events** | **Coverage on all evaluation events** |
|---|---|---|---|---|---|---|
| 28 days | Expected variance reduction | 75% | 0.081 | 6.43 | 0.668 | 0.953 |
| 28 days | Next available after seven days | — | 0.213 | 16.84 | 0.641 | 0.951 |

**Notes.** Every policy starts with the original retained questionnaires and predicts all common withheld events before revealing any requested score. Additional scores enter the model at the next recorded session. All additional collection policies require at least seven days since the same instrument was collected. The variance rule requests a score when it is expected to reduce uncertainty about the average latent state over the questionnaire's recall window by the specified fraction, using a Gaussian approximation. Fractions use recorded questionnaire administrations withheld under the original schedule as the denominator. Counts per patient include patients with no requests. Prediction error and coverage include requested questionnaires, predicted before their scores became available, and later events influenced by those measurements. Intervals are recalibrated separately for each policy using reserved calibration patients. Requests were evaluated at sessions with recorded questionnaires. For the 50% rule, paired RMSE changes relative to the original schedule were −0.028 (95% confidence interval −0.038 to −0.018) at 14 days, −0.032 (−0.043 to −0.020) at 21 days, and −0.059 (−0.077 to −0.043) at 28 days, using 2,000 patient bootstrap samples. No additional measurements were selected at seven days.

**Abbreviations**. RMSE, root mean squared error.

**Supplementary Table 24 | Incremental prediction from earlier RDoC note-content changes in the six held-out clinics.**

**Panel a. Prediction using earlier note history**

| Schedule | Channel | Session pairs | Patients | Base RMSE | Augmented RMSE | RMSE improvement | Incremental $R^2$ |
|---|---|---|---|---|---|---|---|
| 7 days | Anxiety | 6,640 | 227 | 0.1312 | 0.1312 | -0.000053 | -0.000812 |
| 7 days | Depression | 6,709 | 235 | 0.1650 | 0.1652 | -0.000207 | -0.002513 |
| 14 days | Anxiety | 6,639 | 226 | 0.1212 | 0.1213 | -0.000062 | -0.001024 |
| 14 days | Depression | 6,709 | 235 | 0.1401 | 0.1402 | -0.000062 | -0.000889 |
| 21 days | Anxiety | 6,618 | 223 | 0.1047 | 0.1048 | -0.000026 | -0.000504 |
| 21 days | Depression | 6,709 | 235 | 0.1201 | 0.1202 | -0.000120 | -0.002004 |
| 28 days | Anxiety | 6,579 | 220 | 0.1100 | 0.1101 | -0.000122 | -0.002223 |
| 28 days | Depression | 6,709 | 235 | 0.1263 | 0.1265 | -0.000136 | -0.002149 |

**Panel b. Coefficients and tests using earlier note history**

| Schedule | Channel | Coefficient norm | Largest coefficient domain | Permutation P | Adjusted P |
|---|---|---|---|---|---|
| 7 days | Anxiety | 0.0052 | Cognitive systems | 0.0746 | 0.3731 |
| 7 days | Depression | 0.0062 | Social processes | 0.5274 | 1.0000 |
| 14 days | Anxiety | 0.0062 | Arousal and regulatory | 0.0149 | 0.1194 |
| 14 days | Depression | 0.0073 | Arousal and regulatory | 0.0299 | 0.1791 |
| 21 days | Anxiety | 0.0032 | Social processes | 0.0199 | 0.1393 |
| 21 days | Depression | 0.0037 | Cognitive systems | 0.1891 | 0.7562 |
| 28 days | Anxiety | 0.0038 | Arousal and regulatory | 0.7811 | 1.0000 |
| 28 days | Depression | 0.0062 | Arousal and regulatory | 0.2687 | 0.8060 |

**Panel c. Prediction using complete note history**

| Schedule | Channel | Session pairs | Patients | Base RMSE | Augmented RMSE | RMSE improvement | Incremental R² |
|---|---|---|---|---|---|---|---|
| 7 days | Anxiety | 11,184 | 311 | 0.1526 | 0.1526 | -0.000018 | -0.000232 |
| 7 days | Depression | 11,436 | 330 | 0.2269 | 0.2270 | -0.000145 | -0.001280 |
| 14 days | Anxiety | 11,140 | 308 | 0.1458 | 0.1458 | -0.000017 | -0.000231 |
| 14 days | Depression | 11,433 | 329 | 0.2015 | 0.2017 | -0.000135 | -0.001345 |
| 21 days | Anxiety | 11,129 | 305 | 0.1242 | 0.1243 | -0.000045 | -0.000727 |
| 21 days | Depression | 11,433 | 329 | 0.1852 | 0.1854 | -0.000184 | -0.001984 |
| 28 days | Anxiety | 10,990 | 297 | 0.1311 | 0.1311 | -0.000038 | -0.000582 |
| 28 days | Depression | 11,433 | 329 | 0.1981 | 0.1981 | -0.000086 | -0.000873 |

**Panel d. Coefficients and tests using complete note history**

| Schedule | Channel | Coefficient norm | Largest coefficient domain | Permutation P | Adjusted P |
|---|---|---|---|---|---|
| 7 days | Anxiety | 0.0034 | Sensorimotor | 0.2040 | 1.0000 |
| 7 days | Depression | 0.0043 | Sensorimotor | 0.7910 | 1.0000 |
| 14 days | Anxiety | 0.0042 | Arousal and regulatory | 0.2587 | 1.0000 |
| 14 days | Depression | 0.0040 | Positive valence | 0.8856 | 1.0000 |
| 21 days | Anxiety | 0.0028 | Sensorimotor | 0.6567 | 1.0000 |
| 21 days | Depression | 0.0027 | Sensorimotor | 0.9950 | 1.0000 |
| 28 days | Anxiety | 0.0019 | Sensorimotor | 0.5970 | 1.0000 |
| 28 days | Depression | 0.0037 | Positive valence | 0.6517 | 1.0000 |

**Notes.** We fitted ridge regressions in five rounds, each time predicting a different group of patients excluded from that fit. The base model includes current BALL-estimated severity, elapsed days, days since the most recent earlier note, and the patient’s mean across earlier note profiles. The augmented model adds six changes between the most recent earlier profile and that prior mean. Root mean squared error reduction is the base-model error minus the augmented-model error, so positive values favor the note-content terms. For two hundred repeated comparisons, we shifted the six note-score differences together along each patient’s session sequence, returning values past the end to the beginning and holding earlier averages fixed. These comparisons test whether improvement depends on the timing of the notes. Holm adjustment accounts for the eight combinations of schedule and latent channel. Panels a and b use earlier history only, and panels c and d use the complete record to define the history mean.

**Abbreviations**. BALL, Bayesian Anchored Latent Learning. RDoC, Research Domain Criteria. RMSE, root mean squared error.

**Supplementary Table 25 | Agreement between the two language-model scorers for RDoC note-content emphasis.**

| Domain | Pearson correlation (95% confidence interval) | Spearman rank correlation | Exact agreement, % | Quadratic weighted kappa | Mean absolute difference |
|---|---|---|---|---|---|
| negative valence | 0.863 (0.860, 0.865) | 0.857 | 83.8 | 0.862 | 0.085 |
| positive valence | 0.588 (0.582, 0.595) | 0.590 | 58.4 | 0.572 | 0.227 |
| cognitive systems | 0.765 (0.761, 0.769) | 0.746 | 71.5 | 0.750 | 0.147 |
| social processes | 0.839 (0.836, 0.841) | 0.827 | 83.2 | 0.837 | 0.084 |
| arousal/regulatory | 0.695 (0.689, 0.700) | 0.696 | 63.0 | 0.658 | 0.203 |
| sensorimotor | 0.619 (0.613, 0.625) | 0.588 | 83.7 | 0.500 | 0.090 |

**Notes**. Agreement is calculated across 39,556 paired narrative fields after excluding structured diagnosis lists. Each scorer assigns 0, 0.5, or 1 to each RDoC domain.

**Abbreviations**. RDoC, Research Domain Criteria.

**Supplementary Table 26 | Distribution of averaged RDoC note-content emphasis scores.**

| Domain | Scored narrative fields | Score 0, % | Score 0.25, % | Score 0.5, % | Score 0.75, % | Score 1, % |
|---|---|---|---|---|---|---|
| negative valence | 39,556 | 18.3 | 7.0 | 10.3 | 8.3 | 56.1 |
| positive valence | 39,556 | 42.5 | 20.5 | 10.8 | 17.3 | 8.9 |
| cognitive systems | 39,556 | 44.9 | 19.3 | 12.5 | 8.3 | 15.1 |
| social processes | 39,556 | 53.7 | 10.6 | 19.2 | 6.1 | 10.4 |
| arousal/regulatory | 39,556 | 18.9 | 17.2 | 17.5 | 16.3 | 30.1 |
| sensorimotor | 39,556 | 77.9 | 11.6 | 7.0 | 2.9 | 0.6 |

**Notes**. Each score is the average of the two ratings. Because each scorer assigns 0, 0.5, or 1, the possible averages are 0, 0.25, 0.5, 0.75, and 1. Percentages describe the scored narrative fields.

**Abbreviations**. RDoC, Research Domain Criteria.

**Supplementary Figure 1 | Coefficient recovery when RDoC scores enter only through their transition coefficients.**

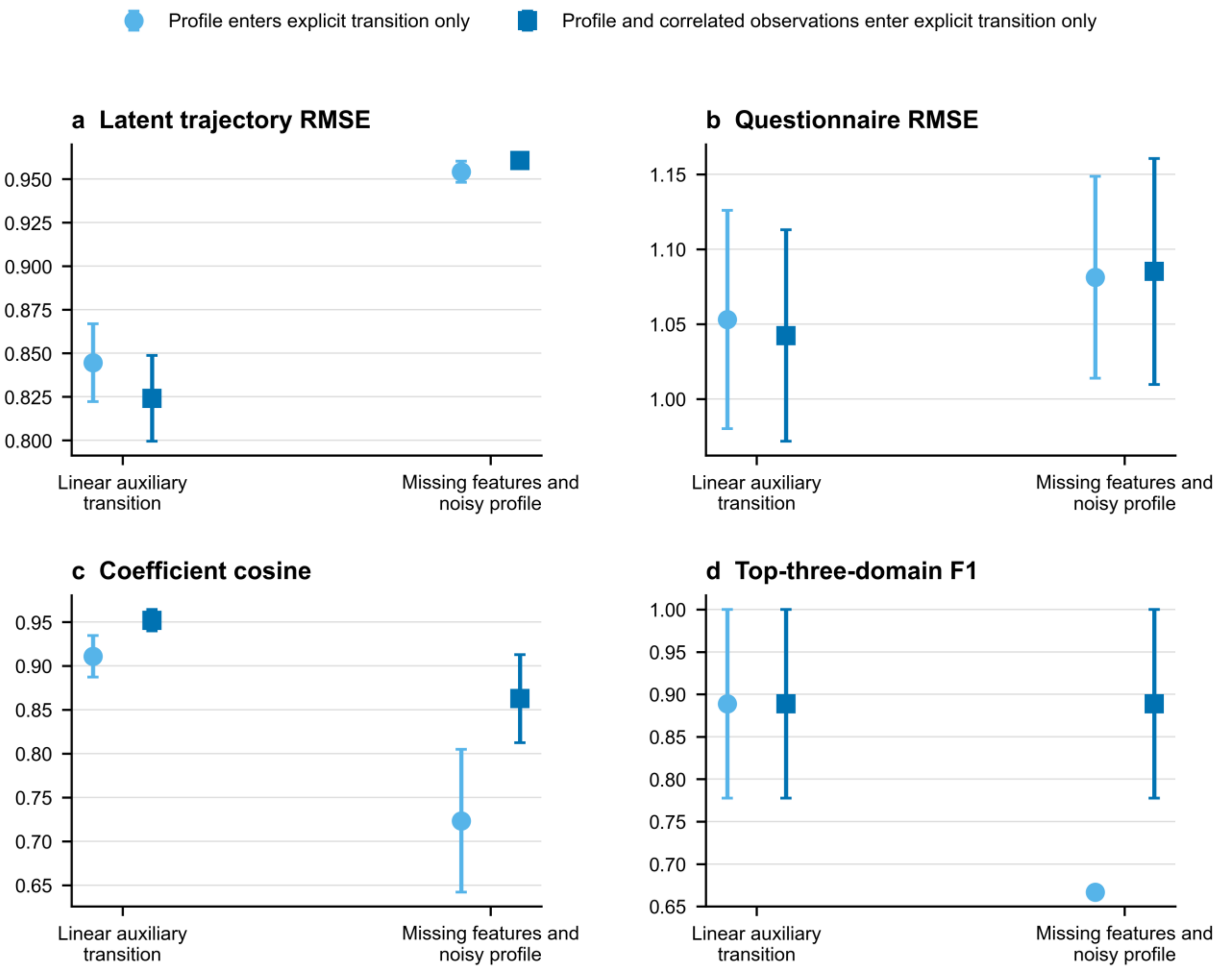


**Notes**. Panels a and b report latent-trajectory and held-out questionnaire error. Panels c and d report alignment with the programmed coefficient and identification of the three active domains. Points are means across three seeds, and error bars are Monte Carlo standard errors. The first restriction removes observed RDoC scores from encoder and flexible part of the transition model, and the second also removes correlated structured variables. Lower root mean squared error and higher cosine similarity and F1 indicate better performance. Selecting three of six domains gives an expected F1 of 0.50 by chance.

**Abbreviations**. BALL, Bayesian Anchored Latent Learning. F1, harmonic mean of precision and recall. RDoC, Research Domain Criteria. RMSE, root mean squared error.

**Supplementary Figure 2 | Sensitivity to daily persistence and teacher questionnaire weighting.**

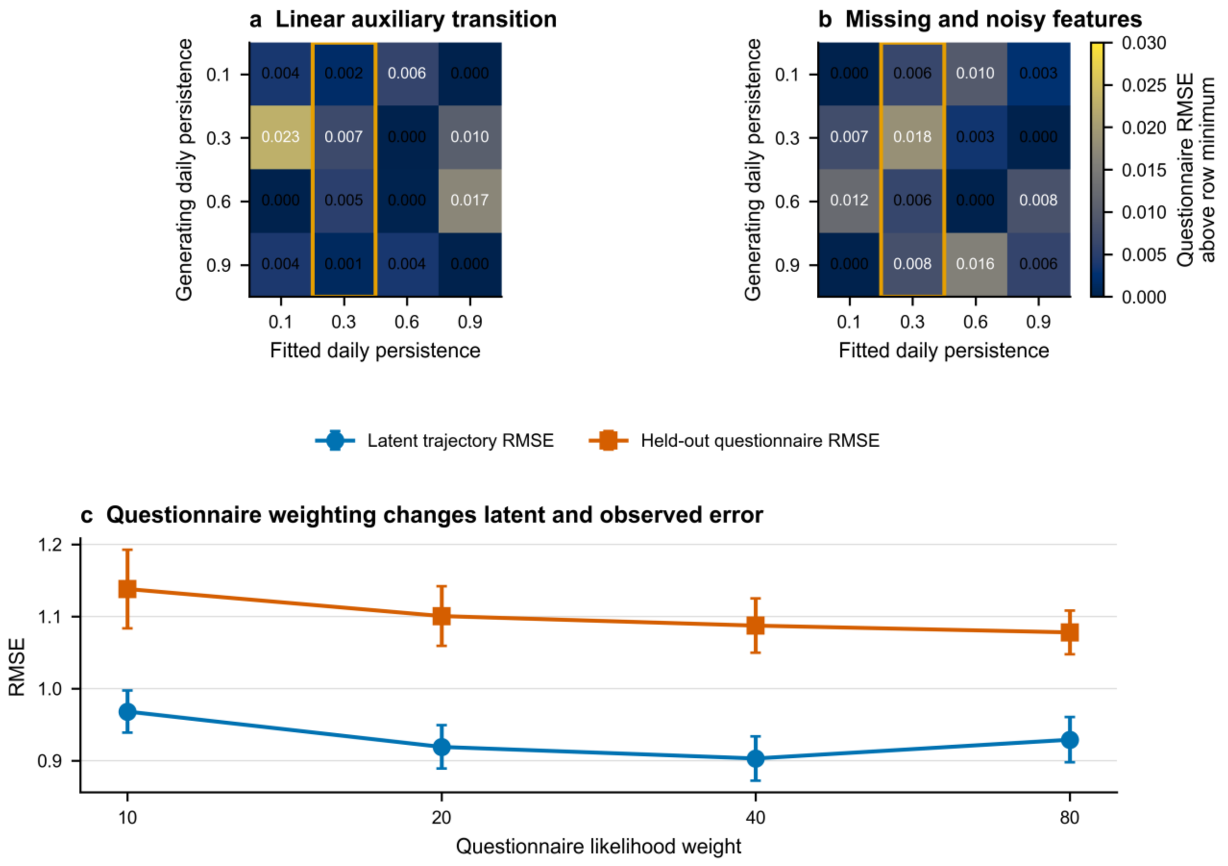


**Notes**. Panels a and b show the increase in held-out questionnaire root mean squared error relative to the best fitted persistence within each row, which fixes the persistence used to generate the data. The outlined column marks the primary fitted BALL persistence of 0.30. Panel c varies the questionnaire weight in the retrospective teacher objective. Points are means across three seeds in each condition, and error bars are Monte Carlo standard errors.

**Abbreviations**. BALL, Bayesian Anchored Latent Learning. RMSE, root mean squared error.

**Supplementary Figure 3 | Simulation calibration and conditional width of BALL latent-state intervals.**

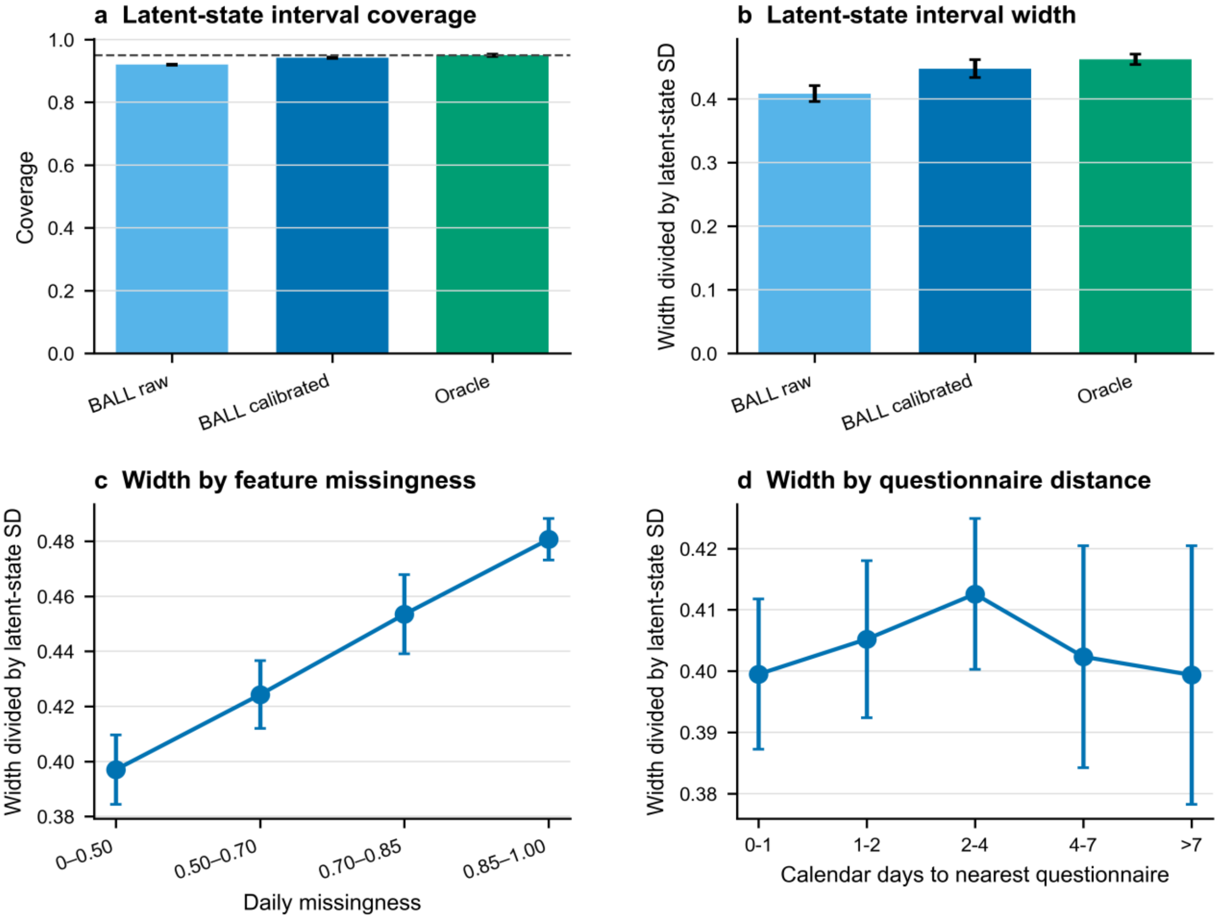


**Notes**. Panel a reports coverage of the known simulated trajectory, with the dashed line marking the 0.95 target. Panel b reports mean interval width divided by the latent-state standard deviation. “BALL raw” denotes intervals before calibration, and “Oracle” uses known latent states for calibration. Panels c and d show uncalibrated interval width by structured-feature missingness and distance to the nearest observed questionnaire. Bars and points are means across three seeds, and error bars are Monte Carlo standard errors. Supplementary Table 13 provides coverage and counts for these groups.

**Abbreviations**. BALL, Bayesian Anchored Latent Learning. SD, standard deviation.

**Supplementary Figure 4 | Positive and negative controls for the programmed auxiliary transition coefficient.**

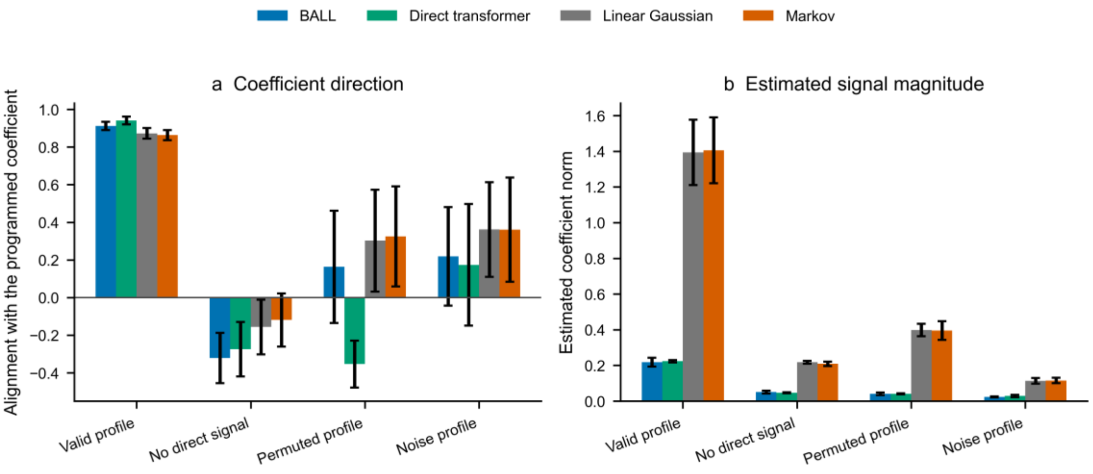


**Notes.** The positive control retained an RDoC contribution scaled to 25% of the sum of the component variances in mean symptom change. The zero-signal control removed the direct contribution. The permutation control shuffled profiles across sessions, moving all six domain scores together so that their relationships with one another were preserved. The noise control replaced the observed profile with independent values. Panel a reports the cosine similarity between each fitted coefficient and the direction programmed in the positive control. Panel b reports the overall size of the coefficients, measured by their Euclidean norm. Bars are means across three matched seeds, and error bars are Monte Carlo standard errors. BALL appears once because the teacher and student share the same generative coefficient.

**Abbreviations.** BALL means Bayesian Anchored Latent Learning, and RDoC means Research Domain Criteria.

**Supplementary Figure 5 | Focused clinical analyses and evaluation in a later patient cohort.**

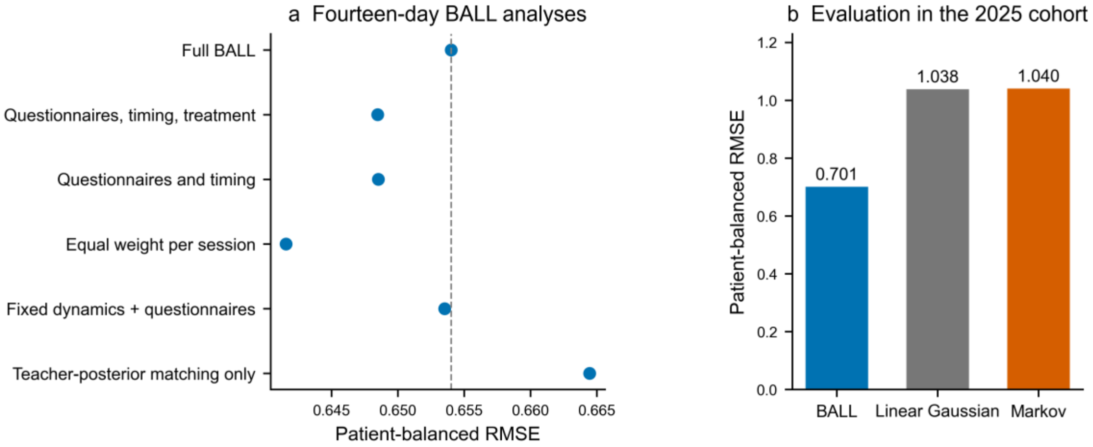


**Notes.** Panel a compares the full 14-day BALL model with models using fewer inputs, equal questionnaire weight per session, or different student-training targets. Panel b reports the 2025 evaluation among patients absent from the pre-2025 training data. Root mean squared error gives each patient equal total weight after normalization within instrument. The later-cohort comparison uses prospective predictions from BALL and the linear Gaussian and Markov filters on matched withheld events.

**Abbreviations**. BALL, Bayesian Anchored Latent Learning. BDI-II, Beck Depression Inventory-II. GAD-7, Generalized Anxiety Disorder-7. GP, Gaussian process. PHQ-9, Patient Health Questionnaire-9. RMSE, root mean squared error.

**Supplementary Figure 6 | Within-patient smoothness and between-patient separation of BALL estimates.**

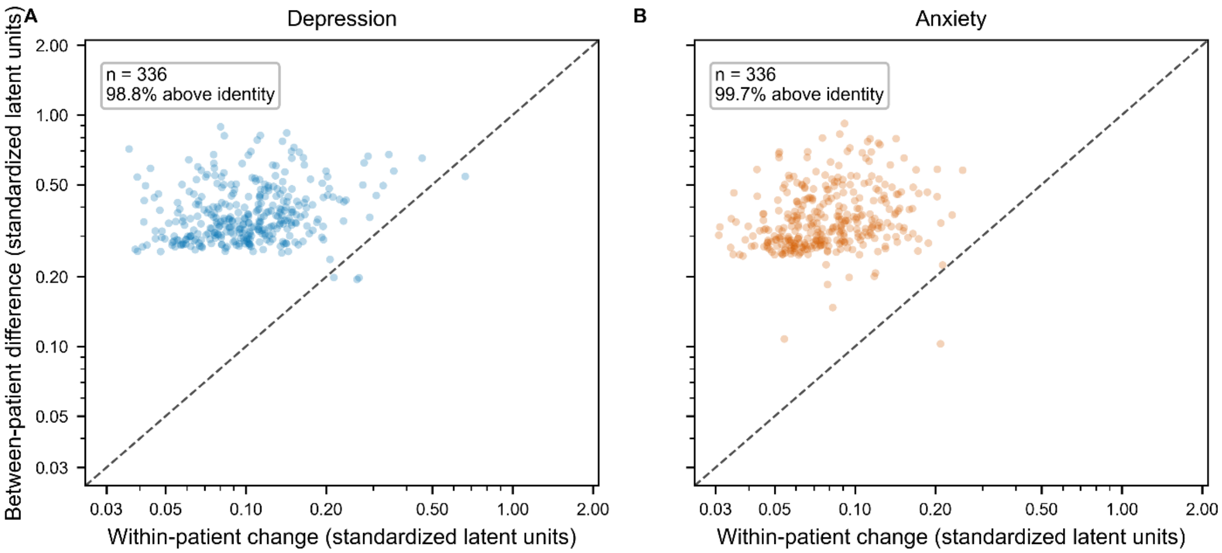


**Notes**. Panels a and b use BALL student estimates from the final 14-day model during treatment days 0 through 84. Each point represents one patient. The horizontal axis is the patient's mean absolute change between consecutive sessions on different calendar days, and the vertical axis is the patient's mean difference from other patients observed in the same treatment week. Each session pair is assigned to the treatment week of its earlier session, whose state enters the weekly mean. Both axes use logarithmic scales, and the dashed diagonal line marks equal values. Patient intervals use 5,000 bootstrap samples.

**Abbreviations**. BALL, Bayesian Anchored Latent Learning.

**Supplementary Figure 7 | Earlier RDoC note-content changes and subsequent BALL-estimated symptom change.**

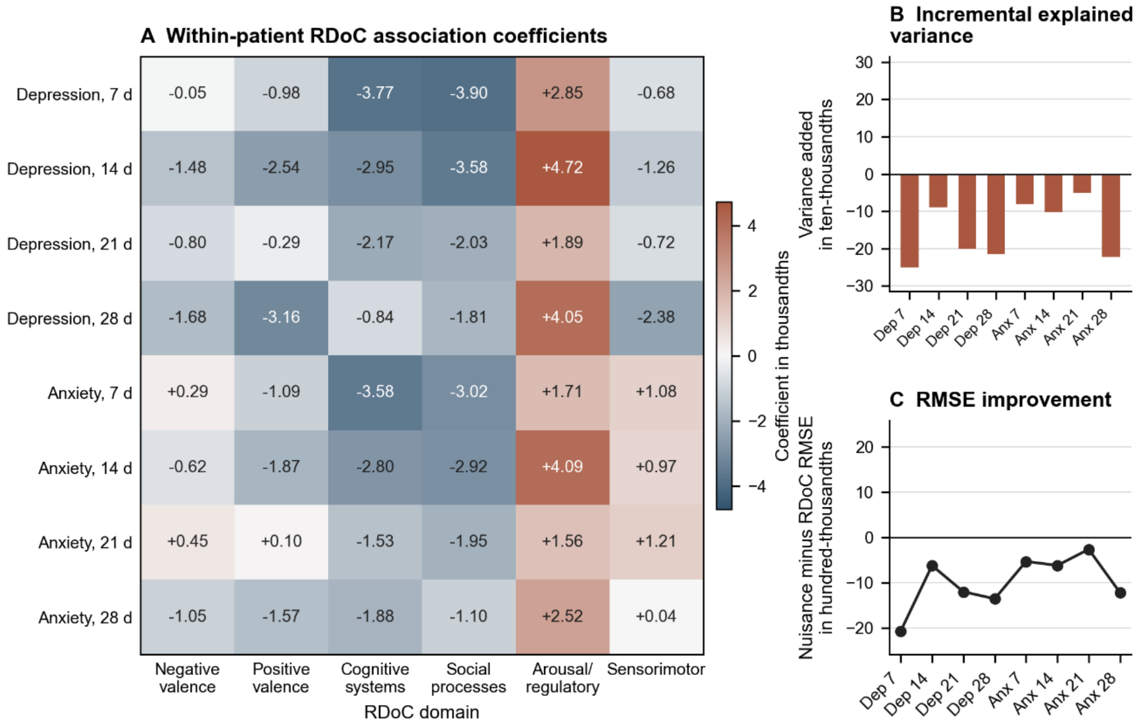


**Notes**. Panel a reports coefficients for the difference between the most recent earlier note profile and the patient's mean across earlier profiles, adjusted for current BALL-estimated severity, elapsed days, days since the note, and the earlier mean profile. Repeated sessions carrying the same note do not create additional profiles. Panel b reports the change in held-out explained variance after adding all six profile differences. Panel c reports base-model root mean squared error minus augmented-model error, so positive values favor the note-content terms. Predictions are evaluated in patients excluded from the regression fit. For 200 repeated comparisons, we shifted all six note-score differences together along each patient's session sequence, returning values past the end to the beginning and holding earlier averages fixed.

**Abbreviations**. BALL, Bayesian Anchored Latent Learning. LLM, large language model. RDoC, Research Domain Criteria. RMSE, root mean squared error. rTMS, repetitive transcranial magnetic stimulation.
